# The trustworthiness and impact of trial preprints for COVID-19 decision-making: A methodological study

**DOI:** 10.1101/2022.04.04.22273372

**Authors:** Dena Zeraatkar, Tyler Pitre, Gareth Leung, Ellen Cusano, Arnav Agarwal, Faran Khalid, Zaira Escamilla, Matthew Cooper, Maryam Ghadimi, Ying Wang, Francisca Verdugo, Gabriel Rada, Elena Kum, Anila Qasim, Jessica J Bartoszko, Reed Siemieniuk, Chirag J. Patel, Gordon Guyatt, Romina Brignardello-Petersen

**Affiliations:** Department of Biomedical Informatics, Harvard Medical School; Department of Health Research Methods, Evidence, and Impact, McMaster University; Department of Internal Medicine, McMaster University; Faculty of Medicine, University of Ottawa; Cumming School of Medicine, University of Calgary; Department of Medicine, McMaster University; Department of Medicine, University of Alberta; Epistemonikos Foundation, Santiago, Chile

## Abstract

**Purpose:** To assess the trustworthiness and impact of preprint trial reports during the COVID-19 pandemic.

**Data sources:** WHO COVID-19 database and the L-OVE COVID-19 platform by the Epistemonikos Foundation (up to August 3^rd^, 2021)

**Design:** We compare the characteristics of COVID-19 trials with and without preprints, estimate time to publication of COVID-19 preprint reports, describe discrepancies in key methods and results between preprint and published trial reports, report the number of retracted preprints and publications, and assess whether including versus excluding preprint reports affects meta-analytic estimates and the certainty of evidence. For the effects of eight therapies on mortality and mechanical ventilation, we performed meta-analyses including preprints and excluding preprints at 1 month, 3 months, and 6 months after the first trial addressing the therapy became available either as a preprint or publication (120 meta-analyses in total).

**Results:** We included 356 trials, 101 of which are only available as preprints, 181 as journal publications, and 74 as preprints first and subsequently published in journals. Half of all preprints remain unpublished at six months and a third at one year. There were few important differences in key methods and results between trial preprints and their subsequent published reports. We identified four retracted trials, three of which were published in peer-reviewed journals. With two exceptions (2/60; 3.3%), point estimates were consistent between meta-analyses including versus excluding preprints as to whether they indicated benefit, no appreciable effect, or harm. There were nine comparisons (9/60; 15%) for which the rating of the certainty of evidence differed when preprints were included versus excluded, for four of these comparisons the certainty of evidence including preprints was higher and for five of these comparisons the certainty of evidence including preprints was lower.

**Limitations:** The generalizability of our results is limited to COVID-19. Preprints that are subsequently published in journals may be the most rigorous and may not represent all trial preprints.

**Conclusion:** We found no compelling evidence that preprints provide less trustworthy results than published papers. We show that preprints remain the only source of findings of many trials for several months, a length of time that is unacceptable in a health emergency. We show that including preprints may affect the results of meta-analyses and the certainty of evidence. We encourage evidence users to consider data from preprints in contexts in which decisions are being made rapidly and evidence is being produced faster than can be peer-reviewed.

Summary Box 1

What is already known on this topic

- Clinicians and decision-makers need rapidly available and credible data addressing the comparative effectiveness of treatments and prophylaxis for COVID-19.
- Investigators have adopted preprint servers, which allow the rapid dissemination of research findings before publication in peer-reviewed journals.

What this study adds

- We found no compelling evidence that preprints provide less trustworthy results than published papers.
- We show that including preprints may affect the results of meta-analyses and the certainty of evidence and we encourage evidence users to consider data from preprints in contexts in which decisions are being made rapidly and evidence is being produced faster than can be peer-reviewed.

## Background

During the COVID-19 pandemic, the scientific community adopted preprint servers, which allow investigators to disseminate research findings before publication in peer-reviewed journals. Authors of seminal COVID-19 trials, for example, representing massive international collaborations, such as RECOVERY (1-4) and SOLIDARITY (5, 6), reported their results in preprints before subsequent publication in journals.

Growing interest in preprints predates the COVID-19 pandemic (7, 8). Researchers and evidence users have raised concerns that the traditional publication model is slow, peer review may not always improve the quality of manuscripts, journals impede dissemination due to paywalls and high publication fees and encourage publication bias by prioritizing statistically significant or anomalous findings—issues preprints may avoid (9-15). Despite these concerns, and the potential of preprints to address them, because preprints may result in the dissemination of provisional findings that contain important errors that, presumably, published papers do not, the medical community has been cautious regarding their adoption (16, 17).

Authors of systematic reviews, guideline developers, and other decision makers face a trade-off when considering preprints: on the one hand, including preprints could reduce the credibility of evidence syntheses and risk serious errors if important differences appear in published reports; on the other, including preprints may increase the precision of estimates; allow timely dissemination of research, and may minimize the effects of publication bias.

Knowledge of the extent to which preprints may accelerate the dissemination of findings, the frequency and nature of discrepancies between pre-prints and subsequent published reports, frequency of retractions of preprints compared to publications, and the impact preprints on meta-analytic estimates could inform the trade-off that evidence users face. Our study capitalizes on our living systematic reviews and network meta-analyses (SRNMAs) of drug treatments, antiviral antibodies and cellular therapies, and prophylaxis for COVID-19—an initiative launched in July 2020 that provides real-time summaries addressing the comparative effectiveness of treatments and prophylaxis for COVID-19—to report on the characteristics, trustworthiness—that is, complete and consistent reporting of key aspects of the methods and results between preprint and published trial reports—and impact of COVID-19 trial preprint reports (18-20).

## Methods

We submitted a protocol for this study for publication on September 7^th^, 2021. Because the protocol was still under review at the time the study was completed, we withdrew the protocol for publication and present it in Supplement 1.

### Search

Our study uses the search strategy of our living SRNMA that includes daily searches in the World Health Organization (WHO) COVID-19 database—a comprehensive multilingual source of global published and preprint literature on COVID-19 (https://search.bvsalud.org/global-literature-on-novel-coronavirus-2019-ncov/). Prior to its merge with the WHO COVID-19 database on 9 October 2020, we searched the US Centers for Disease Control and Prevention (CDC) COVID-19 Research Articles Downloadable Database. A validated machine learning model facilitates efficient identification of randomized trials (21).

Our search is supplemented by ongoing surveillance of living evidence retrieval services, including the Living Overview of the Evidence (L-OVE) COVID-19 platform by the Epistemonikos Foundation (https://app.iloveevidence.com/loves/5e6fdb9669c00e4ac072701d) and the Systematic and Living Map on COVID-19 Evidence by the Norwegian Institute of Public Health (https://www.fhi.no/en/qk/systematic-reviews-hta/map/). Using the above sources, we also monitor for retraction notices and concerns regarding trial integrity. Supplement 2 includes additional details of our search strategy.

### Study selection

As part of the living SRNMA, pairs of reviewers, following calibration exercises to ensure sufficient agreement, work independently and in duplicate to screen titles and abstracts of search records and subsequently the full texts of records determined potentially eligible at the title and abstract screening stage. Reviewers also link preprint reports with their subsequent publications based on trial registration numbers, the names of investigators, recruiting centres and countries, dates of recruitment, and other trial characteristics. When links between preprints and subsequent publications are unclear, we contact the author for confirmation. Reviewers resolve discrepancies by discussion or, when necessary, by adjudication with a third-party reviewer.

Eligible preprint and peer reviewed articles report trials that randomize patients with suspected, probable, or confirmed COVID-19 to drug treatments, antiviral antibodies and cellular therapies, placebo, or standard care or trials that randomize healthy participants exposed or unexposed to COVID-19 to prophylactic drugs, standard care, or placebo. We do not apply any restrictions on severity of illness, setting, or language of publication but do exclude trials that report on nutritional interventions, traditional Chinese herbal medicines without standardization in formulations and dosing across batches, and non-drug supportive care interventions.

We did not perform a sample size calculation since we included all eligible trial reports identified through our living SRNMAS up to August 3^rd^, 2021.

### Data collection

As part of the living SRNMA, for each eligible trial, pairs of reviewers, following training and calibration exercises, independently extract trial characteristics, methods, and results using a standardized, pilot tested data extraction form. To assess risk of bias, reviewers, following training and calibration exercises, use a revision of the Cochrane tool for assessing risk of bias in randomized trials (RoB 2.0) (22) (Supplement 3). Reviewers resolve discrepancies by discussion and, when necessary, by adjudication with a third party.

For the current study, pairs of trained and calibrated reviewers, working independently and in duplicate and using a pilot-tested form, collected data on differences in key methods and results between preprint and published trial reports. Key methods included description of the randomization process and allocation concealment, blinding of patients and healthcare providers, extent of and handling of missing outcome data, blinding of outcome assessors and adjudicators, and prespecification of outcomes and analyses. Key results included number of participants analyzed and number of events in each trial arm for dichotomous outcomes and number of participants analyzed, means or medians and measures of variability for continuous outcomes. We focused on the same outcomes as our living SRNMA: mortality, mechanical ventilation, adverse events leading to discontinuation, viral clearance, admission to hospital, viral clearance, hospital length of stay, ICU length of stay, duration of mechanical ventilation, time to symptom resolution or clinical improvement, time to viral clearance, days free from mechanical ventilation, and time to viral clearance. For preprints with more than one version, we extracted data from the first version of the preprint, which is the least likely to have been modified in response to peer review.

Because risk of bias may vary across outcomes, for this analysis we present risk of bias judgements corresponding to the following hierarchy of outcomes for therapy trials: mortality, mechanical ventilation, duration of hospitalization, time to symptom resolution or clinical improvement, and virologic outcomes. For prophylaxis trials, we used the following hierarchy: mortality, laboratory confirmed and suspected COVID-19 infection, and laboratory confirmed COVID-19 infection. These hierarchies represent the relative importance of outcomes based on rankings made by the linked WHO guideline panel (23).

### Data synthesis and analysis

#### Characteristics and risk of bias of trials

We compare the characteristics and risk of bias of trials with preprints, trials with publications, and trials first posted as a preprint and subsequently published by calculating differences in proportions and associated confidence intervals.

#### Time to publication of trial preprints

We calculated the median time from a trial being posted on a preprint server to its eventual publication in a journal and used Kaplan-Meier curves and log-rank tests to assess whether source of funding, number of centers and participants, early termination for benefit, intensity of care (inpatient versus outpatient), and severity (mild/moderate versus severe/critical COVID-19), statistically significant primary or secondary outcomes (based on cut-offs defined by the authors or, when no cut-offs were defined, based on a cut-off of p<0.05 or confidence intervals not including the null), and risk of bias (trials rated at low versus high risk of bias) are predictive of time to publication of trial preprints.

#### Differences between preprint and published trial reports

Among trial preprints that were subsequently published in a peer reviewed journal, we described the number and types of discrepancies in key methods and results between preprint and published trial reports. For discrepancies in the reporting of key methods, we report the number and percentage of the changes between preprints and publications that affected risk of bias judgements—changes that we considered to be critical.

#### Retractions

We compare the number of preprint and published trials that have been retracted.

#### Comparison of meta-analyses including versus excluding evidence from preprints

For trials that report on interventions that have been addressed by the linked WHO living guideline (23) up to August 3^rd^ 2021 (i.e., corticosteroids, remdesivir, lopinavir-ritonavir, hydroxychloroquine, ivermectin, IL-6 receptor blockers, and convalescent plasma for treatment and hydroxychloroquine for prophylaxis) and the two most commonly reported outcomes (i.e., mortality, mechanical ventilation), we conducted pairwise frequentist random-effects meta-analyses with the restricted maximum likelihood estimator including versus excluding evidence from preprints at one, three, and six months after the first trial addressing the drug of interest was made public, either via preprint or publication. We also conducted an analysis including versus excluding evidence from preprints at August 3^rd^ —the longest timepoint at which we collected data. For hydroxychloroquine for prophylaxis, because mechanical ventilation was not an outcome of interest for prophylaxis trials, we report only on mortality.

To facilitate interpretation, we calculated absolute effects. To calculate absolute effects, for drug treatments, we used mortality data from the CDC and data on ventilation from the International Severe Acute Respiratory and Emerging Infection COVID-19 database (24-26). For prophylaxis, we used the event rate among all participants randomized to standard care or placebo to calculate the baseline risk.

We compared the direction of effect between meta-analyses including versus excluding preprints. We considered the direction of effect different if one point estimate suggested no effect and another suggested a benefit or harm or if one point estimate suggested benefit and another suggested harm. For treatment, we considered an effect to be beneficial if the point estimate indicated a reduction in risk of mortality of 1% or greater or a reduction in risk of mechanical ventilation of 2% or greater for treatment. For prophylaxis, we considered an effect to be beneficial if the point estimate indicated a reduction in risk of mortality of 0.5% or greater. For treatment, we considered an effect to be harmful if the point estimate indicated that there was an increase in risk of mortality of 1% or greater or an increase in risk of mechanical ventilation of 2% or greater. For prophylaxis, we considered an effect to be harmful if the point estimate indicated an increase in risk of mortality of 0.5% or greater for prophylaxis. Otherwise, we inferred that there was no important effect.

The GRADE approach provided the framework for assessing certainty of evidence and informed whether including versus excluding preprint reports led to differences in ratings of the overall certainty of evidence, judgments related to specific GRADE domains, and whether differences in ratings are likely to impact decision making (i.e., evidence rated as high/moderate versus low/very low) (27). A minimally contextualized approach informed ratings of imprecision (28). We considered any effect on mortality and mechanical ventilation to be important. Thresholds of 1% risk difference for mortality and a 2% risk difference for mechanical ventilation informed judgements of minimal or no treatment effect. For prophylaxis and mortality, we used a 0.5% risk difference (28).

We performed all statistical analysis in R (version 4.03, R Foundation for Statistical Computing), using the meta, forestplot, survival, and survminer packages.

### Patient/Public Involvement

Patients were involved in outcome selection, interpretation of results, and the generation of parallel recommendations, as part of the parallel SRNMA and guidelines (23).

## Results

### Trial characteristics

As of August 3^rd^, we identified 356 eligible trials, 101 of which were only available as preprints, 181 only available as journal publications, and 74 first available as preprints and subsequently published as journal articles. Supplement 4 presents additional details on the results of the search and Table 1 presents overall trial characteristics overall and trial characteristics stratified by publication status.

**Table 1:** Trial characteristics.

| **Table 1: Trial characteristics** |  |  |  |  |  |  |  |  |
| --- | --- | --- | --- | --- | --- | --- | --- | --- |
|  | **All trials**<br>**n (%)**<br>**N=356** | **Published**<br>**trials**<br>**without**<br>**preprints**<br>**n (%)**<br>**N=181** | **Trials only**<br>**available as**<br>**preprints**<br>**n (%)**<br>**N=101** | **% difference**<br>**between trials only**<br>**available as**<br>**preprints and**<br>**published trials**<br>**without preprints**<br>**(95% Cis)** | **P**<br>**value** | **Trials first posted as**<br>**preprints and**<br>**subsequently**<br>**published**<br>**n (%)**<br>**N=74** | **% difference between trials first**<br>**posted as preprints and**<br>**subsequently published and**<br>**published trials without preprints**<br>**(95% Cis)** | **P**<br>**value** |
| **Trial registered** |  |  |  |  |  |  |  |  |
| Yes | 319 (89.6%) | 149 (82.3%) | 97 (96.0%) | 13.72% (7.0 to 20.5) | <0.001 | 73 (98.6%) | 16.3% (10.2 to 22.5) | <0.001 |
| No | 37 (10.4%) | 32 (17.7%) | 4 (4.0%) | -13.72% (-20.5 to -7.0) | <0.001 | 1 (1.4%) | -16.3% (-22.5 to -10.2) | <0.001 |
| **Study status** |  |  |  |  |  |  |  |  |
| Completed | 268 (75.3%) | 142 (78.5%) | 75 (74.3%) | -4.2% (-14.6 to 6.2) | 0.44 | 51 (68.9%) | -9.5% (-21.7 to 2.6) | 0.12 |
| Interim results | 20 (5.6%) | 5 (2.8%) | 10 (9.9%) | 7.1% (0.8 to 13.4) | 0.03 | 5 (6.8%) | 4.0% (-2.2 to 10.2) | 0.21 |
| Terminated early for benefit | 2 (0.6%) | 0 (0%) | 2 (2.0%) | 2.0% (-0.7 to 4.7) | 0.15 | 0 (0%) | 0 (0%) | 1.00 |
| Terminated early due to feasibility | 66 (18.5%) | 34 (18.8%) | 14 (13.9%) | -4.9% (-13.7 to 3.9) | 0.28 | 18 (24.3%) | 5.5% (-5.8 to 16.9) | 0.34 |
| **Type of treatment** |  |  |  |  |  |  |  |  |
| Drug therapy | 293 (82.3%) | 157 (86.7%) | 77 (76.2%) | -10.5% (-20.2 to -0.8) | 0.03 | 59 (79.7%) | -7.0% (-17.4 to 3.4) | 0.19 |
| Prophylaxis | 17 (4.8%) | 7 (3.9%) | 8 (7.9%) | 4.0% (-1.9 to 10.0) | 0.18 | 2 (2.7%) | -1.2% (-5.8 to 3.5) | 0.64 |
| Antiviral antibodies and cellular therapies | 46 (12.9%) | 17 (9.4%) | 16 (15.8%) | 6.5% (-1.8 to 14.7) | 0.13 | 13 (17.6%) | 8.2% (-1.5 to 17.8) | 0.10 |
| Inpatient† | 252 (74.3%) | 129 (72.3%) | 65 (69.9%) | -4.3% (-15.6 to 7.1) | 0.47 | 58 (80.6%) | 6.4% (-4.8 to 17.6) | 0.27 |
| Severe/critical disease† | 88 (26.0%) | 46 (26.4%) | 26 (28.0%) | 1.5% (-9.7 to 12.8) | 0.80 | 16 (22.2%) | -4.2% (-15.8 to 7.4) | 0.49 |
| **Number of centers** |  |  |  |  |  |  |  |  |
| Single center | 151 (42.4%) | 86 (47.5%) | 39 (38.6%) | -8.9% (-20.9 to 3.1) | 0.15 | 26 (35.1%) | -12.4% (-25.5 to 0.7) | 0.06 |
| Multicenter | 177 (49.7%) | 85 (47.0%) | 48 (47.5%) | 0.6% (-12.0 to 12.7) | 0.93 | 44 (59.5%) | 12.5% (-0.8 to 25.8) | 0.07 |
| Median [IQR] number of participants | 106 [60 to 266] | 101 [60 to 208] | 163 [64 to 412] |  |  | 100 [60 to 249] |  |  |
| Primary outcome statistically significant | 151 (42.4%) | 74 (40.9%) | 49 (48.5%) | 7.6% (-4.5 to 19.7) | 0.22 | 28 (37.8%) | -3.1% (-16.2 to 10.1) | 0.66 |
| Any secondary outcome(s) statistically significant | 186 (52.2%) | 90 (49.7%) | 53 (52.5%) | 2.8% (-9.4 to 14.9) | 0.67 | 43 (58.1%) | 8.4% (-5.0 to 21.8) | 0.22 |
| **Risk of bias** |  |  |  |  |  |  |  |  |
| High risk of bias due to randomization | 122 (34.3%) | 67 (37.0%) | 34 (33.7%) | -3.4% (-15.0 to 8.2) | 0.58 | 19 (25.7%) | -11.3% (-23.5 to 0.9) | 0.07 |
| High risk of bias due to deviations from intended intervention | 214 (60.1%) | 111 (61.3%) | 56 (55.5%) | -5.9% (-17.9 to 6.1) | 0.34 | 47 (63.5%) | 2.2% (-10.9 to 15.3) | 0.76 |
| High risk of bias due to missing outcome data | 19 (5.3%) | 11 (6.1%) | 5 (5.0%) | -1.1% (-6.6 to 4.4) | 0.70 | 3 (4.1%) | -2.0% (-7.7 to 3.7) | 0.50 |
| High risk of bias due to measurement of the outcome | 7 (2%) | 6 (3.3%) | 1 (1.0%) | -2.3% (-5.6 to 0.9) | 0.16 | 0 (0%) | -3.3% (-5.9 to -0.7) | 0.01 |
| High risk of bias due to selective reporting | 22 (6.2%) | 9 (5.0%) | 8 (7.9%) | 3.0% (-3.2 to 9.1) | 0.35 | 5 (6.8%) | 1.8% (-4.8 to 8.3) | 0.60 |
| High risk of bias overall | 228 (64%) | 117 (64.6%) | 61 (60.4%) | -4.2% (-16.1 to 7.6) | 0.49 | 50 (67.6%) | 2.9% (-9.8 to 15.7) | 0.67 |
| **Funding\*** |  |  |  |  |  |  |  |  |
| Industry | 79 (22.2%) | 32 (17.7%) | 30 (29.7%) | 12.0% (1.5 to 22.5) | 0.02 | 17 (23%) | 5.3% (-5.8 to 16.4) | 0.35 |
| Government | 126 (35.4%) | 47 (26.0%) | 37 (36.6%) | 10.7% (-0.7 to 22.0) | 0.07 | 42 (56.8%) | 30.8% (17.8 to 43.8) | <0.001 |
| Institution | 128 (36.0%) | 69 (38.1%) | 32 (31.7%) | -6.4% (-17.9 to 5.1) | 0.28 | 27 (36.5%) | -1.6% (-14.7 to 11.4) | 0.82 |
| Not-for-profit | 51 (14.3%) | 24 (13.3%) | 12 (11.9%) | -1.4% (-9.4 to 6.6) | 0.75 | 15 (20.3%) | 7.0% (-3.4 to 17.4) | 0.19 |
| Not reported | 23 (6.5%) | 12 (6.6%) | 10 (9.9%) | 3.3% (-3.6 to 10.1) | 0.36 | 1 (1.4%) | -5.3% (-9.8 to -0.8) | 0.02 |
| No funding | 49 (13.8%) | 33 (18.2%) | 10 (9.9%) | -8.3% (-16.4 to -0.2) | 0.04 | 6 (8.1%) | -10.1% (-18.5 to -1.7) | 0.02 |
| * Trials may be classified in more than one category. |  |  |  |  |  |  |  |  |
| † Estimates only include trials addressing therapy. |  |  |  |  |  |  |  |  |

Typical trials were registered, completed at the time of reporting, addressed drug therapies, enrolled less than 250 participants, reported one or more secondary outcomes that were statistically significant, and were funded by governments and/or institutions. Nearly two thirds of trials were at high risk of bias, primarily due to their open-label design.

Compared to published trials without preprints, trials only available as preprints and trials first available as preprints and then subsequently published were more likely to be registered; trials only available as preprints were more likely to report on interim results, describe drug therapies compared to antiviral antibodies and cellular therapies or prophylaxis, and to have received industry funding; and trials first posted as preprints and subsequently published were more likely to have received government funding.

### Predictors of publication and time to publication

During the 1.5 year span of this study, of 175 preprints, 74 (42.3%) were subsequently published in peer-reviewed journals. Table 2 presents the proportion of preprints published up to one year and table 3 presents predictors of publication. A quarter of preprints were published in peer reviewed journals by 3 months after the preprint first became available and half by 6 months. At one year, a third of preprints remained unpublished.

**Table 2:**
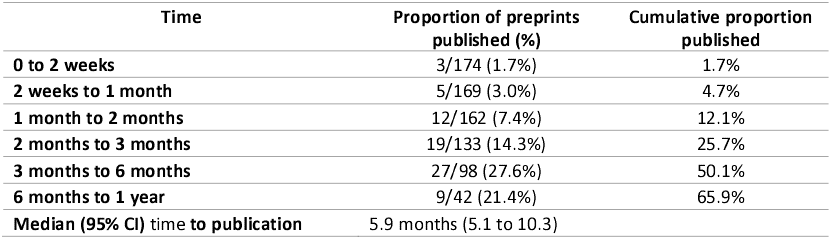
Time to publication of COVID-19 trial preprints.

**Table 3:** Predictors of time to publication of COVID-19 trial preprints.

| Table 3: Predictors of time to publication of COVID-19 trial preprints |  |  |
| --- | --- | --- |
|  | Median days (95% Cis) | P value |
| Funding |  |  |
| Industry funding (n=47) | 171 (145 to NA) | 0.72 |
| No industry funding (n=127) | 219 (177 to NA) |  |
| Government funding (n=78) | 155 (120 to 192) | 0.02 |
| No government funding (n=96) | 309 (183 to NA) |  |
| Institutional funding (n=58) | 136 (90 to NA) | 0.14 |
| No institutional funding (n=116) | 192 (166 to NA) |  |
| Not-for-profit funding (n=26) | 110 (79 to NA) | 0.07 |
| No not-for-profit funding (n=148) | 188 (161 to NA) |  |
| Number of centers |  |  |
| Single center (n=65) | undefined (130 to undefined) | 0.13 |
| Multicenter (n=90) | 161 (136 to 219) |  |
| Number of participants |  |  |
| More than median (n=88) | 142 (120 to NA) | 0.14 |
| Less than median (n=85) | 219 (171 to NA) |  |
| Intensity of care |  |  |
| Inpatient (n=122) | 110 (95 to 1440) | <0.0001 |
| Outpatient (n=38) | undefined |  |
| Severity |  |  |
| Mild/moderate (79) | 182 (159 to 263) | <0.0001 |
| Severe/Critical (42) | 101 (89 to 128) |  |
| Early termination for benefit | NA | NA |
| Primary outcome statistically significant |  |  |
| Statistically significant (n=77) | 243 (155 to undefined) | 0.12 |
| Not statistically significant (n=91) | 161 (127 to 219) |  |
| Any secondary outcomes statistically significant |  |  |
| Statistically significant (n=96) | 187 (120 to NA) | 0.66 |
| Not statistically significant (n=64) | 152 (120 to NA) |  |
| Risk of bias |  |  |
| Low (n=52) | 155 (124 to 187) | 0.17 |
| High (n=102) | 122 (98 to 161) |  |

Preprints that received government funding, reported on inpatients, or reported on patients with severe disease were published faster than preprints that did not receive government funding, reported on outpatients, or patients with mild/moderate disease, respectively.

### Differences between preprint and published trial reports

Forty-two trials (56.8%) had one or more discrepancies in the reporting of key methods and results between the preprint and the later published trial report. Supplement 5 describes these discrepancies.

Thirty trials (40.5%) had one or more discrepancies in the reporting of key methods--all of which had to do with information either missing from the preprint or the publication. The most common discrepancy in the reporting of key methods was the description of allocation concealment, which occurred in eight trials. Box 1 presents an example. Our judgement of risk of bias for the randomization domain changed from ‘probably high risk of bias’ to ‘low risk of bias’ for four of these eight trials due to additional details reported in the published report.

**Box 1: Example of a trial that reported additional information on allocation concealment in the published report**

The PANAMO trial, which was initially available as a preprint on SSRN and later published in *Lancet Rheumatology*, provided additional details in the publication on allocation concealment. The publication describes central randomization with an online tool and the development of the randomization list by a third party—all of which were not reported in the preprint (29, 30).

**Preprint**

“Patients were randomly assigned in a 1:1 ratio to receive IFX-1, at a dose of 800 mg intravenously, for a maximum of seven doses, plus best supportive care, or best supportive care only. [..] Randomization was performed with an online tool within the eCRF [electronic case report form] and was stratified by study site.”

**Publication**

“Patients were randomly assigned in a 1:1 ratio to IFX-1 plus best supportive care (the IFX-1 group) or to best supportive care only (the control group). Randomisation was done by investigators centrally with an online tool within the electronic case report form and was stratified by study site. The tool used a randomised variable block length of either 2 or 4. The randomisation list was only available to contract research organisation (Metronomia) staff involved in the production of the randomisation list and set-up of the online randomisation tool.”

Other differences in the reporting of key methods were the publication reporting one or more additional statistics important for meta-analysis (e.g., IQR or SD) that were not previously reported in the preprint (n=6; 8.1%), the preprint reporting on interim results and the publication on completed trial results (n=4; 5.4%), and the publication including a protocol and/or statistical analysis plan as a supplementary that was not previously included with the preprint (n=3; 4.1%). The overall trial rating of risk of bias, however, changed only for one trial based on additional information provided in the published report.

Thirty-one trials (41.9%) had one or more differences in the reporting of key results between preprints and publications. The most common discrepancy in the reporting of key results were changes in outcome data between preprints and publications, which was seen in 20 trials (27.0%)—though most of these discrepancies may be attributed to events accumulating in trials from the time at which the preprint was posted to when the trial was published. Box 2 presents an example.

**Box 2: Example of a trial that reported different outcome data between the preprint and publication**

The RECOVERY publication on dexamethasone, for example, reported different results for mortality and mechanical ventilation—likely not because of an error in the preprint but because of events accumulating in the trial from when the preprint was posted to when it was published (2, 31).

| Outcome | Preprint (31) | Publication (2) |
| --- | --- | --- |
| Mortality | Corticosteroids: 454/2104 (21.6 %)<br>Standard care: 1065/4321 (24.6%) | Corticosteroids: 482/2104 (22.9%)<br>Standard care: 1110/4321 (25.7%) |
| Mechanical ventilation | Corticosteroids: 92/1780 (5.2%)<br>Standard care: 258/3638 (7.1%) | Corticosteroids: 110/1780 (6.2%)<br>Standard care: 298/3638 (8.2%) |

Despite discrepancies in outcome data being common, the magnitude and precision of effects were similar between preprints and publications. Figure 1 shows differences in results on mortality and mechanical ventilation between preprints and publications. Among all preprints with differences in outcomes, differences in relative effects did not exceed 15%, except for one trial with very few events that included just one additional event in the publication (32, 33). Other differences between preprints and publications in key results included the publication reporting an outcome that was not included in the preprint (n=11; 14.9%).

**Figure 1:**
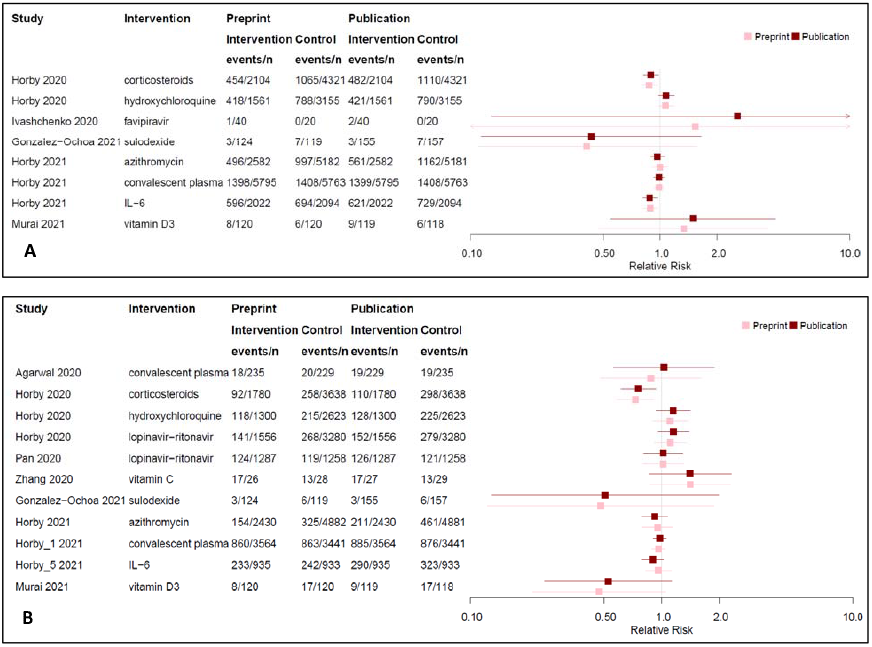
Differences in results reported between preprints and publications for mortality (A) and mechanical ventilation (B)

### Retractions

We identified four retracted trials (34-41). Two trials reported on (hydroxy)chloroquine (34, 35)(36, 37), two on favipiravir (36, 37)(38, 39), and two on ivermectin (34, 35)(40, 41). One of the four trials was single-centre (40, 41) and the remainder were multi-centre trials. One of the trials was retracted when the authors noticed an error in their analysis (40, 41) and the remainder were retracted due to concerns about data fabrication or falsification (e.g., inconsistencies between the eligibility criteria and patients included in the trial, discrepancies between when the trial was reported to have been conducted and when patients were recruited, inconsistencies between the dataset and the results reported in the preprint, inconsistencies between the distribution of baseline variables and the described randomization procedure). We compared the number of retractions between preprints and journal publications. One of the retracted trials was posted as a preprint (34, 35) and the remainder were published in peer-reviewed journals.

### Evidence synthesis including versus excluding preprint reports

Table 4 presents results of meta-analyses including and excluding data from unpublished preprints for the comparison of placebo or standard care with corticosteroids, remdesivir, lopinavir-ritonavir, hydroxychloroquine, ivermectin, IL-6 receptor blockers, and convalescent plasma and mortality and mechanical ventilation at 1 month, 3 months, and 6 months after the first trial addressing the intervention was made public, either as a preprint or a publication, and at the longest point of follow-up of the trials (August 3^rd^). In total, we performed and assessed the certainty of evidence of 120 meta-analyses, 60 of which included preprints and 60 of which excluded preprints. Supplement 6 presents forest plots for meta-analyses.

Because of insufficient data, we could not perform meta-analyses for six comparisons without preprints: mortality and mechanical ventilation, at one month, for ivermectin vs. placebo/standard care and IL-6 receptor blockers vs. placebo/standard care and mortality and mechanical ventilation, at three months, for ivermectin vs. placebo/standard care.

### Differences in estimates from meta-analyses including versus excluding preprints

Except for two cases, across all meta-analyses including and excluding results from unpublished preprints, the point estimates were consistent as to whether they indicated benefit, no appreciable effect, or harm. The meta-analysis of corticosteroids at one month suggested a reduction in risk of mechanical ventilation when preprints were excluded (43 fewer per 1,000 people [95% CI 59.24 fewer to 22.12 fewer]; moderate certainty) and no appreciable effect when preprints were included (1.2 more per 1,000 people [95% CI 60.3 fewer to 131.1 more]; very low certainty). The meta-analysis of ivermectin at 6 months suggested no appreciable effect on risk of mechanical ventilation when preprints were excluded (2.3 fewer per 1,000 people [95% CI 52.2 fewer to 83.5 more]; low certainty) and a reduction in risk of mechanical ventilation (26.7 fewer per 1,000 people [95% CI 74.2 fewer to 75.4 more]; very low certainty) when preprints were included.

### Differences in ratings of the certainty of evidence from meta-analyses including versus excluding preprints

There were nine cases for which the rating of the certainty of evidence was different when preprints were included versus excluded. For four of these nine cases, we rated the certainty of evidence including preprints higher than the evidence excluding preprints due to no serious concerns in imprecision in analyses including preprints. For five of these nine cases, we rated the certainty of evidence excluding preprints higher than the evidence including preprints, due to serious imprecision for four of five cases and due to serious imprecision and serious risk of bias for one of five cases. In six of these cases, differences in ratings of the certainty of evidence may have impacted decision-making (i.e., evidence including preprints is rated as high/moderate whereas evidence excluding preprints is rated as low/very low or vice versa).

### Differences in ratings of the GRADE risk of bias domain from meta-analyses including versus excluding preprints

Between meta-analyses including versus excluding preprints, judgements related to the GRADE risk of bias domain differed only for the meta-analysis of remdesivir and mechanical ventilation at 6 months. We rated the meta-analysis excluding preprints to not have any concerns related to risk of bias and downgraded the meta-analysis including preprints due to serious risk of bias.

### Differences in ratings of the GRADE imprecision domain from meta-analyses including versus excluding preprints

Between meta-analyses including versus excluding preprints, judgements related to the GRADE imprecision domain differed for 13 meta-analyses. We judged nine meta-analyses excluding preprints to have more serious concerns related to imprecision than their counterparts including preprints (i.e., additional data from preprints narrowed confidence intervals) and we judged four meta-analyses excluding preprints to have less serious concerns related to imprecision that their counterparts including preprints (i.e., additional data from preprints increased statistical heterogeneity and hence imprecision).

## Discussion

### Main findings

Our study presents a detailed assessment of the trustworthiness and impact of COVID-19 trial preprint reports. We show that preprints remain the only source of findings of many trials for several months. Half of all preprints, for example, remain unpublished at six months and a third at one year—a length of time that may be unacceptable in a health emergency like COVID-19. Preprints can importantly accelerate the time to dissemination of trial findings.

We did not find compelling evidence of important differences between preprint and published reports of trials—though preprint reports of trials that are subsequently published in journals may not be representative of all trial preprints. Further, we found retractions to occur for both preprints and publications, suggesting that publication in a peer-reviewed journal alone does not indicate trustworthiness of a trial report.

We also found that in most cases, meta-analyses including versus excluding evidence from preprints yielded consistent results and the same certainty of evidence. In a minority of circumstances, however, including preprints improved the certainty of evidence. At six months after trial data first became available, when preprints were excluded, we found low certainty evidence that IL-6 receptor blockers may reduce mortality—downgraded due to risk of bias and imprecision. Conversely, we found moderate certainty evidence IL-6 receptor blockers probably reduce mortality when preprints were included— downgraded due to risk of bias. For the case of IL-6 receptor blockers, considering evidence from preprints may have importantly accelerated the time to incorporation of IL-6 receptor blockers as part of standard care (23). In a minority of circumstances, including preprints also reduced the certainty of evidence. At one month, for example, when preprints were excluded, we found moderate certainty evidence that corticosteroids probably reduce mechanical ventilation—downgraded due to serious risk of bias—and very low certainty evidence when we included preprints—downgraded due to serious risk of bias and very serious imprecision. For the case of corticosteroids, including preprints increased heterogeneity (and hence increased imprecision), and may have misinformed evidence users.

### Implications

Our findings have implications for evidence users who are concerned with the trustworthiness of preprints and for systematic reviewers and guideline developers deciding whether to consider preprint reports in systematic reviews and guideline recommendations. Our results support the overall trustworthiness of preprints and preprints as a venue through which the dissemination of trial findings may be accelerated. Peer review likely only addresses transparency of the trial reports and interpretation of results because at the time of submission the trial has already been conducted and major methodological decisions (such as whether to collect data on an outcome or whether to blind investigators) cannot be changed. We encourage systematic reviewers and guideline developers to consider trial evidence from preprints, especially in circumstances in which decisions are being made rapidly and evidence is being produced faster than can be peer reviewed and published.

We caution, however, that preprints (and publications) may describe untrustworthy trials with fabricated or falsified data. Evidence users may consider scrutinizing both preprint and published trials for anomalies that may suggest fabrication and falsification (Examples of which are reported in Box 3) (42, 43). While such methods cannot be used to definitively identify untrustworthy trials and require subjective judgements, they may be useful to identify trials that are at high risk of such issues. Evidence users may subsequently investigate such trials further or systematic reviewers may consider sensitivity analyses excluding such trials from meta-analyses. We direct readers to other sources that describe these methods (44).

**Box 3: Methods to assess for fabrication and falsification in clinical trials (44)**

1. Judge whether the reported recruitment speed is feasible given local disease patterns, trial eligibility criteria, and capacity of the recruiting centres (45, 46).
2. Review the trial registration or protocol and assess the consistency between the registration and the manuscript in aspects of the trial that cannot be modified after completion (e.g., blinding status).
3. Review profiles of investigators and/or institutions involved for a history of research misconduct.
4. Review baseline patient characteristics and test whether reported distributions are consistent with randomization (42, 43, 47).
5. Review primary data, when available, for duplicate records or inconsistencies between the data and reported statistics in the trial manuscript.
6. Review primary data, when available, and assess whether correlations between variables are plausible (45).

### Relation to previous work

Our study is the first to present data addressing the contribution of preprints to the body of evidence addressing the comparative effectiveness of COVID-19 therapies and prophylaxis.

Two studies have reported on differences between preprint and published study reports and citations and Altmetric attention metrics (48, 49). One study addressed publication characteristics and dissemination of COVID-19 preprints and the other spin in interpretation of results. Both studies were, however, restricted to only publications up to August and October 2020—which is not representative of the current landscape of COVID-19 research and which does not include the majority of evidence being currently used to guide COVID-19 care, including critical trials addressing the effects of corticosteroids and IL-6 receptor blockers (1, 2). These studies did not compare the effects of including preprints on meta-analytic estimates and the certainty of the body of evidence, which is particularly important because evidence users use the totality of the body of evidence, rather than single studies, to make treatment decisions and recommendations (48).

The COVID-19 pandemic highlighted the need for rapid dissemination of research and incited increased interest in preprint servers, which yielded to an incredible amount of research that was published on preprint servers and which made this study possible. We are not aware of studies addressing the trustworthiness or impact of preprints in other areas—though such research in other areas would also be useful.

### Strengths and limitations

The strengths of this study include the comprehensive search for, and inclusion of preprint and published COVID-19 trial reports and rigorous data collection. The generalizability of our results is, however, limited to COVID-19. Journals have expedited the publication of COVID-19 research and have been publishing more prolifically on COVID-19 than in other areas, which may reduce opportunity for revisions between preprints and their subsequent publications and may mean time to and predictors of publication may be different than in other research areas.

Although the WHO COVID-19 database is a comprehensive source of published and preprint literature, it does not include all preprint servers—though preprint servers not covered by our search address other subjects and are unlikely to include COVID-19 trials.

It is likely that preprint reports of trials that are subsequently published in journals represent the most rigorous or transparently reported preprints and that they are not representative of all trial preprints.

To assess preprint trustworthiness, we compared reporting of key aspects of the methods and results between preprint and published trial reports. We acknowledge, however, that published trial reports may still contain errors and that posting trial reports as preprints may allow more errors to be identified prior to final publication.

We report on the number of publications and preprints that were retracted. Preprints, however, may be less likely to be retracted because they may draw less attention and because preprint servers may be less likely than journals to have formal policies addressing research integrity.

Our assessment of the contribution of trial preprint reports to meta-analytic estimates and their effect on the certainty of evidence was undertaken in the context of pairwise meta-analysis and the minimally contextualized approach for assessing the certainty of evidence whereas parallel guideline recommendations have been based on network meta-analyses and the fully contextualized approach (23, 28). There are no compelling reasons that our results will not be generalizable to network meta-analyses. On the other hand, there will be more differences in judgements related to the certainty of evidence in a fully contextualized framework where judgements are more dependent on the magnitude and precision of estimates.

We limited our assessment of the impact of including versus excluding preprint reports on meta-analytic estimates to only interventions that have been addressed by the WHO living guideline at the time of analysis (23). While the effects of including or excluding preprints in meta-analyses may vary across interventions, our analysis addresses the interventions for which there has been sufficient interest and research to instigate guideline recommendations. Interventions informed by fewer trials may be more sensitive to including or excluding evidence from preprints.

Our estimate of the time to publication of preprint reports may be overestimated if some preprint authors do not attempt to subsequently publish in peer-reviewed journals—although evidence shows that most authors of COVID-19 preprints intend to publish their findings (48). Time to publication may also be underestimated if preprints are made public later in the submission process.

Finally, although we describe discrepancies in the reporting of key methods and results, we did not assess differences in the discussion or conclusion sections of trial reports.

## Conclusions

We found no compelling evidence that preprint trial reports provide less trustworthy results than published trials. We show that including preprints may affect the results of meta-analyses and the certainty of evidence and encourage evidence users to consider data from preprints in contexts in which decisions are being made rapidly and evidence is being produced faster than can be peer reviewed and published. Skepticism may still be warranted when suspicion arises regarding falsified data (for which we provide criteria).

## Data Availability

Upon a reasonable request to the corresponding author.

https://osf.io/9adxb/.

## Acknowledgements

None.

---

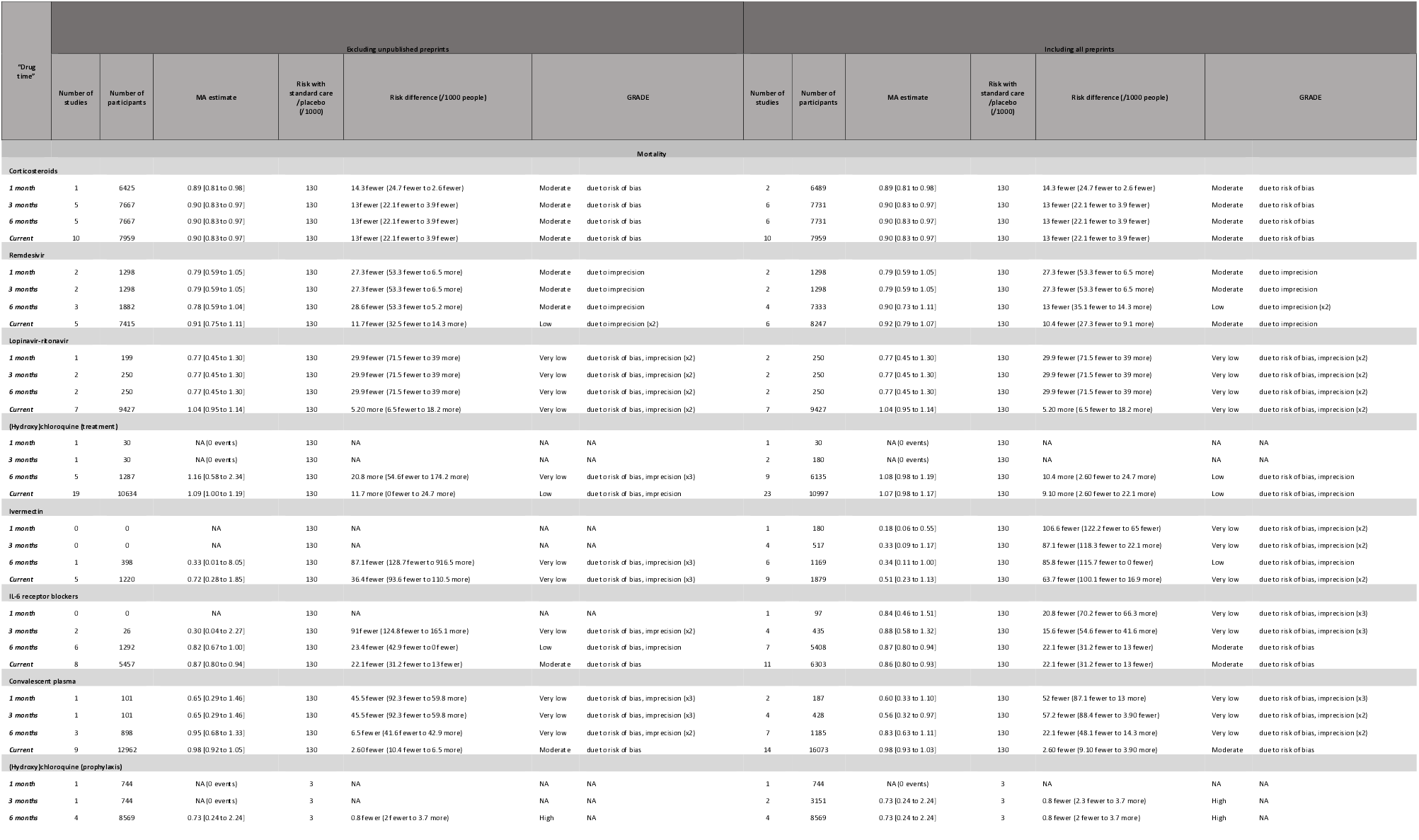
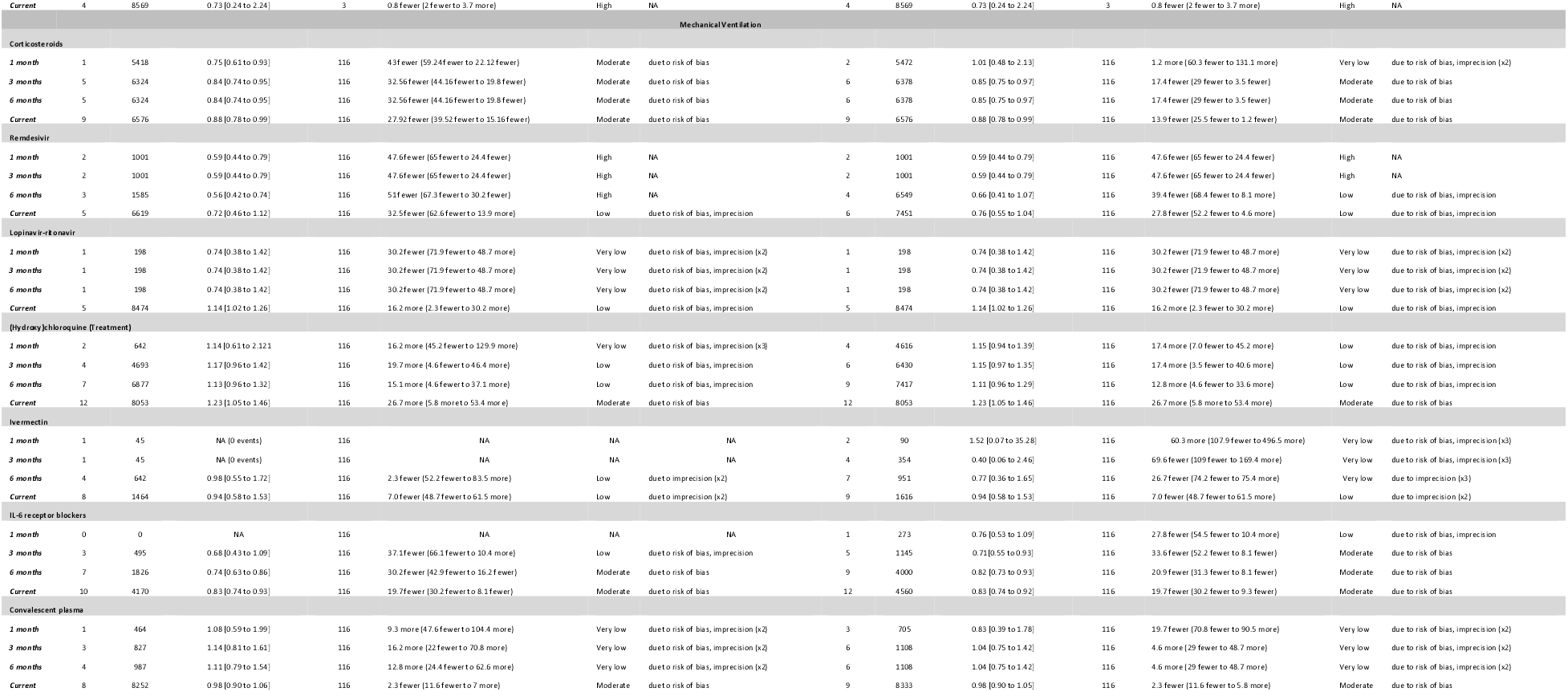

| “Drug time” | Excluding unpublished preprints |  |  |  |  |  |  | Including all preprints |  |  |  |  |  |  |
| --- | --- | --- | --- | --- | --- | --- | --- | --- | --- | --- | --- | --- | --- | --- |
|  | Number of studies | Number of participants | MA estimate | Risk with standard care /placebo (/1000) | Risk difference (/1000 people) | GRADE |  | Number of studies | Number of participants | MA estimate | Risk with standard care /placebo (/1000) | Risk difference (/1000 people) | GRADE |  |
| Mortality |  |  |  |  |  |  |  |  |  |  |  |  |  |  |
| Corticosteroids |  |  |  |  |  |  |  |  |  |  |  |  |  |  |
| 1 month | 1 | 6425 | 0.89 [0.81 to 0.98] | 130 | 14.3 fewer (24.7 fewer to 2.6 fewer) | Moderate | due to risk of bias | 2 | 6489 | 0.89 [0.81 to 0.98] | 130 | 14.3 fewer (24.7 fewer to 2.6 fewer) | Moderate | due to risk of bias |
| 3 months | 5 | 7667 | 0.90 [0.83 to 0.97] | 130 | 13 fewer (22.1 fewer to 3.9 fewer) | Moderate | due to risk of bias | 6 | 7731 | 0.90 [0.83 to 0.97] | 130 | 13 fewer (22.1 fewer to 3.9 fewer) | Moderate | due to risk of bias |
| 6 months | 5 | 7667 | 0.90 [0.83 to 0.97] | 130 | 13 fewer (22.1 fewer to 3.9 fewer) | Moderate | due to risk of bias | 6 | 7731 | 0.90 [0.83 to 0.97] | 130 | 13 fewer (22.1 fewer to 3.9 fewer) | Moderate | due to risk of bias |
| Current | 10 | 7959 | 0.90 [0.83 to 0.97] | 130 | 13 fewer (22.1 fewer to 3.9 fewer) | Moderate | due to risk of bias | 10 | 7959 | 0.90 [0.83 to 0.97] | 130 | 13 fewer (22.1 fewer to 3.9 fewer) | Moderate | due to risk of bias |
| Remdesivir |  |  |  |  |  |  |  |  |  |  |  |  |  |  |
| 1 month | 2 | 1298 | 0.79 [0.59 to 1.05] | 130 | 27.3 fewer (53.3 fewer to 6.5 more) | Moderate | due to imprecision | 2 | 1298 | 0.79 [0.59 to 1.05] | 130 | 27.3 fewer (53.3 fewer to 6.5 more) | Moderate | due to imprecision |
| 3 months | 2 | 1298 | 0.79 [0.59 to 1.05] | 130 | 27.3 fewer (53.3 fewer to 6.5 more) | Moderate | due to imprecision | 2 | 1298 | 0.79 [0.59 to 1.05] | 130 | 27.3 fewer (53.3 fewer to 6.5 more) | Moderate | due to imprecision |
| 6 months | 3 | 1882 | 0.78 [0.59 to 1.04] | 130 | 28.6 fewer (53.3 fewer to 5.2 more) | Moderate | due to imprecision | 4 | 7333 | 0.90 [0.73 to 1.11] | 130 | 13 fewer (35.1 fewer to 14.3 more) | Low | due to imprecision (x2) |
| Current | 5 | 7415 | 0.91 [0.75 to 1.11] | 130 | 11.7 fewer (32.5 fewer to 14.3 more) | Low | due to imprecision (x2) | 6 | 8247 | 0.92 [0.79 to 1.07] | 130 | 10.4 fewer (27.3 fewer to 9.1 more) | Moderate | due to imprecision |
| Lopinavir-ritonavir |  |  |  |  |  |  |  |  |  |  |  |  |  |  |
| 1 month | 1 | 199 | 0.77 [0.45 to 1.30] | 130 | 29.9 fewer (71.5 fewer to 39 more) | Very low | due to risk of bias, imprecision (x2) | 2 | 250 | 0.77 [0.45 to 1.30] | 130 | 29.9 fewer (71.5 fewer to 39 more) | Very low | due to risk of bias, imprecision (x2) |
| 3 months | 2 | 250 | 0.77 [0.45 to 1.30] | 130 | 29.9 fewer (71.5 fewer to 39 more) | Very low | due to risk of bias, imprecision (x2) | 2 | 250 | 0.77 [0.45 to 1.30] | 130 | 29.9 fewer (71.5 fewer to 39 more) | Very low | due to risk of bias, imprecision (x2) |
| 6 months | 2 | 250 | 0.77 [0.45 to 1.30] | 130 | 29.9 fewer (71.5 fewer to 39 more) | Very low | due to risk of bias, imprecision (x2) | 2 | 250 | 0.77 [0.45 to 1.30] | 130 | 29.9 fewer (71.5 fewer to 39 more) | Very low | due to risk of bias, imprecision (x2) |
| Current | 7 | 9427 | 1.04 [0.95 to 1.14] | 130 | 5.20 more (6.5 fewer to 18.2 more) | Very low | due to risk of bias, imprecision (x2) | 7 | 9427 | 1.04 [0.95 to 1.14] | 130 | 5.20 more (6.5 fewer to 18.2 more) | Very low | due to risk of bias, imprecision (x2) |
| (Hydroxy)chloroquine (treatment) |  |  |  |  |  |  |  |  |  |  |  |  |  |  |
| 1 month | 1 | 30 | NA (0 events) | 130 | NA | NA | NA | 1 | 30 | NA (0 events) | 130 | NA | NA | NA |
| 3 months | 1 | 30 | NA (0 events) | 130 | NA | NA | NA | 2 | 180 | NA (0 events) | 130 | NA | NA | NA |
| 6 months | 5 | 1287 | 1.16 [0.58 to 2.34] | 130 | 20.8 more (54.6 fewer to 174.2 more) | Very low | due to risk of bias, imprecision (x3) | 9 | 6135 | 1.08 [0.98 to 1.19] | 130 | 10.4 more (2.60 fewer to 24.7 more) | Low | due to risk of bias, imprecision |
| Current | 19 | 10634 | 1.09 [1.00 to 1.19] | 130 | 11.7 more (0 fewer to 24.7 more) | Low | due to risk of bias, imprecision | 23 | 10997 | 1.07 [0.98 to 1.17] | 130 | 9.10 more (2.60 fewer to 22.1 more) | Low | due to risk of bias, imprecision |
| Ivermectin |  |  |  |  |  |  |  |  |  |  |  |  |  |  |
| 1 month | 0 | 0 | NA | 130 | NA | NA | NA | 1 | 180 | 0.18 [0.06 to 0.55] | 130 | 106.6 fewer (122.2 fewer to 65 fewer) | Very low | due to risk of bias, imprecision (x2) |
| 3 months | 0 | 0 | NA | 130 | NA | NA | NA | 4 | 517 | 0.33 [0.09 to 1.17] | 130 | 87.1 fewer (118.3 fewer to 22.1 more) | Very low | due to risk of bias, imprecision (x2) |
| 6 months | 1 | 398 | 0.33 [0.01 to 8.05] | 130 | 87.1 fewer (128.7 fewer to 916.5 more) | Very low | due to risk of bias, imprecision (x3) | 6 | 1169 | 0.34 [0.11 to 1.00] | 130 | 85.8 fewer (115.7 fewer to 0 fewer) | Low | due to risk of bias, imprecision |
| Current | 5 | 1220 | 0.72 [0.28 to 1.85] | 130 | 36.4 fewer (93.6 fewer to 110.5 more) | Very low | due to risk of bias, imprecision (x3) | 9 | 1879 | 0.51 [0.23 to 1.13] | 130 | 63.7 fewer (100.1 fewer to 16.9 more) | Very low | due to risk of bias, imprecision (x2) |
| IL-6 receptor blockers |  |  |  |  |  |  |  |  |  |  |  |  |  |  |
| 1 month | 0 | 0 | NA | 130 | NA | NA | NA | 1 | 97 | 0.84 [0.46 to 1.51] | 130 | 20.8 fewer (70.2 fewer to 66.3 more) | Very low | due to risk of bias, imprecision (x3) |
| 3 months | 2 | 26 | 0.30 [0.04 to 2.27] | 130 | 91 fewer (124.8 fewer to 165.1 more) | Very low | due to risk of bias, imprecision (x2) | 4 | 435 | 0.88 [0.58 to 1.32] | 130 | 15.6 fewer (54.6 fewer to 41.6 more) | Very low | due to risk of bias, imprecision (x3) |
| 6 months | 6 | 1292 | 0.82 [0.67 to 1.00] | 130 | 23.4 fewer (42.9 fewer to 0 fewer) | Low | due to risk of bias, imprecision | 7 | 5408 | 0.87 [0.80 to 0.94] | 130 | 22.1 fewer (31.2 fewer to 13 fewer) | Moderate | due to risk of bias |
| Current | 8 | 5457 | 0.87 [0.80 to 0.94] | 130 | 22.1 fewer (31.2 fewer to 13 fewer) | Moderate | due to risk of bias | 11 | 6303 | 0.86 [0.80 to 0.93] | 130 | 22.1 fewer (31.2 fewer to 13 fewer) | Moderate | due to risk of bias |
| Convalescent plasma |  |  |  |  |  |  |  |  |  |  |  |  |  |  |
| 1 month | 1 | 101 | 0.65 [0.29 to 1.46] | 130 | 45.5 fewer (92.3 fewer to 59.8 more) | Very low | due to risk of bias, imprecision (x3) | 2 | 187 | 0.60 [0.33 to 1.10] | 130 | 52 fewer (87.1 fewer to 13 more) | Very low | due to risk of bias, imprecision (x3) |
| 3 months | 1 | 101 | 0.65 [0.29 to 1.46] | 130 | 45.5 fewer (92.3 fewer to 59.8 more) | Very low | due to risk of bias, imprecision (x3) | 4 | 428 | 0.56 [0.32 to 0.97] | 130 | 57.2 fewer (88.4 fewer to 3.90 fewer) | Very low | due to risk of bias, imprecision (x2) |
| 6 months | 3 | 898 | 0.95 [0.68 to 1.33] | 130 | 6.5 fewer (41.6 fewer to 42.9 more) | Very low | due to risk of bias, imprecision (x2) | 7 | 1185 | 0.83 [0.63 to 1.11] | 130 | 22.1 fewer (48.1 fewer to 14.3 more) | Very low | due to risk of bias, imprecision (x2) |
| Current | 9 | 12962 | 0.98 [0.92 to 1.05] | 130 | 2.60 fewer (10.4 fewer to 6.5 more) | Moderate | due to risk of bias | 14 | 16073 | 0.98 [0.93 to 1.03] | 130 | 2.60 fewer (9.10 fewer to 3.90 more) | Moderate | due to risk of bias |
| (Hydroxy)chloroquine (prophylaxis) |  |  |  |  |  |  |  |  |  |  |  |  |  |  |
| 1 month | 1 | 744 | NA (0 events) | 3 | NA | NA | NA | 1 | 744 | NA (0 events) | 3 | NA | NA | NA |
| 3 months | 1 | 744 | NA (0 events) | 3 | NA | NA | NA | 2 | 3151 | 0.73 [0.24 to 2.24] | 3 | 0.8 fewer (2.3 fewer to 3.7 more) | High | NA |
| 6 months | 4 | 8569 | 0.73 [0.24 to 2.24] | 3 | 0.8 fewer (2 fewer to 3.7 more) | High | NA | 4 | 8569 | 0.73 [0.24 to 2.24] | 3 | 0.8 fewer (2 fewer to 3.7 more) | High | NA |
| Current | 4 | 8569 | 0.73 [0.24 to 2.24] | 3 | 0.8 fewer (2 fewer to 3.7 more) | High | NA | 4 | 8569 | 0.73 [0.24 to 2.24] | 3 | 0.8 fewer (2 fewer to 3.7 more) | High | NA |
| Mechanical Ventilation |  |  |  |  |  |  |  |  |  |  |  |  |  |  |
| Corticosteroids |  |  |  |  |  |  |  |  |  |  |  |  |  |  |
| 1 month | 1 | 5418 | 0.75 [0.61 to 0.93] | 116 | 43 fewer (59.24 fewer to 22.12 fewer) | Moderate | due to risk of bias | 2 | 5472 | 1.01 [0.48 to 2.13] | 116 | 1.2 more (60.3 fewer to 131.1 more) | Very low | due to risk of bias, imprecision (x2) |
| 3 months | 5 | 6324 | 0.84 [0.74 to 0.95] | 116 | 32.56 fewer (44.16 fewer to 19.8 fewer) | Moderate | due to risk of bias | 6 | 6378 | 0.85 [0.75 to 0.97] | 116 | 17.4 fewer (29 fewer to 3.5 fewer) | Moderate | due to risk of bias |
| 6 months | 5 | 6324 | 0.84 [0.74 to 0.95] | 116 | 32.56 fewer (44.16 fewer to 19.8 fewer) | Moderate | due to risk of bias | 6 | 6378 | 0.85 [0.75 to 0.97] | 116 | 17.4 fewer (29 fewer to 3.5 fewer) | Moderate | due to risk of bias |
| Current | 9 | 6576 | 0.88 [0.78 to 0.99] | 116 | 27.92 fewer (39.52 fewer to 15.16 fewer) | Moderate | due to risk of bias | 9 | 6576 | 0.88 [0.78 to 0.99] | 116 | 13.9 fewer (25.5 fewer to 1.2 fewer) | Moderate | due to risk of bias |
| Remdesivir |  |  |  |  |  |  |  |  |  |  |  |  |  |  |
| 1 month | 2 | 1001 | 0.59 [0.44 to 0.79] | 116 | 47.6 fewer (65 fewer to 24.4 fewer) | High | NA | 2 | 1001 | 0.59 [0.44 to 0.79] | 116 | 47.6 fewer (65 fewer to 24.4 fewer) | High | NA |
| 3 months | 2 | 1001 | 0.59 [0.44 to 0.79] | 116 | 47.6 fewer (65 fewer to 24.4 fewer) | High | NA | 2 | 1001 | 0.59 [0.44 to 0.79] | 116 | 47.6 fewer (65 fewer to 24.4 fewer) | High | NA |
| 6 months | 3 | 1585 | 0.56 [0.42 to 0.74] | 116 | 51 fewer (67.3 fewer to 30.2 fewer) | High | NA | 4 | 6549 | 0.66 [0.41 to 1.07] | 116 | 39.4 fewer (68.4 fewer to 8.1 more) | Low | due to risk of bias, imprecision |
| Current | 5 | 6619 | 0.72 [0.46 to 1.12] | 116 | 32.5 fewer (62.6 fewer to 13.9 more) | Low | due to risk of bias, imprecision | 6 | 7451 | 0.76 [0.55 to 1.04] | 116 | 27.8 fewer (52.2 fewer to 4.6 more) | Low | due to risk of bias, imprecision |
| Lopinavir-ritonavir |  |  |  |  |  |  |  |  |  |  |  |  |  |  |
| 1 month | 1 | 198 | 0.74 [0.38 to 1.42] | 116 | 30.2 fewer (71.9 fewer to 48.7 more) | Very low | due to risk of bias, imprecision (x2) | 1 | 198 | 0.74 [0.38 to 1.42] | 116 | 30.2 fewer (71.9 fewer to 48.7 more) | Very low | due to risk of bias, imprecision (x2) |
| 3 months | 1 | 198 | 0.74 [0.38 to 1.42] | 116 | 30.2 fewer (71.9 fewer to 48.7 more) | Very low | due to risk of bias, imprecision (x2) | 1 | 198 | 0.74 [0.38 to 1.42] | 116 | 30.2 fewer (71.9 fewer to 48.7 more) | Very low | due to risk of bias, imprecision (x2) |
| 6 months | 1 | 198 | 0.74 [0.38 to 1.42] | 116 | 30.2 fewer (71.9 fewer to 48.7 more) | Very low | due to risk of bias, imprecision (x2) | 1 | 198 | 0.74 [0.38 to 1.42] | 116 | 30.2 fewer (71.9 fewer to 48.7 more) | Very low | due to risk of bias, imprecision (x2) |
| Current | 5 | 8474 | 1.14 [1.02 to 1.26] | 116 | 16.2 more (2.3 fewer to 30.2 more) | Low | due to risk of bias, imprecision | 5 | 8474 | 1.14 [1.02 to 1.26] | 116 | 16.2 more (2.3 fewer to 30.2 more) | Low | due to risk of bias, imprecision |
| (Hydroxy)chloroquine (Treatment) |  |  |  |  |  |  |  |  |  |  |  |  |  |  |
| 1 month | 2 | 642 | 1.14 [0.61 to 2.12] | 116 | 16.2 more (45.2 fewer to 129.9 more) | Very low | due to risk of bias, imprecision (x3) | 4 | 4616 | 1.15 [0.94 to 1.39] | 116 | 17.4 more (7.0 fewer to 45.2 more) | Low | due to risk of bias, imprecision |
| 3 months | 4 | 4693 | 1.17 [0.96 to 1.42] | 116 | 19.7 more (4.6 fewer to 46.4 more) | Low | due to risk of bias, imprecision | 6 | 6430 | 1.15 [0.97 to 1.35] | 116 | 17.4 more (3.5 fewer to 40.6 more) | Low | due to risk of bias, imprecision |
| 6 months | 7 | 6877 | 1.13 [0.96 to 1.32] | 116 | 15.1 more (4.6 fewer to 37.1 more) | Low | due to risk of bias, imprecision | 9 | 7417 | 1.11 [0.96 to 1.29] | 116 | 12.8 more (4.6 fewer to 33.6 more) | Low | due to risk of bias, imprecision |
| Current | 12 | 8053 | 1.23 [1.05 to 1.46] | 116 | 26.7 more (5.8 more to 53.4 more) | Moderate | due to risk of bias | 12 | 8053 | 1.23 [1.05 to 1.46] | 116 | 26.7 more (5.8 more to 53.4 more) | Moderate | due to risk of bias |
| Ivermectin |  |  |  |  |  |  |  |  |  |  |  |  |  |  |
| 1 month | 1 | 45 | NA (0 events) | 116 | NA | NA | NA | 2 | 90 | 1.52 [0.07 to 35.28] | 116 | 60.3 more (107.9 fewer to 496.5 more) | Very low | due to risk of bias, imprecision (x3) |
| 3 months | 1 | 45 | NA (0 events) | 116 | NA | NA | NA | 4 | 354 | 0.40 [0.06 to 2.46] | 116 | 69.6 fewer (109 fewer to 169.4 more) | Very low | due to risk of bias, imprecision (x3) |
| 6 months | 4 | 642 | 0.98 [0.55 to 1.72] | 116 | 2.3 fewer (52.2 fewer to 83.5 more) | Low | due to imprecision (x2) | 7 | 951 | 0.77 [0.36 to 1.65] | 116 | 26.7 fewer (74.2 fewer to 75.4 more) | Very low | due to imprecision (x3) |
| Current | 8 | 1464 | 0.94 [0.58 to 1.53] | 116 | 7.0 fewer (48.7 fewer to 61.5 more) | Low | due to imprecision (x2) | 9 | 1616 | 0.94 [0.58 to 1.53] | 116 | 7.0 fewer (48.7 fewer to 61.5 more) | Low | due to imprecision (x2) |
| IL-6 receptor blockers |  |  |  |  |  |  |  |  |  |  |  |  |  |  |
| 1 month | 0 | 0 | NA | 116 | NA | NA | NA | 1 | 273 | 0.76 [0.53 to 1.09] | 116 | 27.8 fewer (54.5 fewer to 10.4 more) | Low | due to risk of bias, imprecision |
| 3 months | 3 | 495 | 0.68 [0.43 to 1.09] | 116 | 37.1 fewer (66.1 fewer to 10.4 more) | Low | due to risk of bias, imprecision | 5 | 1145 | 0.71[0.55 to 0.93] | 116 | 33.6 fewer (52.2 fewer to 8.1 fewer) | Moderate | due to risk of bias |
| 6 months | 7 | 1826 | 0.74 [0.63 to 0.86] | 116 | 30.2 fewer (42.9 fewer to 16.2 fewer) | Moderate | due to risk of bias | 9 | 4000 | 0.82 [0.73 to 0.93] | 116 | 20.9 fewer (31.3 fewer to 8.1 fewer) | Moderate | due to risk of bias |
| Current | 10 | 4170 | 0.83 [0.74 to 0.93] | 116 | 19.7 fewer (30.2 fewer to 8.1 fewer) | Moderate | due to risk of bias | 12 | 4560 | 0.83 [0.74 to 0.92] | 116 | 19.7 fewer (30.2 fewer to 9.3 fewer) | Moderate | due to risk of bias |
| Convalescent plasma |  |  |  |  |  |  |  |  |  |  |  |  |  |  |
| 1 month | 1 | 464 | 1.08 [0.59 to 1.99] | 116 | 9.3 more (47.6 fewer to 104.4 more) | Very low | due to risk of bias, imprecision (x2) | 3 | 705 | 0.83 [0.39 to 1.78] | 116 | 19.7 fewer (70.8 fewer to 90.5 more) | Very low | due to risk of bias, imprecision (x2) |
| 3 months | 3 | 827 | 1.14 [0.81 to 1.61] | 116 | 16.2 more (22 fewer to 70.8 more) | Very low | due to risk of bias, imprecision (x2) | 6 | 1108 | 1.04 [0.75 to 1.42] | 116 | 4.6 more (29 fewer to 48.7 more) | Very low | due to risk of bias, imprecision (x2) |
| 6 months | 4 | 987 | 1.11 [0.79 to 1.54] | 116 | 12.8 more (24.4 fewer to 62.6 more) | Very low | due to risk of bias, imprecision (x2) | 6 | 1108 | 1.04 [0.75 to 1.42] | 116 | 4.6 more (29 fewer to 48.7 more) | Very low | due to risk of bias, imprecision (x2) |
| Current | 8 | 8252 | 0.98 [0.90 to 1.06] | 116 | 2.3 fewer (11.6 fewer to 7 more) | Moderate | due to risk of bias | 9 | 8333 | 0.98 [0.90 to 1.05] | 116 | 2.3 fewer (11.6 fewer to 5.8 more) | Moderate | due to risk of bias |

## References

1. Horby P, Lim WS, Emberson J, Mafham M, Bell J, Linsell L, et al. Effect of Dexamethasone in Hospitalized Patients with COVID-19: Preliminary Report. medRxiv. 2020:2020.06.22.20137273.

2. Horby P, Lim WS, Emberson JR, Mafham M, Bell JL, Linsell L, et al. Dexamethasone in Hospitalized Patients with Covid-19 - Preliminary Report. N Engl J Med. 2020.

3. Horby P, Mafham M, Linsell L, Bell JL, Staplin N, Emberson JR, et al. Effect of Hydroxychloroquine in Hospitalized Patients with COVID-19: Preliminary results from a multi-centre, randomized, controlled trial. medRxiv. 2020:2020.07.15.20151852.

4. Horby P, Mafham M, Linsell L, Bell JL, Staplin N, Emberson JR, et al. Effect of Hydroxychloroquine in Hospitalized Patients with Covid-19. N Engl J Med. 2020;383(21):2030–40.

5. Pan H, Peto R, Karim QA, Alejandria M, Henao-Restrepo AM, García CH, et al. Repurposed antiviral drugs for COVID-19 –interim WHO SOLIDARITY trial results. medRxiv. 2020:2020.10.15.20209817.

6. Pan H, Peto R, Henao-Restrepo AM, Preziosi MP, Sathiyamoorthy V, Abdool Karim Q, et al. Repurposed Antiviral Drugs for Covid-19 - Interim WHO Solidarity Trial Results. N Engl J Med. 2020.

7. Maslove DM. Medical Preprints-A Debate Worth Having. Jama. 2018;319(5):443–4.

8. Lauer MS, Krumholz HM, Topol EJ. Time for a prepublication culture in clinical research? Lancet. 2015;386(10012):2447–9.

9. Carneiro CF, Queiroz VG, Moulin TC, Carvalho CA, Haas CB, Rayêe D, et al. Comparing quality of reporting between preprints and peer-reviewed articles in the biomedical literature. BioRxiv. 2019:581892.

10. Walker R, Rocha da Silva P. Emerging trends in peer review-a survey. Front Neurosci. 2015;9:169.

11. Chan AW, Song F, Vickers A, Jefferson T, Dickersin K, Gøtzsche PC, et al. Increasing value and reducing waste: addressing inaccessible research. Lancet. 2014;383(9913):257–66.

12. Franco A, Malhotra N, Simonovits G. Social science. Publication bias in the social sciences: unlocking the file drawer. Science. 2014;345(6203):1502–5.

13. Schmucker C, Schell LK, Portalupi S, Oeller P, Cabrera L, Bassler D, et al. Extent of non-publication in cohorts of studies approved by research ethics committees or included in trial registries. PLoS One. 2014;9(12):e114023.

14. Scherer RW, Meerpohl JJ, Pfeifer N, Schmucker C, Schwarzer G, von Elm E. Full publication of results initially presented in abstracts. Cochrane Database Syst Rev. 2018;11(11):Mr000005.

15. Rising K, Bacchetti P, Bero L. Reporting bias in drug trials submitted to the Food and Drug Administration: review of publication and presentation. PLoS Med. 2008;5(11):e217. discussion e.

16. van Schalkwyk MCI, Hird TR, Maani N, Petticrew M, Gilmore AB. The perils of preprints. Bmj. 2020;370:m3111.

17. Flanagin A, Fontanarosa PB, Bauchner H. Preprints Involving Medical Research-Do the Benefits Outweigh the Challenges? Jama. 2020;324(18):1840–3.

18. Siemieniuk RA, Bartoszko JJ, Ge L, Zeraatkar D, Izcovich A, Kum E, et al. Drug treatments for covid-19: living systematic review and network meta-analysis. Bmj. 2020;370:m2980.

19. Siemieniuk RA, Bartoszko JJ, Díaz Martinez JP, Kum E, Qasim A, Zeraatkar D, et al. Antibody and cellular therapies for treatment of covid-19: a living systematic review and network meta-analysis. Bmj. 2021;374:n2231.

20. Bartoszko JJ, Siemieniuk RAC, Kum E, Qasim A, Zeraatkar D, Ge L, et al. Prophylaxis against covid-19: living systematic review and network meta-analysis. Bmj. 2021;373:n949.

21. Marshall IJ, Noel-Storr A, Kuiper J, Thomas J, Wallace BC. Machine learning for identifying Randomized Controlled Trials: An evaluation and practitioner’s guide. Res Synth Methods. 2018;9(4):602–14.

22. Sterne JAC, Savović J, Page MJ, Elbers RG, Blencowe NS, Boutron I, et al. RoB 2: a revised tool for assessing risk of bias in randomised trials. Bmj. 2019;366:l4898.

23. Lamontagne F, Agoritsas T, Macdonald H, Leo YS, Diaz J, Agarwal A, et al. A living WHO guideline on drugs for covid-19. Bmj. 2020;370:m3379.

24. Centers for Disease Control and Prevention. COVIDView. A weekly surveillance summary of U.S COVID-19 activity 2020 [Available from:.

26. ISARIC (International Severe Acute Respiratory and Emerging Infections Consortium). COVID-19 Report: 08 June 2020. medRxiv 2020.

27. Guyatt GH, Oxman AD, Vist GE, Kunz R, Falck-Ytter Y, Alonso-Coello P, et al. GRADE: an emerging consensus on rating quality of evidence and strength of recommendations. Bmj. 2008;336(7650):924–6.

28. Hultcrantz M, Rind D, Akl EA, Treweek S, Mustafa RA, Iorio A, et al. The GRADE Working Group clarifies the construct of certainty of evidence. J Clin Epidemiol. 2017;87:4–13.

29. Vlaar APJ, de Bruin S, Busch M, Timmermans S, van Zeggeren IE, Koning R, et al. Anti-C5a antibody IFX-1 (vilobelimab) treatment versus best supportive care for patients with severe COVID-19 (PANAMO): an exploratory, open-label, phase 2 randomised controlled trial. Lancet Rheumatol. 2020;2(12):e764–e73.

30. Vlaar APJadB, Sanne and Busch, Matthias and Timmermans, Sjoerd and van Zeggeren, Ingeborg E. and Koning, Rutger and ter Horst, Liora and Bulle, Esther B. and van Baarle, Frank E.H.P. and van de Poll, Marcel C.G. and Kemper, E. Marleen and van der Horst, Iwan C.C. and Schultz, Marcus J. and Horn, Janneke and Paulus, Frederique and Bos, Lieuwe D. and Wiersinga, W. Joost and Witzenrath, Martin and Rueckinger, Simon and Pilz, Korinna and Brouwer, Matthijs C. and Guo, Ren-Feng and Heunks, Leo and van Paassen, Pieter and Riedemann, Niels. C. and van de Beek, Diederik, Anti-C5a Antibody (IFX-1) Treatment of Severe COVID-19: An Exploratory Phase 2 Randomized Controlled Trial..

31. Horby P, Lim WS, Emberson J, Mafham M, Bell J, Linsell L, et al. Effect of Dexamethasone in Hospitalized Patients with COVID-19 – Preliminary Report. medRxiv. 2020:2020.06.22.20137273.

32. Ivashchenko AA, Dmitriev KA, Vostokova NV, Azarova VN, Blinow AA, Egorova AN, et al. AVIFAVIR for Treatment of Patients With Moderate Coronavirus Disease 2019 (COVID-19): Interim Results of a Phase II/III Multicenter Randomized Clinical Trial. Clin Infect Dis. 2021;73(3):531–4.

33. Ivashchenko AA, Dmitriev KA, Vostokova NV, Azarova VN, Blinow AA, Egorova AN, et al. AVIFAVIR for Treatment of Patients with Moderate COVID-19: Interim Results of a Phase II/III Multicenter Randomized Clinical Trial. medRxiv. 2020:2020.07.26.20154724.

34. Elgazzar A, Hany B, Youssef SA, Hany B, Hafez M, Moussa H. Efficacy and Safety of Ivermectin for Treatment and prophylaxis of COVID-19 Pandemic. Research Square. 2021.

35. Brown N. A bug and a dilemma 2021 [Available from:.

36. Dabbous HM, Abd-Elsalam S, El-Sayed MH, Sherief AF, Ebeid FFS, El Ghafar MSA, et al. Efficacy of favipiravir in COVID-19 treatment: a multi-center randomized study. Arch Virol. 2021;166(3):949–54.

37. Dabbous HM, Abd-Elsalam S, El-Sayed MH, Sherief AF, Ebeid FFS, El Ghafar MSA, et al. Retraction Note to: Efficacy of favipiravir in COVIDIZI19 treatment: a multiIZIcenter randomized study. Arch Virol. 2021:1.

38. Dabbous HM, El-Sayed MH, El Assal G, Elghazaly H, Ebeid FFS, Sherief AF, et al. Retraction Note: Safety and efficacy of favipiravir versus hydroxychloroquine in management of COVID-19: A randomised controlled trial. Sci Rep. 2021;11(1):18983.

39. Dabbous HM, El-Sayed MH, El Assal G, Elghazaly H, Ebeid FFS, Sherief AF, et al. Safety and efficacy of favipiravir versus hydroxychloroquine in management of COVID-19: A randomised controlled trial. Sci Rep. 2021;11(1):7282.

40. Samaha AA, Mouawia H, Fawaz M, Hassan H, Salami A, Bazzal AA, et al. Effects of a Single Dose of Ivermectin on Viral and Clinical Outcomes in Asymptomatic SARS-CoV-2 Infected Subjects: A Pilot Clinical Trial in Lebanon. Viruses. 2021;13(6).

41. Samaha AA, Mouawia H, Fawaz M, Hassan H, Salami A, Bazzal AA, et al. Retraction: Samaha et al. Effects of a Single Dose of Ivermectin on Viral and Clinical Outcomes in Asymptomatic SARS-CoV-2 Infected Subjects: A Pilot Clinical Trial in Lebanon. Viruses 2021, 13, 989. Viruses. 2021;13(11).

42. Carlisle JB. The analysis of 168 randomised controlled trials to test data integrity. Anaesthesia. 2012;67(5):521–37.

43. Carlisle JB. Data fabrication and other reasons for non-random sampling in 5087 randomised, controlled trials in anaesthetic and general medical journals. Anaesthesia. 2017;72(8):944–52.

44. Bordewijk EM, Li W, van Eekelen R, Wang R, Showell M, Mol BW, et al. Methods to assess research misconduct in health-related research: A scoping review. J Clin Epidemiol. 2021;136:189–202.

45. Kirkwood AA, Cox T, Hackshaw A. Application of methods for central statistical monitoring in clinical trials. Clin Trials. 2013;10(5):783–806.

46. van den Bor RM, Vaessen PWJ, Oosterman BJ, Zuithoff NPA, Grobbee DE, Roes KCB. A computationally simple central monitoring procedure, effectively applied to empirical trial data with known fraud. J Clin Epidemiol. 2017;87:59–69.

47. Carlisle JB, Dexter F, Pandit JJ, Shafer SL, Yentis SM. Calculating the probability of random sampling for continuous variables in submitted or published randomised controlled trials. Anaesthesia. 2015;70(7):848–58.

48. Oikonomidi T, Boutron I, Pierre O, Cabanac G, Ravaud P, the C-NMAC. Changes in evidence for studies assessing interventions for COVID-19 reported in preprints: meta-research study. BMC Medicine. 2020;18(1):402.

49. Bero L, Lawrence R, Leslie L, Chiu K, McDonald S J Page M, et al. Comparison of preprints and final journal publications from COVID-19 Studies: Discrepancies in results reporting and spin in interpretation. medRxiv. 2021:2021.04.12.21255329.

