## Supplementary material for "The trustworthiness and impact of trial preprints for COVID-19 decision-making: A methodological study"

Dena Zeraatkar, PhD

Department of Biomedical Informatics, Harvard Medical School

Department of Health Research Methods, Evidence, and Impact, McMaster University

1280 Main St W, Hamilton, ON L8S 4L8, Canada (905 525 9140 ext. 26771)

### Supplement 1 – Study protocol

### The credibility and utility of trial preprints during the COVID-19 pandemic: A protocol for a methodological study

#### Background

Clinicians and other decision makers need rapidly available and credible data addressing the comparative effectiveness of potential treatments and prophylaxis for the coronavirus disease 2019 (COVID-19). During the COVID-19 pandemic, the scientific community has adopted preprint servers, which allow investigators to disseminate research findings before publication in peer-reviewed journals.

Growing interest in preprints predates the COVID-19 pandemic (1, 2). Researchers and evidence users have raised concerns that the traditional publication model is slow, peer review may not always improve the quality of manuscripts, journals impede dissemination due to paywalls and high publication fees and encourage publication bias by prioritizing statistically significant or anomalous findings—issues preprints may avoid (3-9). Despite these concerns, and the potential of preprints to address them, because preprints may result in the dissemination of provisional findings that contain important errors, the medical community has been cautious about their adoption (10, 11). Authors of systematic reviews, guideline developers, and other decision makers face a trade-off when considering preprints: on the one hand inclusion could reduce the credibility of evidence syntheses and risk serious errors if important differences appear in later published reports; on the other, including preprints may increase the precision of estimates, allow timely dissemination, and minimize the effects of publication bias.

Knowledge of the extent to which preprints may accelerate the dissemination of findings, the frequency and nature of discrepancies between pre-prints and subsequent reports, and their impact on meta-analytic estimates could inform the trade-off that evidence users face. Our study will capitalize on the methods and data of our living systematic review and network meta-analysis (SRNMA) of drug treatments, antiviral antibodies and cellular therapies, and prophylaxis for COVID-19—an initiative launched in July 2020 that provides real-time summaries addressing the comparative effectiveness of potential treatments and prophylaxis for COVID-19—to report on the characteristics, credibility, and utility of COVID-19 trial preprint reports (12). We define credibility as complete and consistent reporting of key aspects of the methods and results between preprint and published trial reports and utility as the contribution of preprint reports to narrow confidence intervals and produce higher certainty evidence.

#### Methods

Patient and Public Involvement

Patients were involved in outcome selection, interpretation of results, and the generation of parallel recommendations, as part of the BMJ Rapid Recommendations initiative.

Search

Our study will use the search strategy of our living SRNMA that includes daily searches of the World Health Organization (WHO) COVID-19 database—a comprehensive multilingual source of global published and preprint literature on COVID-19 (https://search.bvsalud.org/global-literature-on-novel-coronavirus-2019-ncov/). Prior to its merge with the WHO COVID-19 database on 9 October 2020, we searched the US Centers for Disease Control and Prevention (CDC) COVID-19 Research Articles Downloadable Database. We use a validated machine learning model to identify randomized controlled trials (13). We also search six Chinese databases monthly: Wanfang, Chinese Biomedical Literature, China National Knowledge Infrastructure, VIP, Chinese Medical Journal Net (preprints), and ChinaXiv (preprints).

Our search is supplemented by ongoing surveillance of living evidence retrieval services, including the Living Overview of the Evidence (L-OVE) COVID-19 platform by the Epistemonikos Foundation (https://app.iloveevidence.com/loves/5e6fdb9669c00e4ac072701d) and the Systematic and Living Map on COVID-19 Evidence by the Norwegian Institute of Public Health (<https://www.fhi.no/en/qk/systematic-reviews-hta/map/>).

Supplementary 1 includes additional details of our search strategy.

##### Study selection

As part of the living SRNMA, pairs of reviewers, following calibration exercises, work independently and in duplicate to screen titles and abstracts of search records and subsequently the full texts of records determined potentially eligible at the title and abstract screening stage. Reviewers also link preprint reports with their subsequent publications based on trial registration numbers, authors, and other trial characteristics. Reviewers resolve discrepancies by discussion or, when necessary, by adjudication with a third-party reviewer.

We include preprint and peer reviewed reports of trials that randomize patients with suspected, probable, or confirmed COVID-19 to drug treatments, antiviral antibodies and cellular therapies, placebo, or standard care or trials that randomize healthy participants exposed or unexposed to COVID-19 to prophylactic drugs, standard care, or placebo. We do not apply any restrictions based on severity of illness, setting, or language of publication. We exclude trials that report on nutritional interventions, traditional Chinese herbal medicines without standardization in formulations and dosing across batches, and non-drug supportive care interventions.

For this project, we will include all eligible trial reports identified through our living SRNMA.

##### Data collection

As part of the living SRNMA, for each eligible trial, pairs of reviewers, following training and calibration exercises, independently extract trial characteristics, methods, and results using a standardized, pilot tested data extraction form. To assess risk of bias, reviewers, following training and calibration exercises, use a revision of the Cochrane tool for assessing risk of bias in randomized trials (RoB 2.0) (14) (Supplementary 2). Reviewers resolve discrepancies by discussion and, when necessary, by adjudication with a third party.

For the current study, pairs of trained and calibrated reviewers, working independently and in duplicate and using a pilot-tested form, will collect data on differences between preprint and published trial reports in key methods and results. Key methods include description of the randomization process and allocation concealment, blinding of patients and healthcare providers, extent of and handling of missing outcome data, blinding of outcome assessors and adjudicators, and prespecification of outcomes and analyses. For key methods, we will consider discrepancies that may affect the rating of risk of bias. Key results include number of participants analyzed and means or medians and measures of variability for continuous outcomes and the number of events for dichotomous outcomes. We will focus on the same outcomes as our living SRNMA: mortality, mechanical ventilation, adverse events leading to discontinuation, viral clearance, admission to hospital, viral clearance, hospital length of stay, ICU length of stay, duration of mechanical ventilation, time to symptom resolution or clinical improvement, time to viral clearance, days free from mechanical ventilation, and time to viral clearance. For preprints with more than one version, we will extract data from the first version of the preprint, which is the least likely to have been modified in response to peer review.

##### Data synthesis and analysis

We will compare the characteristics of trials with versus without preprints, including country of recruitment, registration, study status, type of interventions studied (drug therapy, antiviral antibodies and cellular therapies, or prophylaxis), severity of disease (inpatient/outpatient and whether patients were severe/critical), number of centers, number of participants, statistical significance of primary and secondary outcomes (based on cut-offs defined by the authors or, when no cut-offs are defined, based on a cut-off of p<0.05 or confidence intervals not including the null), risk of bias, and source of funding, by calculating differences in proportions and associated 95% confidence intervals. Because risk of bias may vary across outcomes, we will present risk of bias ratings corresponding to the following hierarchy which represents the relative importance of outcomes for clinical decisions and recommendations: mortality, mechanical ventilation, duration of hospitalization, time to symptom resolution or clinical improvement, and virologic outcomes. For prophylaxis trials, we will use the following hierarchy: mortality, laboratory confirmed and suspected COVID-19 infection, and laboratory confirmed COVID-19 infection.

We will calculate the median time from a trial being posted on a preprint server to its eventual publication in a journal and will assess whether source of funding, number of centers and participants, intensity of care (inpatient versus outpatient), early termination for benefit, statistically significant primary or secondary outcomes (based on cut-offs defined by the authors or, when no cut-offs were defined, based on a cut-off of p<0.05 or confidence intervals not including the null), and risk of bias are predictive of time to publication using Kaplan-Meier curves with log-rank tests. We anticipate large, multicenter trials, industry-funded trials, trials that are terminated early for benefit, trials that report on inpatients, trials with statistically significant results, and trials at low risk of bias to be published faster.

We will describe the number and types of discrepancies in the reporting and presentation of key methods and results between preprint and published trial reports. For discrepancies in the reporting of methods, we will assess whether the differences changed risk of bias ratings.

To investigate differences in meta-analyses that include versus exclude evidence from preprints, we will focus on interventions that have been addressed by the WHO living guideline (15), IL-6 receptor blockers, ivermectin, hydroxychloroquine, lopinavir-ritonavir, remdesivir, and corticosteroids, and the two most commonly reported outcomes in trials (i.e., mortality, mechanical ventilation). For these interventions and outcomes, we will conduct pairwise frequentist random-effects meta-analyses with the restricted maximum likelihood estimator that include and exclude evidence from available preprints at one, three, and six months after the first trial preprint or published report addressing the intervention of interest became publicly available.

To facilitate interpretation, we will use baseline risks from the CDC and International Severe Acute Respiratory and Emerging Infection COVID-19 database to calculate absolute effects (16-18). We will assess the certainty of evidence using GRADE approach and report whether including versus excluding preprint reports leads to important differences in the effect estimates, ratings of the overall certainty of evidence, judgments related to specific domains of GRADE, and whether differences in ratings are likely to impact decision making (i.e., evidence rated as high/moderate is downgraded to low/very low or vice versa) (19).

We will consider differences in effect estimates important if a meta-analysis including preprints suggests benefit and a meta-analysis excluding preprints suggests harm, or vice versa, or if a meta-analysis including preprints suggests no effect and a meta-analysis excluding preprints suggests benefit or harm, or vice versa. Judgments of imprecision will be made using a minimally contextualized approach. The minimally contextualized approach considers only whether confidence intervals include the null effect and thus does not consider whether confidence intervals include both important and trivial effects. To evaluate certainty of no effect, we will use a 1% risk difference threshold for mortality and a 2% risk difference for mechanical ventilation (20).

#### Discussion

Clinicians and decision-makers need rapidly available and credible data on the comparative effectiveness of potential treatments and prophylaxis for COVID-19. Preprints have become central venues through which trial authors can quickly disseminate their findings (1-4, 21-23). Authors of seminal COVID-19 trials, for example, representing massive international collaborations, such as RECOVERY (24-27) and SOLIDARITY (28, 29), chose to report their results in preprints before subsequent publication in journals. Evidence users have, however, expressed concerns about the credibility of trial preprints (10, 11).

Our study will present a detailed assessment of the credibility and utility of COVID-19 trial preprint reports. We will show the extent to which preprints accelerate time to dissemination of trial findings, differences between preprints and their subsequent published reports in key methods and results, and test whether including preprints in meta-analyses improves the precision and overall certainty of evidence.

##### Implications

Our findings will have implications for evidence users and decision makers who are concerned with the credibility of preprint reports and for systematic reviewers and guideline developers deciding whether to consider preprint reports in systematic reviews and guideline recommendations. Evidence that preprints accelerate dissemination of findings, do not report results which are inconsistent with published trial reports, and that including preprint reports in systematic reviews results in higher certainty evidence will lend further support to the credibility and utility of preprints for consideration in systematic reviews and guidelines. Opposite results will mandate consideration of excluding preprints. Future health emergencies will also necessitate rapid dissemination of research and our study will inform whether evidence-users can confidently rely on preprint trial reports during health emergencies.

##### Relation to previous work

Our study will be the first to present data addressing the relative contribution of preprint reports to the evidence regarding the comparative effectiveness of COVID-19 therapies and prophylaxis, and to test the robustness of meta-analyses and conclusions that include versus exclude preprint reports.

Two studies have reported on differences between preprint and published study reports and citations and Altmetric attention metrics (30, 31). One study additionally addressed publication characteristics and dissemination of COVID-19 preprints and the other spin in interpretation of results. Both studies were, however, restricted to publications up to August and October 2020—which is not representative of the current landscape of COVID-19 research and which does not include the majority of evidence being currently used to guide COVID-19 care, including critical trials addressing the effects of corticosteroids (24, 25). These studies also included all study designs rather than focusing only on randomized trials that are primarily used to guide clinical decisions and recommendations (15, 32), and did not compare the effects of including preprints on meta-analytic estimates and the certainty of the body of evidence (30). The latter issue is particularly important because evidence users use the totality of the body of evidence, rather than single studies, to make treatment decisions and recommendations.

##### Strengths and limitations

The strengths of this study include the comprehensive search for and inclusion of preprint and published COVID-19 trial reports and rigorous data collection. We also focus on the implications of preprints for evidence users and decision-makers rather than only on only discrepancies between preprints and publications that may not matter importantly. The generalizability of our results is, however, limited to COVID-19. Journals have been expediting publication of COVID-19 research and have been publishing more prolifically on COVID-19 than in other areas, which may reduce opportunity for revisions between preprints and their subsequent publications and may mean time to publication and predictors of publication may be different than in other research areas.

Although the WHO COVID-19 database is a comprehensive source of published and preprint literature, it does not include all preprint servers—though preprint servers not covered by our search address other subjects and are unlikely to include COVID-19 trials.

We will limit our assessment of the effects of including versus excluding preprint reports on meta-analytic estimates and the certainty of evidence to only interventions that have been addressed by the WHO living guideline. It is possible that preprint reports of trials that are subsequently published in journals represent the most rigorous or transparently reported preprints and that they are not representative of all trial preprints. Our estimate of the time to publication of preprint reports may be overestimated if some preprint authors did not attempt to subsequently publish in peer-reviewed journals—although evidence shows that most preprint authors of COVID-19 studies intend to publish their findings (30). Finally, although we will describe discrepancies in the reporting of key methods and results between preprint and published trial reports, we will not assess differences in the discussion or conclusion sections of trial reports and the interpretation of findings. It is possible that preprint reports may contain more spin and positive interpretation of results compared to published trial reports (31).

#### References

1.	Maslove DM. Medical Preprints-A Debate Worth Having. Jama. 2018;319(5):443-4.

2.	Lauer MS, Krumholz HM, Topol EJ. Time for a prepublication culture in clinical research? Lancet. 2015;386(10012):2447-9.

3.	Carneiro CF, Queiroz VG, Moulin TC, Carvalho CA, Haas CB, Rayêe D, et al. Comparing quality of reporting between preprints and peer-reviewed articles in the biomedical literature. BioRxiv. 2019:581892.

4.	Walker R, Rocha da Silva P. Emerging trends in peer review-a survey. Front Neurosci. 2015;9:169.

5.	Chan AW, Song F, Vickers A, Jefferson T, Dickersin K, Gøtzsche PC, et al. Increasing value and reducing waste: addressing inaccessible research. Lancet. 2014;383(9913):257-66.

6.	Franco A, Malhotra N, Simonovits G. Social science. Publication bias in the social sciences: unlocking the file drawer. Science. 2014;345(6203):1502-5.

7.	Schmucker C, Schell LK, Portalupi S, Oeller P, Cabrera L, Bassler D, et al. Extent of non-publication in cohorts of studies approved by research ethics committees or included in trial registries. PLoS One. 2014;9(12):e114023.

8.	Scherer RW, Meerpohl JJ, Pfeifer N, Schmucker C, Schwarzer G, von Elm E. Full publication of results initially presented in abstracts. Cochrane Database Syst Rev. 2018;11(11):Mr000005.

9.	Rising K, Bacchetti P, Bero L. Reporting bias in drug trials submitted to the Food and Drug Administration: review of publication and presentation. PLoS Med. 2008;5(11):e217; discussion e.

10.	van Schalkwyk MCI, Hird TR, Maani N, Petticrew M, Gilmore AB. The perils of preprints. Bmj. 2020;370:m3111.

11.	Flanagin A, Fontanarosa PB, Bauchner H. Preprints Involving Medical Research-Do the Benefits Outweigh the Challenges? Jama. 2020;324(18):1840-3.

12.	Siemieniuk RA, Bartoszko JJ, Ge L, Zeraatkar D, Izcovich A, Kum E, et al. Drug treatments for covid-19: living systematic review and network meta-analysis. Bmj. 2020;370:m2980.

13.	Marshall IJ, Noel-Storr A, Kuiper J, Thomas J, Wallace BC. Machine learning for identifying Randomized Controlled Trials: An evaluation and practitioner's guide. Res Synth Methods. 2018;9(4):602-14.

14.	Sterne JAC, Savović J, Page MJ, Elbers RG, Blencowe NS, Boutron I, et al. RoB 2: a revised tool for assessing risk of bias in randomised trials. Bmj. 2019;366:l4898.

15.	Lamontagne F, Agoritsas T, Macdonald H, Leo YS, Diaz J, Agarwal A, et al. A living WHO guideline on drugs for covid-19. Bmj. 2020;370:m3379.

16.	Centers for Disease Control and Prevention. COVIDView. A weekly surveillance summary of U.S COVID-19 activity 2020 [Available from: <https://www.cdc.gov/coronavirus/2019-ncov/covid-data/covidview/index.html>.

17.	Centers for Disease Control and Prevention. Daily updates of totals by week and state: provisional death counts for coronavirus disease 2019 (COVID-19) 2020 [Available from: <https://www.cdc.gov/nchs/nvss/vsrr/COVID19/index.htm>.

18.	ISARIC (International Severe Acute Respiratory and Emerging Infections Consortium). COVID-19 Report: 08 June 2020. medRxiv 2020.

19.	Guyatt GH, Oxman AD, Vist GE, Kunz R, Falck-Ytter Y, Alonso-Coello P, et al. GRADE: an emerging consensus on rating quality of evidence and strength of recommendations. Bmj. 2008;336(7650):924-6.

20.	Hultcrantz M, Rind D, Akl EA, Treweek S, Mustafa RA, Iorio A, et al. The GRADE Working Group clarifies the construct of certainty of evidence. J Clin Epidemiol. 2017;87:4-13.

21.	Majumder MS, Mandl KD. Early in the epidemic: impact of preprints on global discourse about COVID-19 transmissibility. Lancet Glob Health. 2020;8(5):e627-e30.

22.	Fidahic M, Nujic D, Runjic R, Civljak M, Markotic F, Lovric Makaric Z, et al. Research methodology and characteristics of journal articles with original data, preprint articles and registered clinical trial protocols about COVID-19. BMC Med Res Methodol. 2020;20(1):161.

23.	Smyth AR, Rawlinson C, Jenkins G. Preprint servers: a 'rush to publish' or 'just in time delivery' for science? Thorax. 2020;75(7):532-3.

24.	Horby P, Lim WS, Emberson J, Mafham M, Bell J, Linsell L, et al. Effect of Dexamethasone in Hospitalized Patients with COVID-19: Preliminary Report. medRxiv. 2020:2020.06.22.20137273.

25.	Horby P, Lim WS, Emberson JR, Mafham M, Bell JL, Linsell L, et al. Dexamethasone in Hospitalized Patients with Covid-19 - Preliminary Report. N Engl J Med. 2020.

26.	Horby P, Mafham M, Linsell L, Bell JL, Staplin N, Emberson JR, et al. Effect of Hydroxychloroquine in Hospitalized Patients with COVID-19: Preliminary results from a multi-centre, randomized, controlled trial. medRxiv. 2020:2020.07.15.20151852.

27.	Horby P, Mafham M, Linsell L, Bell JL, Staplin N, Emberson JR, et al. Effect of Hydroxychloroquine in Hospitalized Patients with Covid-19. N Engl J Med. 2020;383(21):2030-40.

28.	Pan H, Peto R, Karim QA, Alejandria M, Henao-Restrepo AM, García CH, et al. Repurposed antiviral drugs for COVID-19 –interim WHO SOLIDARITY trial results. medRxiv. 2020:2020.10.15.20209817.

29.	Pan H, Peto R, Henao-Restrepo AM, Preziosi MP, Sathiyamoorthy V, Abdool Karim Q, et al. Repurposed Antiviral Drugs for Covid-19 - Interim WHO Solidarity Trial Results. N Engl J Med. 2020.

30.	Oikonomidi T, Boutron I, Pierre O, Cabanac G, Ravaud P, the C-NMAC. Changes in evidence for studies assessing interventions for COVID-19 reported in preprints: meta-research study. BMC Medicine. 2020;18(1):402.

31.	Bero L, Lawrence R, Leslie L, Chiu K, McDonald S, J Page M, et al. Comparison of preprints and final journal publications from COVID-19 Studies: Discrepancies in results reporting and spin in interpretation. medRxiv. 2021:2021.04.12.21255329.

32.	COVID-19 Living Evidence Map 2020 [Available from: <https://covid19.evidenceprime.ca/>.

### Supplement 2 – Search Strategy

**Search purpose**: Systematic search of the COVID-19 literature performed Monday through Friday for the WHO Database. Searches performed by Tomas Allen, Kavita Kothari, and Martha Knuth.

Use following commands to pull daily new entries:

• Entry_date:( [20210101 TO 20210120])

• Entry_date:( 20210105)

**Duplicates:** Duplicates are found in EndNote and Distillr using the Wichor method. Further screening is done by expert reviewers but some duplicates may still be in the database.

**Daily Search Strategy:**

| **Database** | **Search Strategy** |
| --- | --- |
| Medline  (Ovid)  1946- | (coronavir* OR corona virus* OR corona pandemic* OR betacoronavir* OR covid19 OR covid OR nCoV OR novel CoV OR CoV 2 OR CoV2 OR sarscov2 OR sars2 OR 2019nCoV OR wuhan virus*).mp. OR (sars AND cov).mp. OR ((wuhan OR hubei OR huanan) AND (severe acute respiratory OR pneumonia*) AND outbreak*).mp. OR Coronavirus Infections/ OR Coronavirus/ OR betacoronavirus/  Limits: 2020- |
| CAB Abstracts(Ovid)  1910- | (coronavir* OR corona virus* OR corona pandemic* OR betacoronavir* OR covid19 OR covid OR nCoV OR novel CoV OR CoV 2 OR CoV2 OR sarscov2 OR sars2 OR 2019nCoV OR wuhan virus*).mp. OR (sars AND cov).mp. OR ((wuhan OR hubei OR huanan) AND (severe acute respiratory OR pneumonia*) AND outbreak*).mp. OR exp Betacoronavirus/ |
| Global Health (Ovid)  1910- | (coronavir* OR corona virus* OR corona pandemic* OR betacoronavir* OR covid19 OR covid OR nCoV OR novel CoV OR CoV 2 OR CoV2 OR sarscov2 OR sars2 OR 2019nCoV OR wuhan virus*).mp. OR (sars AND cov).mp. OR ((wuhan OR hubei OR huanan) AND (severe acute respiratory OR pneumonia*) AND outbreak*).mp. OR exp Betacoronavirus/ |
| PsycInfo (Ovid)  1806- | (coronavir* OR corona virus* OR corona pandemic* OR betacoronavir* OR covid19 OR covid OR nCoV OR novel CoV OR CoV 2 OR CoV2 OR sarscov2 OR sars2 OR 2019nCoV OR wuhan virus*).mp. OR (sars AND cov).mp. OR ((wuhan OR hubei OR huanan) AND (severe acute respiratory OR pneumonia*) AND outbreak*).mp.  Limits: 2020- |
| Scopus  1960- | TITLE-ABS-KEY ( coronavir* OR "corona virus" OR "corona pandemic" OR betacoronavir* OR covid19 OR covid OR ncov OR "CoV 2" OR cov2 OR sarscov2 OR sars2 OR 2019ncov OR "novel CoV" OR "wuhan virus" ) OR TITLE-ABS-KEY(sars AND cov) OR ( TITLE-ABS-KEY ( wuhan OR hubei OR huanan ) AND TITLE-ABS-KEY ( "severe acute respiratory" OR pneumonia* ) AND TITLE-ABS-KEY ( outbreak* ) ) AND ( LIMIT-TO ( PUBYEAR , 2021 ) OR LIMIT-TO ( PUBYEAR , 2020 ) ) |
| Academic Search Complete (Ebsco) | TI,AB,SU( ( coronavir* OR "corona virus" OR "corona pandemic" OR betacoronavir* OR covid19 OR covid OR ncov OR "CoV 2" OR cov2 OR sarscov2 OR sars2 OR 2019ncov OR "novel CoV" OR "wuhan virus" ) OR (sars AND cov) OR ( ( wuhan OR hubei OR huanan ) AND ( "severe acute respiratory" OR pneumonia* ) AND ( outbreak* ) ) ) OR ( (MH "Coronavirus") OR (MH "Coronavirus Infections") ) Limits: Dec. 2019-, peer-reviewed |
| Africa Wide Information (Ebsco) | TI,AB,SU( ( coronavir* OR "corona virus" OR "corona pandemic" OR betacoronavir* OR covid19 OR covid OR ncov OR "CoV 2" OR cov2 OR sarscov2 OR sars2 OR 2019ncov OR "novel CoV" OR "wuhan virus" ) OR (sars AND cov) OR ( ( wuhan OR hubei OR huanan ) AND ( "severe acute respiratory" OR pneumonia* ) AND ( outbreak* ) ) ) Limits: 2019-, |
| CINAHL (Ebsco) | TI,AB,SU( ( coronavir* OR "corona virus" OR "corona pandemic" OR betacoronavir* OR covid19 OR covid OR ncov OR "CoV 2" OR cov2 OR sarscov2 OR sars2 OR 2019ncov OR "novel CoV" OR "wuhan virus" ) OR (sars AND cov) OR ( ( wuhan OR hubei OR huanan ) AND ( "severe acute respiratory" OR pneumonia* ) AND ( outbreak* ) ) ) OR ( (MH "Coronavirus") OR (MH "Coronavirus Infections") )  Limits: Dec. 2019-, peer-reviewed |
| ProQuest Central (Proquest)  1952- | TI,AB,SU( ( coronavir* OR "corona virus" OR "corona pandemic" OR betacoronavir* OR covid19 OR covid OR ncov OR "CoV 2" OR cov2 OR sarscov2 OR sars2 OR 2019ncov OR "novel CoV" OR "wuhan virus" ) OR (sars AND cov) OR ( ( wuhan OR hubei OR huanan ) AND ( "severe acute respiratory" OR pneumonia* ) AND ( outbreak* ) ) )  Limits: Dec. 2019-, peer-reviewed |
| China CDC MMWR | Covid OR cov2 OR coronavirus OR “sars cov” OR ncov |
| CDC Reports | Covid OR cov2 OR coronavirus OR “sars cov” OR ncov |
| bioRxiv  medRxiv  chemRxiv (preprints) | Covid OR cov2 OR coronavirus OR “sars cov” OR ncov |
| Embase (Ovid) | ncov OR (('coronavirus'/exp OR coronavirus) AND ('wuhan'/exp OR wuhan)) OR 'novel coronavirus' OR (('pneumonia'/exp OR pneumonia) AND wuhan:ti,ab) OR 'covid' OR 2019ncov OR 'sars-cov'/exp OR 'sars-cov' OR covid OR (('coronavirus'/exp OR coronavirus) AND novel) OR (('corona virus':ti,ab OR 'coronavirus':ti,ab) AND (outbreak:ti,ab OR epidemic*:ti,ab OR pandemdic*:ti,ab OR quaran*:ti,ab OR lockdown*:ti,ab OR syndemic*:ti,ab)) OR hcov OR 'sars virus'/exp OR 'sars virus' OR 'coronavirus disease 2019'/exp OR 'coronavirus disease 2019' OR 'novel coronavirus pneumonia' OR 'covid 19 virus' OR 'severe acute respiratory syndrome coronavirus 2'/exp OR 'severe acute respiratory syndrome coronavirus 2' OR 'coronavirinae'/exp OR 'coronavirinae' OR 'coronavirus infection'/exp OR 'coronavirus infection' OR 'covid19'/exp OR covid19 OR covid2019 OR 'corona pandemic' OR 'sarscov 2' OR 'sarscov-2' OR 'sars co v 2' OR 'social distancing'/exp OR 'social distancing' OR coivd OR 'flatten the curve' OR 'flattening the curve' OR pandoeconom* OR twindemic* OR 'sars voc' |
| Global Index Medicus | (nCov OR (coronavirus AND wuhan) OR "novel coronavirus" OR (pneumonia AND wuhan) OR covid OR 2019ncov OR "sars-cov " OR covid OR (coronavirus AND novel) OR (("corona virus" OR coronavirus ) AND ( ti:outbreak OR ti:epidemic* OR ti:pandemdic* OR ti:quaran* OR ti:syndem* OR hcov OR "sars virus")) OR "coronavirus disease 2019" OR " novel coronavirus pneumonia" OR "COVID 19 virus" OR "severe acute respiratory syndrome coronavirus 2" OR Coronavirinae OR "Coronavirus infection" OR covid19 OR covid2019 OR lockdown* OR "social distancing" OR “physical distancing” OR "corona pandemic" OR "sarscov 2" OR "sarscov-2" OR "sars co v 2" OR coivd OR "flatten the curve" OR "flattening the curve" OR "sars voc") |
| Web of Science | TI=coronavirus OR TI=covid OR TI=Covid19 OR TI=ncov OR TI=(SARS NEAR/3 COV) OR TI="novel coron*virus" OR TI=2019*ncoV OR TI=2019ncov OR TI=(CORON*VIRUS NEAR/3 (OUTBREAK OR pandemic OR 2019 OR new OR novel)) OR TI=coronavirinae OR TI=coronaviridae OR TI=betacoronavirus OR TI=Sars2 OR TI=COV2 OR TI=”corona pandemic” OR ((TI=wuhan OR TI=hubei OR TI=huanan) AND ( TI="severe acute respiratory" OR TI=pneumonia ) AND (TI=outbreak)) |
| PubMed Central | coronavirus[Title] OR "corona virus" [Title] OR "corona pandemic"[Title] OR coronavirinae[Title] OR coronaviridae[Title] OR betacoronavirus[Title] OR covid19[Title] OR covid[Title] OR nCoV[Title] OR "CoV 2"[Title] OR CoV2[Title] OR sars2[Title] OR sarscov2[Title] OR 2019nCoV[Title] OR "novel CoV"[Title] OR "wuhan virus"[Title] OR coronavirus[Abstract] OR "corona virus" [Abstract] OR "corona pandemic"[Abstract] OR coronavirinae[Abstract] OR coronaviridae[Abstract] OR betacoronavirus[Abstract] OR covid19[Abstract] OR covid[Abstract] OR nCoV[Abstract] OR "CoV 2"[Abstract] OR CoV2[Abstract] OR sars2[Abstract] OR sarscov2[Abstract] OR 2019nCoV[Abstract] OR "novel CoV"[Abstract] OR "wuhan virus"[Abstract] OR "COVID-19" [Supplementary Concept] OR "severe acute respiratory syndrome coronavirus 2" [Supplementary Concept] OR ((wuhan[Title] OR hubei[Title] OR huanan[Title]) OR (wuhan[Abstract] OR hubei[Abstract] OR huanan[Abstract]) AND ("severe acute respiratory"[Title] OR pneumonia[Title])) OR (("severe acute respiratory"[Abstract] OR pneumonia[Abstract]) AND (outbreak[Title]) OR outbreak[Abstract]) |
| Science Direct | COVID OR COVID19 OR 2019Ncov OR Ncov OR Coronavirus OR “corona virus” OR (SARS AND Cov) |
| Wiley Online | COVID-19 OR nCov OR 2019ncov OR (pneumonia AND wuhan) OR (sars AND cov) OR COVID OR Covid19 OR “corona virus” OR coronavirus OR COV2 OR SARS2 OR coronavirinae OR coronaviridae OR betacoronavirus OR "corona pandemic" OR ((wuhan OR hubei OR huanan) AND ( "severe acute respiratory" OR pneumonia ) AND (outbreak)) |

### Supplement 3 – Risk of Bias Guidance

| **Bias from the randomization process** | |
| --- | --- |
| Issues to consider:  Random sequence generation  Allocation concealment | |
| **Definitely low risk of bias** | Trials that assign participants to alternative interventions using a randomly generated sequence and maintain allocation concealment.  Examples of methods for developing a randomly generated allocation sequence include a random number generator, random number table, coin tossing, shuffling cards or envelopes, and throwing dice. If a trial is described as 'randomized' without any additional details related to how the allocation sequence was developed, we will assume that the allocation sequence was appropriately developed.  Examples of methods for maintaining allocation concealment include using central allocation via a computer or phone system, pharmacy-controlled allocation, opaque sealed envelopes, and sequentially numbered drug containers.  *Note that an explicit description of random sequence generation is not necessary for a rating of low risk of bias.* |
| **Probably low risk of bias** | Trials in which healthcare providers were blind to the intervention but which provide no information on allocation concealment and in which there are no major baseline imbalances.  *Note that an explicit description of random sequence generation is not necessary for a rating of probably low risk of bias.* |
| **Probably high risk of bias** | Trials in which healthcare providers were not blind to the intervention and which provide no information on allocation concealment.  Trials in which there are substantial baseline differences between trial arms that suggest a problem with the randomization process but there are no other limitations related to randomization. |
| **Definitely high risk of bias** | Trials in which allocation is by judgment of the clinician, by preference of the participant, by availability of the intervention, based on the results of a laboratory test, or other non-random rules (e.g., birthdate, etc.).  Trials in which investigators enrolling participants could possibly foresee the arm to which each subsequent patient would be randomized, such as allocation using an open allocation schedule (e.g. a list of random numbers), assignment envelopes used without appropriate safeguards (e.g. use of unsealed, non-opaque or not sequentially numbered envelopes), alternation between arms, case record number, or any other explicitly unconcealed procedure, rate as high risk. |
| **Bias due to deviations from the intended intervention** | |
| Issues to consider:  Blinding of healthcare providers/clinicians and participants  Imbalances in cointerventions or behaviors | |
| **Definitely low risk of bias** | Therapy trials in which healthcare providers are blind to the intervention administered and in which there are no significant differences in administered co-interventions.  Therapy trials that are described as double or triple blind.  Prophylaxis trials in which participants are blind to the intervention that they have been randomized.  Prophylaxis trials that are described as double or triple blind. |
| **Probably low risk of bias** |  |
| **Probably high risk of bias** | Therapy trials in which healthcare providers are not blind to the intervention administered.  Therapy trials in which healthcare providers are blind to the intervention administered but there are significant differences in administered co-interventions that suggests that blinding may have been compromised.  Therapy trials in which healthcare providers are described as being blind to the intervention but allocation concealment was inadequate.  Prophylaxis trials in which participants are not blind to the intervention that they have been randomized.  Prophylaxis trials in which participants are blind to the intervention to which they have been randomized but there are significant differences in social distancing and risk-taking behaviors that suggest that blinding may have been compromised.  Prophylaxis trials in which healthcare providers are not blind to the intervention and in which healthcare providers were very involved and counselled patients on social distancing, risk-taking behaviors, or testing for COVID-19. |
| **Definitely high risk of bias** | Therapy trials in which healthcare providers are not blind to the intervention and in which there are significant differences in administered co-interventions.  Prophylaxis trials in which participants are not blind to the intervention and in which there are significant differences in social distancing and risk-taking behaviors. |
| **Bias due to missing data** | |
| Issues to consider:  Missing outcome measures  Loss to follow-up | |
| **Definitely low risk of bias** | Trials in which missing outcome data (including outcome data that has been imputed) < 10%.  For in-patient trials, we will assume low risk of bias due to missing data unless otherwise specified. |
| **Probably low risk of bias** | Trials in which missing outcome data (including outcome data that has been imputed) is between 10% to 15% and missing outcome data is unlikely to be related to the true outcome and there is no imbalance in numbers of or reasons for missing data across intervention groups. |
| **Probably high risk of bias** | Trials in which missing outcome data (including outcome data that has been imputed) is between 10% to 15% and missing outcome data is likely to be related to the true outcome or there are imbalances in numbers of or reasons for missing data across intervention groups. |
| **Definitely high risk of bias** | Trials in which missing outcome data (including outcome data that has been imputed) > 15%. |
| **Bias due to measurement of the outcome** | |
| Issues to consider:  Blinding of outcome adjudicators  Objectivity of outcome  *Note that the judgments may differ across outcomes.* | |
| **Definitely low risk of bias** | Trials in which patients are blind to the intervention and in which outcomes are patient-reported.  Trials in which outcomes are measured by a third-party (investigator or clinician) and in which the third-party is blind to the intervention.  Trials in which the outcomes are objective (e.g., mortality, infection with COVID-19 confirmed by a positive RT-PCR swab, mechanical ventilation, admission to hospital, duration of hospital stay, ICU length of stay, ventilator free days, duration of mechanical ventilation, time to clinical improvement if clinical improvement is measured via objective criteria, viral clearance, time to viral clearance).  Trials that are described as double or triple blind. |
| **Probably low risk of bias** |  |
| **Probably high risk of bias** |  |
| **Definitely high risk of bias** | Trials in which patients are not blind and in which outcomes are patient-reported (e.g., time to symptom resolution).  Trials in which outcome adjudicators are not blind and the outcomes are not objective (e.g., adverse effects leading to discontinuation, transfusion-related acute lung injury, transfusion-associated circulatory overload, allergic reactions, infection with suspected/symptomatic COVID-19, venous thromboembolism, time to symptom resolution including fever, time to clinical improvement if the criteria for clinical improvement are not objective). |
| **Bias in selection of the reported results** | |
| Issues to consider:  Selective reporting of timepoints  Selective reporting of outcome measures  *Note that we are only interested in selective reporting for the outcomes for which we are extracting data.*  *Note that the judgments may differ across outcomes.* | |
| **Definitely low risk of bias** | Results for outcomes that were analyzed and reported according to a pre-specified statistical analysis plan or protocol (including the timepoint for the measurement of the outcome). |
| **Probably low risk of bias** | Results for outcomes that were analyzed and reported but that were not prespecified in a statistical analysis plan or protocol but the timepoint at which results are reported is consistent with the timepoint for other outcomes in the trial report or there is little reason to believe the outcome was selectively reported.  Please note that outcomes that were not prespecified in a protocol or statistical analysis plan and that are reported in the trial preprint or publication should be rated at probably low risk of bias unless there are other important reasons to suspect that results for those outcomes were selectively reported (e.g., results are presented at timepoints that don’t match the timepoints reported for other outcomes). |
| **Probably high risk of bias** | Results for outcomes that were analyzed and reported but that were not prespecified in a statistical analysis plan or protocol but the timepoint at which results are reported is not consistent with the timepoint for other outcomes in the trial report or there are other reasons to believe that the outcome is selectively reported. |
| **Definitely high risk of bias** | Results for outcomes that were analyzed and reported for which there are inconsistencies with the statistical analysis plan or protocol. These inconsistencies may include outcome measures of interest or the timepoints for the measurement of outcomes. |
| **Bias due to competing risks** | |
| Issues to consider:  Competing risks due to early termination (only for continuous outcomes) | |
| **Definitely low risk of bias** | Results are very unlikely to have been affected by competing risk due to death.  For example, the intervention arm increased the risk of death but the duration of hospitalization is shorter in the control arm. |
| **Probably low risk of bias** | Results are unlikely to have been affected by competing risk due to death.  For example, the intervention arm increased the risk of death but the duration of hospitalization is slightly shorter in the control arm or there is no appreciable difference between arms. |
| **Probably high risk of bias** | Results are likely to have been affected by competing risk due to death.  For example, the intervention increased the risk of death and the duration of hospitalization is appreciably lower in the intervention arm.  Note that for outcomes such as ICU length of stay and duration of ventilation in which only patients admitted to the ICU or patients are ventilated may be included in analyses, even small imbalances in deaths across trial arms may lead to bias due to competing risks because patients who die are also likely the ones who were admitted to the ICU or ventilated. While patients who die may make up only a small proportion of the total patients included in the trial, they may make up an appreciable proportion of patients who are admitted to the ICU and who are ventilated. |
| **Definitely high risk of bias** | Results are very likely to have been affected by competing risk due to death.  For example, the intervention arm increased the risk of death and the duration of hospitalization is much lower in the intervention arm. |

### Supplement 4 – Study flow

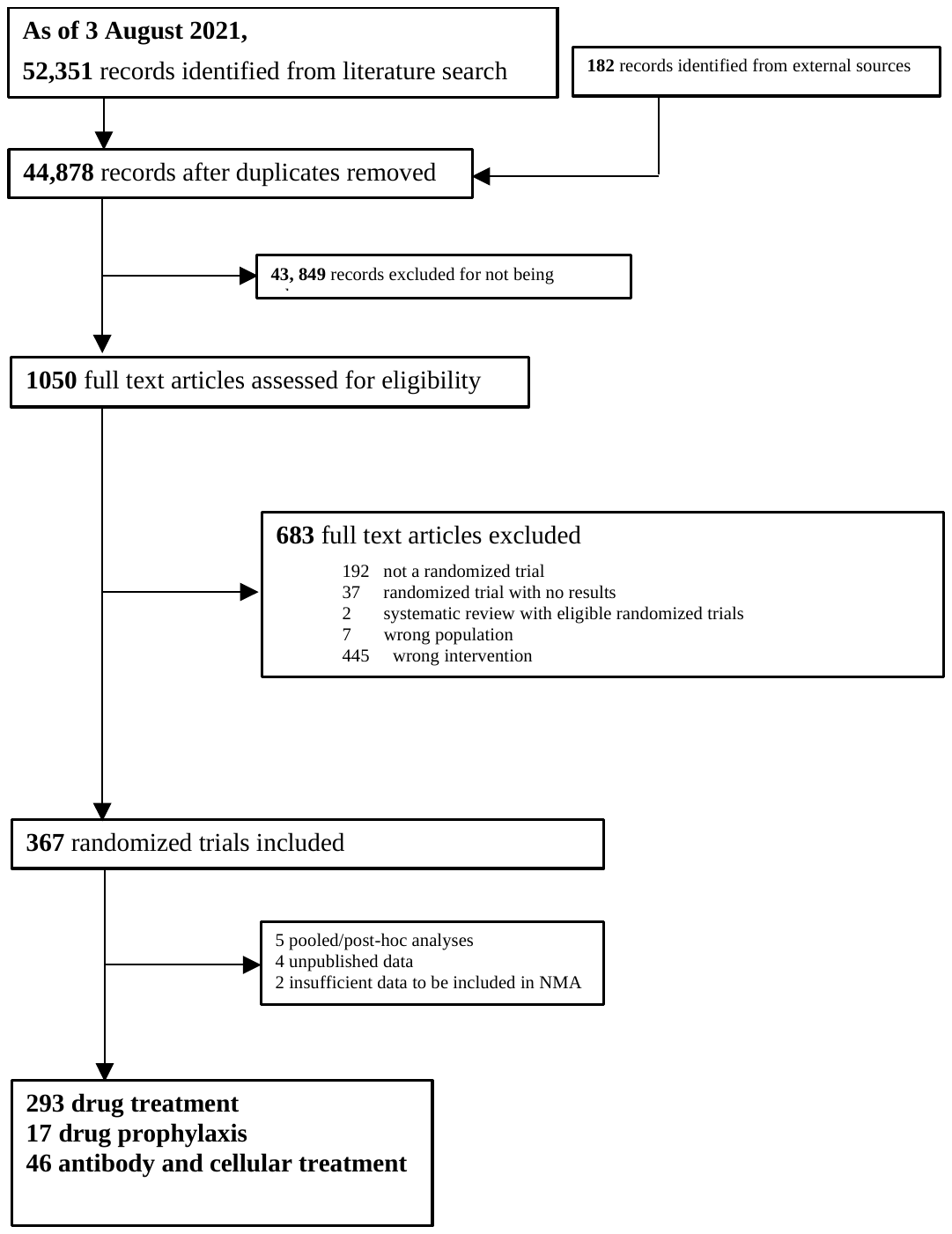

### Supplement 5 – Differences between preprint and published trial reports

| Methods | Number (%) |
| --- | --- |
| The publication reports additional information on allocation concealment. | 8 (10.8%) |
| Resulted in change in RoB (randomization) | 4 (5.4%) |
| The publication reports additional statistic(s) important for meta-analysis | 6 (8.1%) |
| The preprint reports interim results and publication reports complete results | 4 (5.4%) |
| The publication lists one or more additional funding sources | 4 (5.4%) |
| The publication includes SAP/protocol | 3 (4.1%) |
| Resulted in change in RoB (selective reporting) | 1 (1.4%) |
| The publication reports additional information on trial status | 2 (2.7%) |
| Publication and preprint report different types of analyses (i.e., ITT vs. PP) | 2 (2.7%) |
| Resulted in change in RoB (missing outcome data) | 1 (1.4%) |
| The preprint reports outcome for an unspecified subgroup whereas the publication reports outcome data for the full randomized population | 1 (1.4%) |
| Resulted in change in RoB (selective reporting) | 1 (1.4%) |
| The publication reports additional information on missing outcome data | 1 (1.4%) |
| Resulted in change in RoB (missing outcome data) | 1 (1.4%) |
| The publication reports a trial name | 1 (1.4%) |
| The preprint reported an incorrect trial registration | 1 (1.4%) |
| The publication lists an additional country | 1 (1.4%) |
| The number of participants randomized changed between preprint and publication | 1 (1.4%) |
| The publication reports stratified results based on allocation by randomization versus preference whereas the preprint reports results for all patients | 1 (1.4%) |
| The publication reports additional details about the intervention | 1 (1.4%) |
| Results | **Number (%)** |
| Change in outcome data | 20 (27%) |
| The publication reports one or more additional outcome(s) | 11 (14.9%) |
| The preprint reports one or more additional outcome(s) | 6 (8.1%) |
| The preprint excluded patients from analysis that discontinued treatment but the publication included them | 1 (1.4%) |
| The preprint and publication report one or more outcomes at different timepoints | 1 (1.4%) |
| The proportion of data that was missing changed | 1 (1.4%) |

### Supplement 6 – Forest plots for meta-analyses including and excluding preprints

**Corticosteroids for mortality**

**1 month**

*Without preprints*

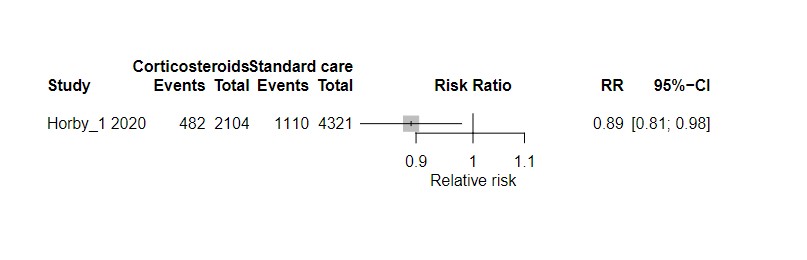

*With preprints*

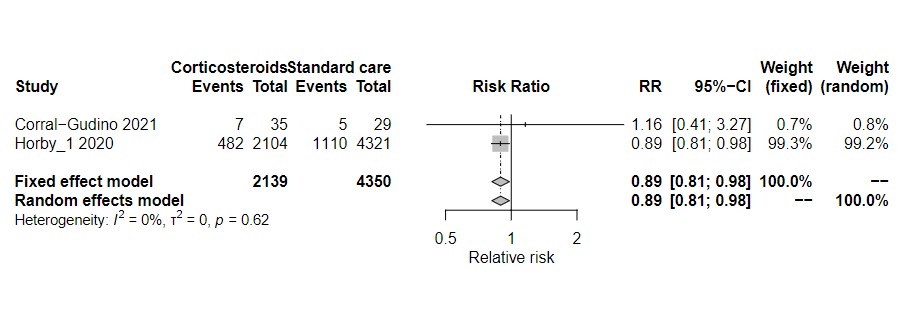

**3 months**

*Without pre-prints*

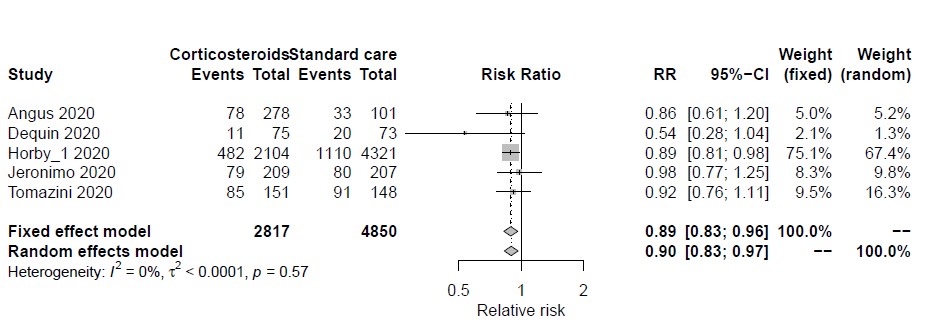

*With pre-prints*

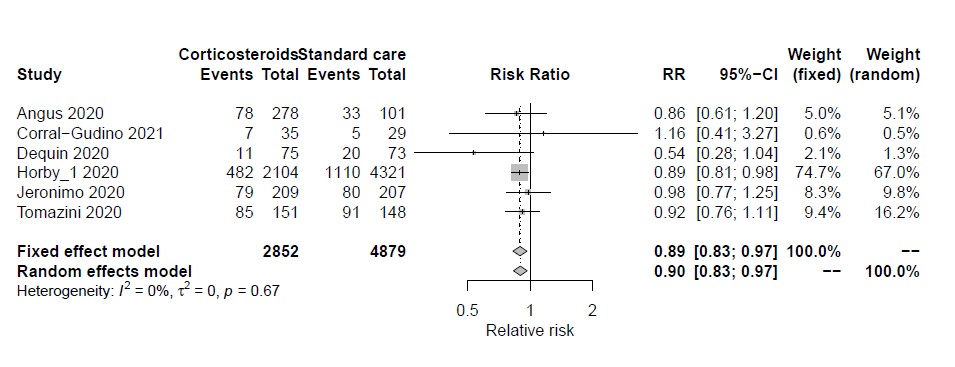

**6 months**

Without pre-prints

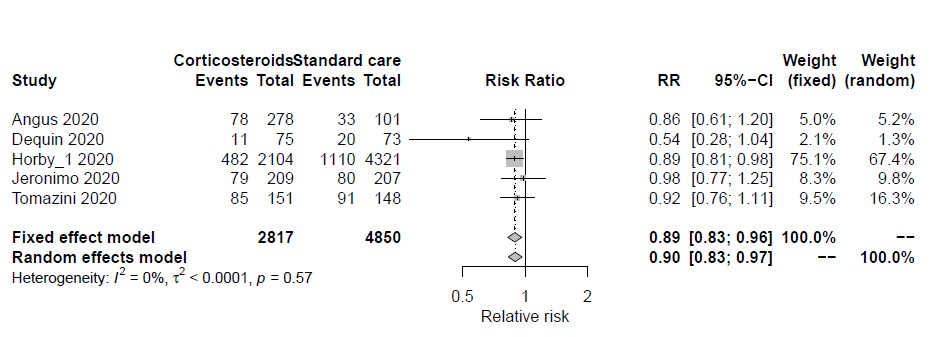

*With pre-prints*

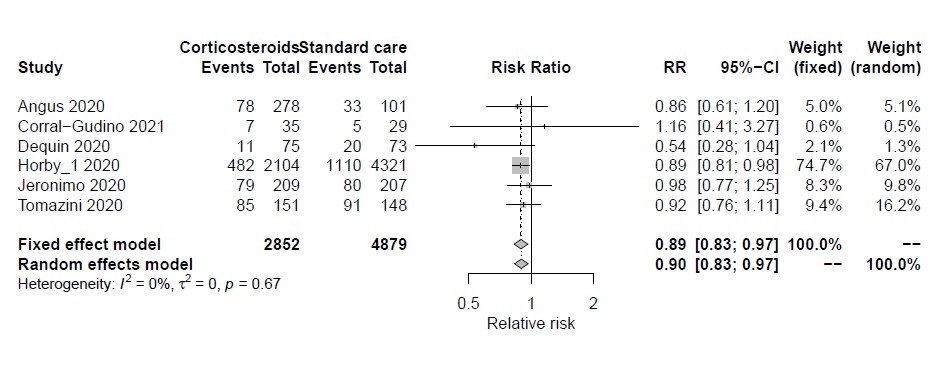

**Current**

*Without pre-prints*

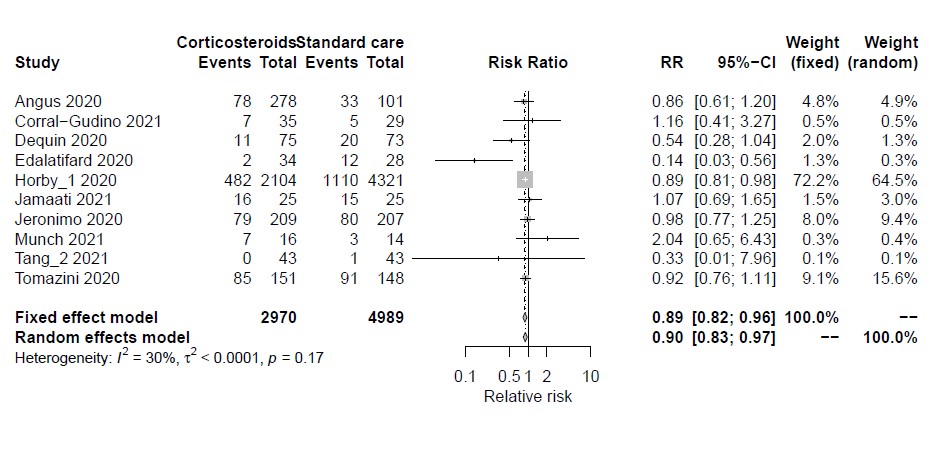

*With pre-prints*

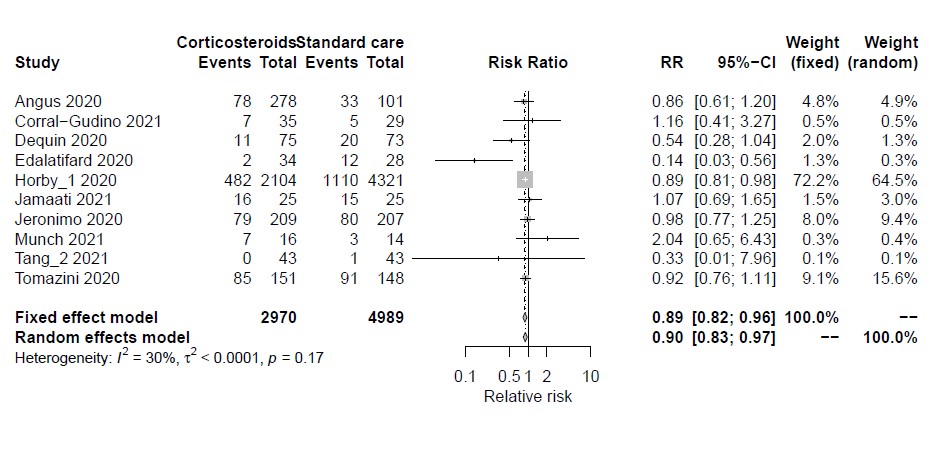

**Remesivir for mortality**

**1 month**

*Without pre-prints*

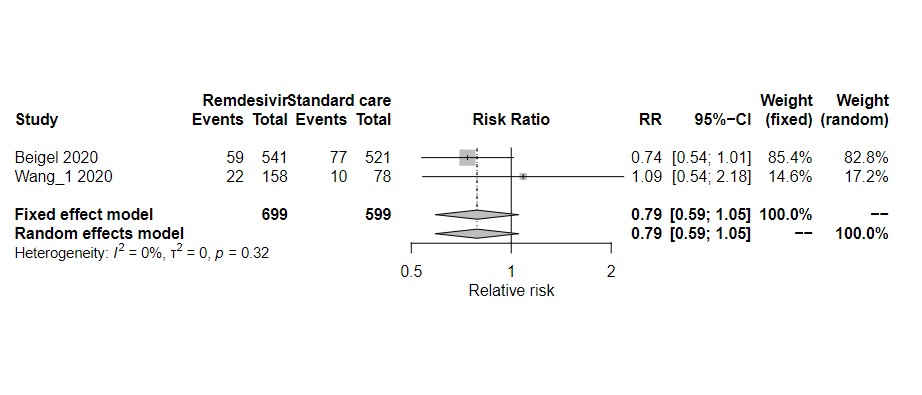

*With pre-prints*

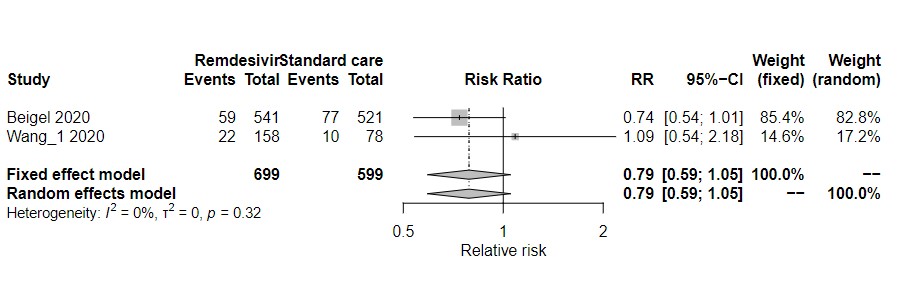

**3 months**

*Without pre-prints*

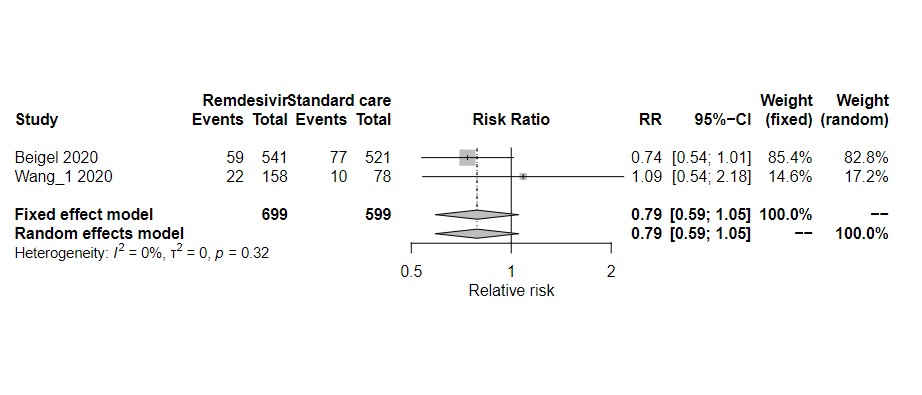

*With pre-prints*

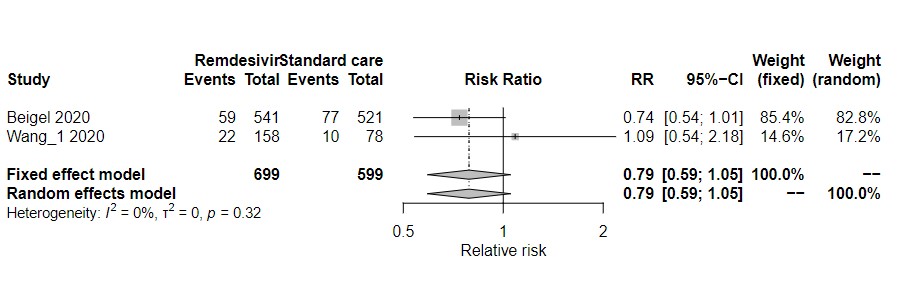

**6 months**

*Without pre-prints*

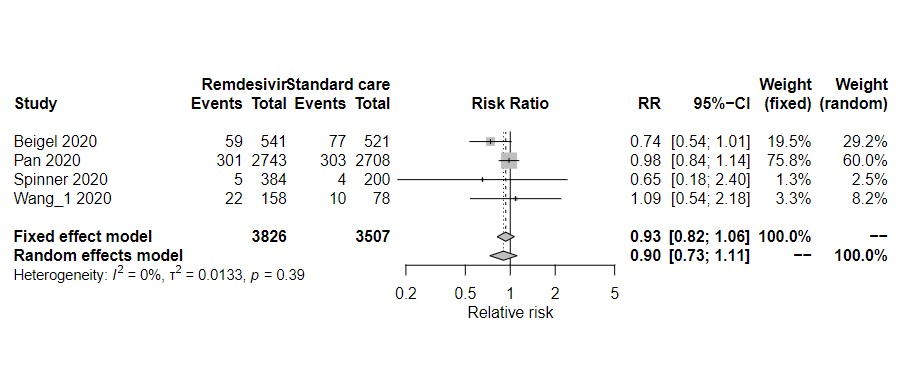

*With pre-prints*

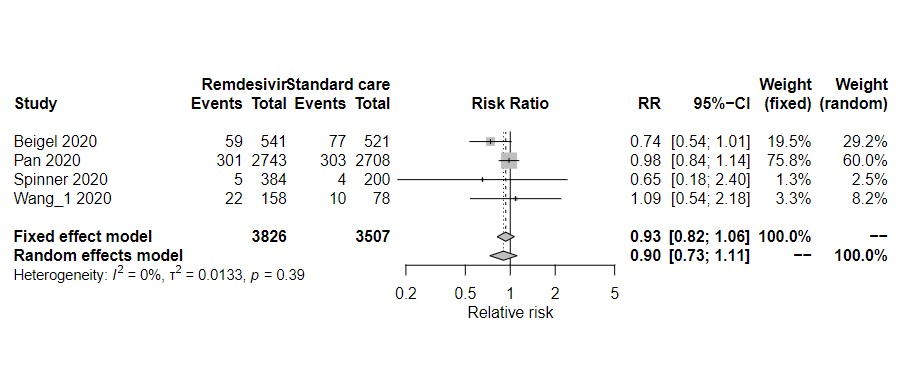

*Without pre-prints*

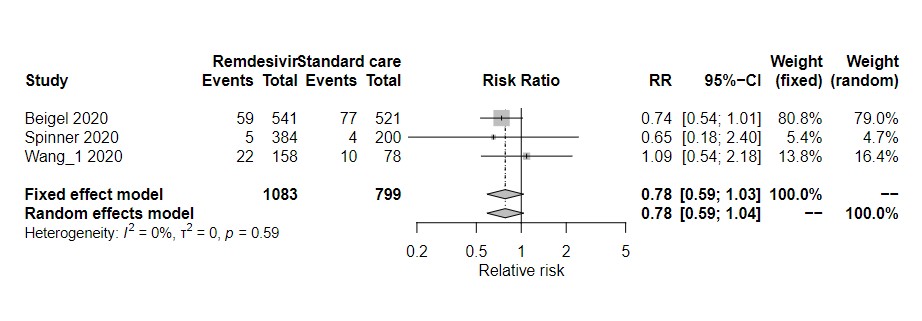

**Current:**

*With pre-prints*

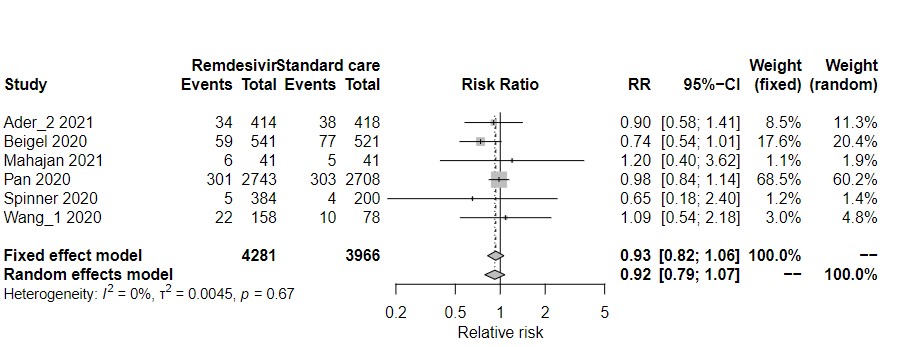

*Without pre-prints*

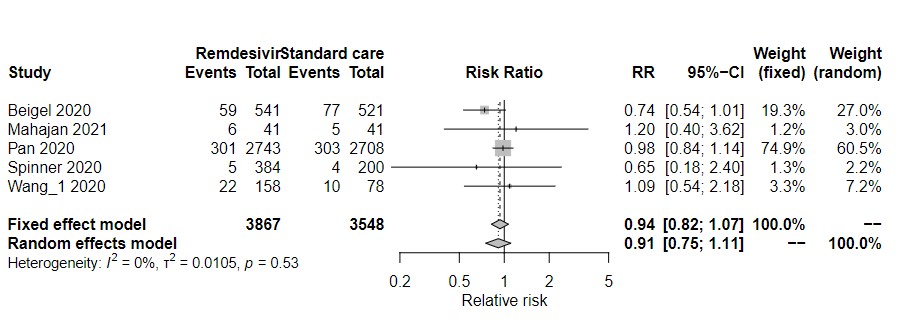

**Lopinavir-ritonavir for mortality**

**1 month**

*With pre-prints*

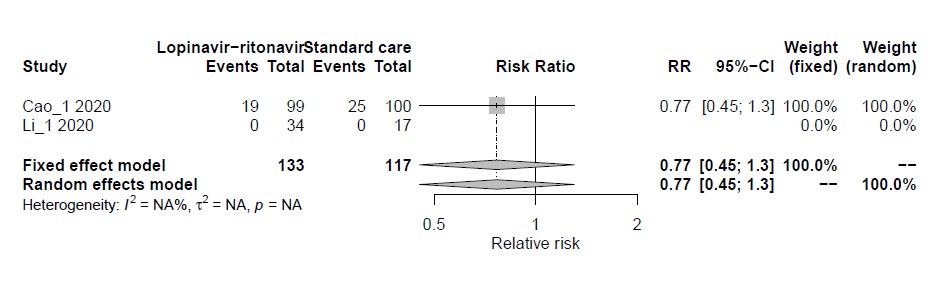

*Without pre-prints*

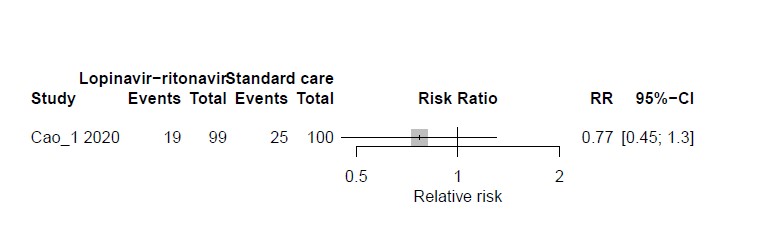

**3 months**

*With pre-prints*

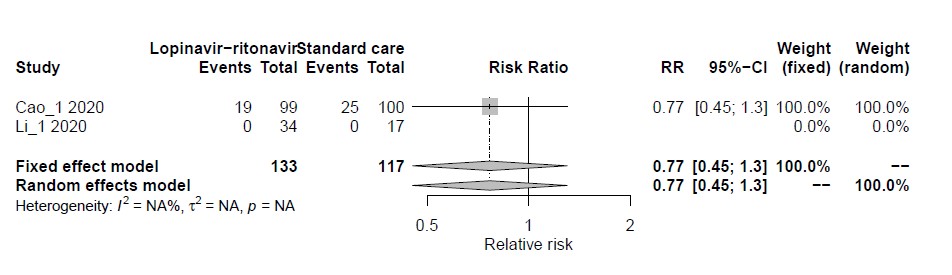

*Without pre-prints*

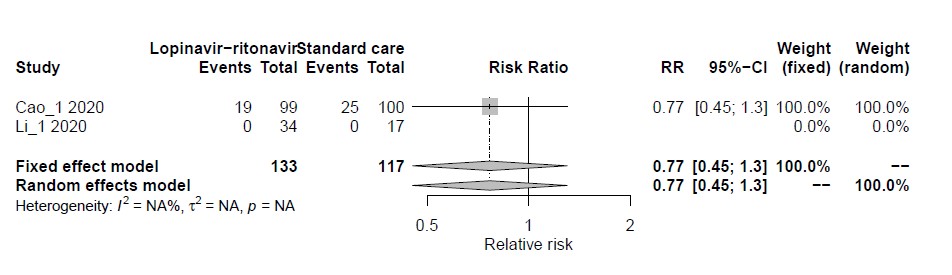

**6 months**

*With pre-prints*

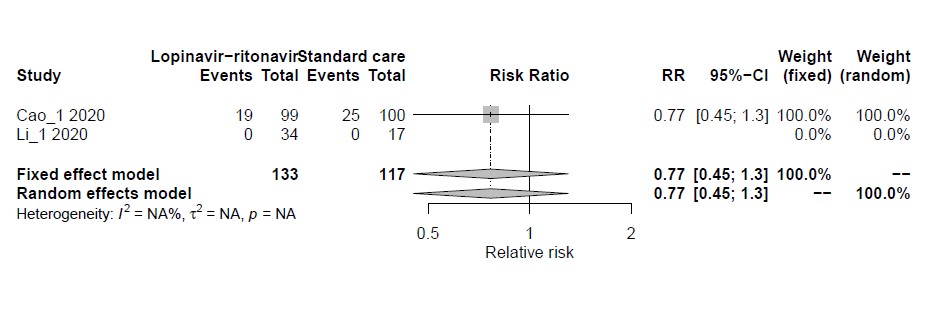

*Without pre-prints*

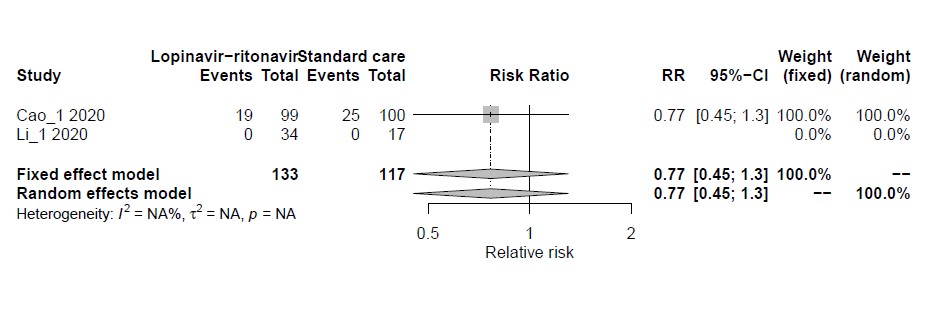

**Current**

*With pre-prints*

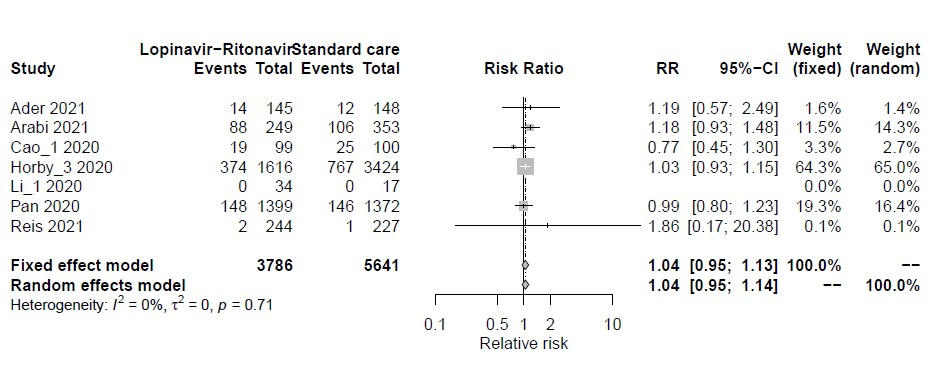

*Without pre-prints*

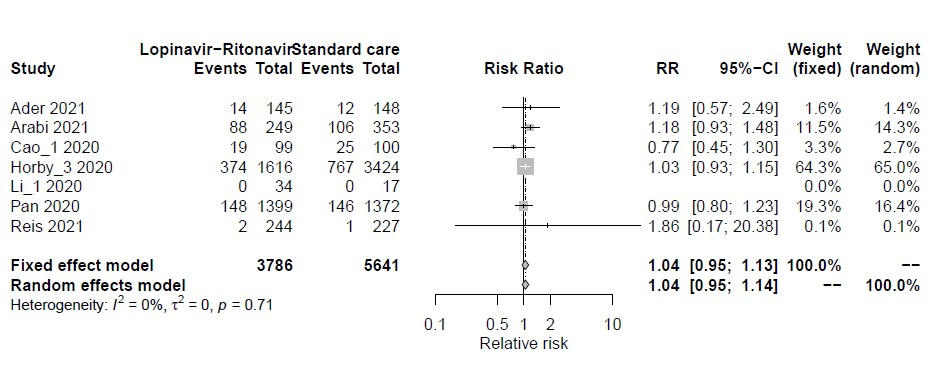

**(Hydroxy)chloroquine (treatment) for mortality**

**1 month**

*With pre-prints*

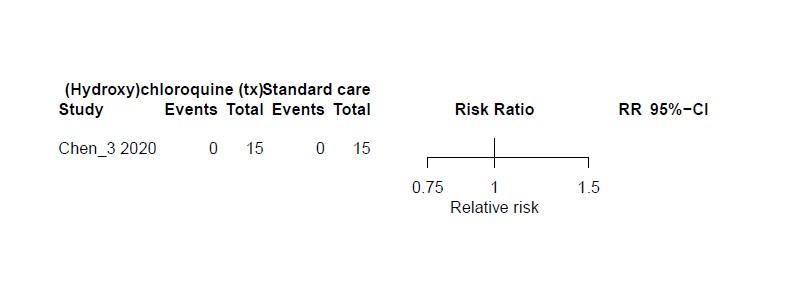

*Without pre-prints*

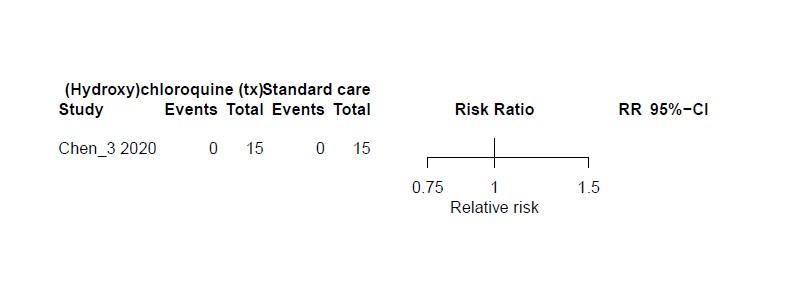

**3 months**

*With pre-prints*

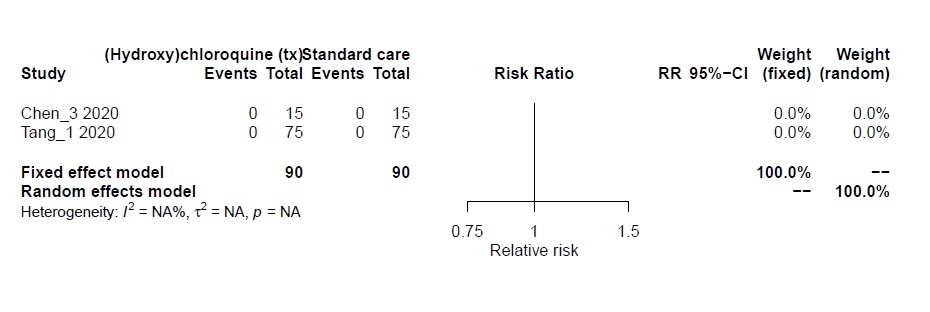

*Without pre-prints*

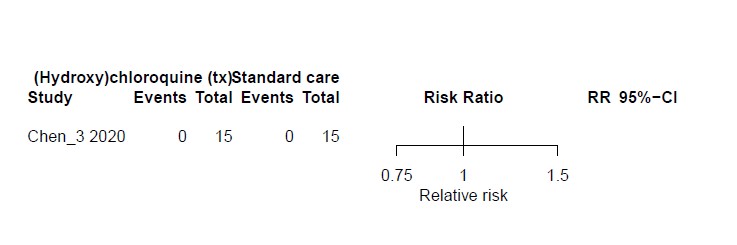

**6 months**

*With pre-prints*

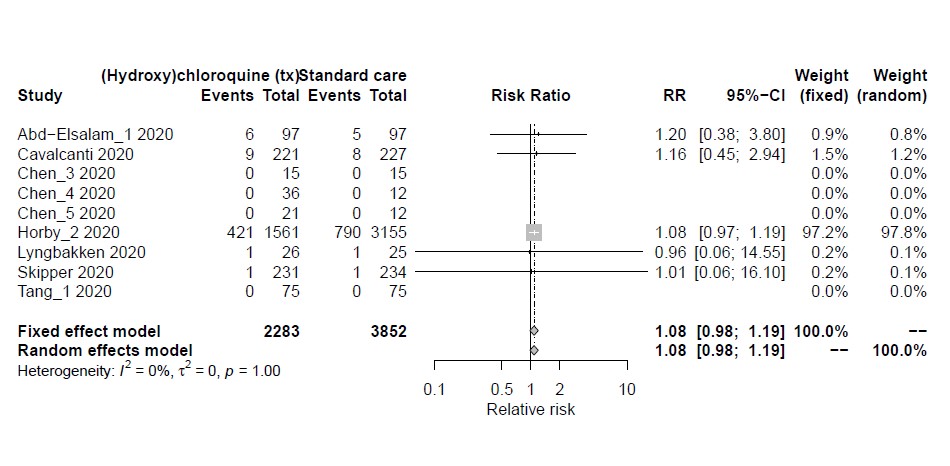

*Without pre-prints*

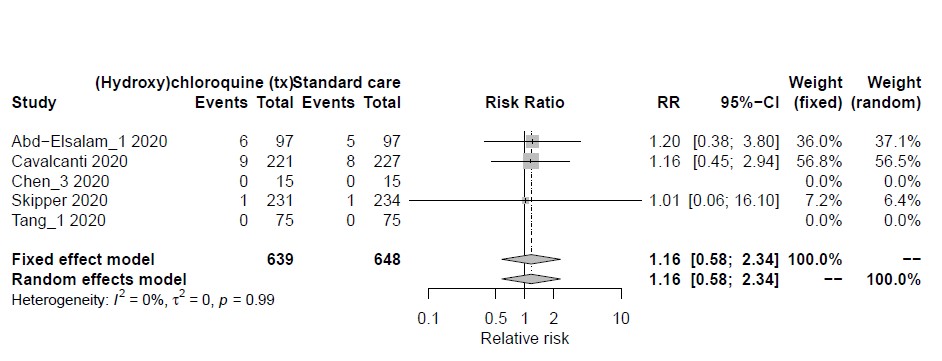

**Current**

*With pre-prints*

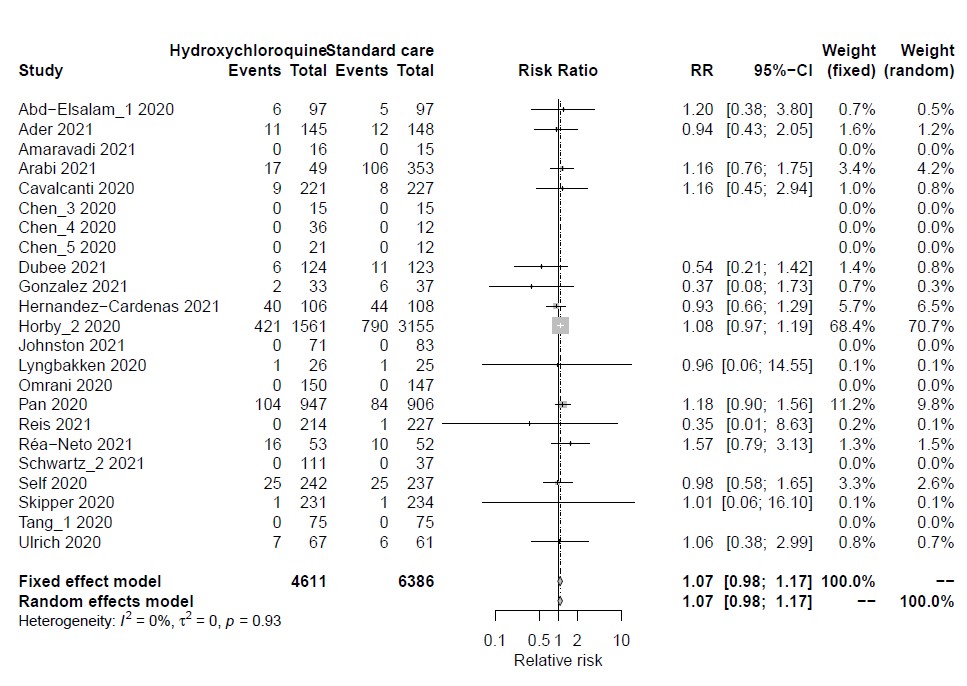

*Without pre-prints*

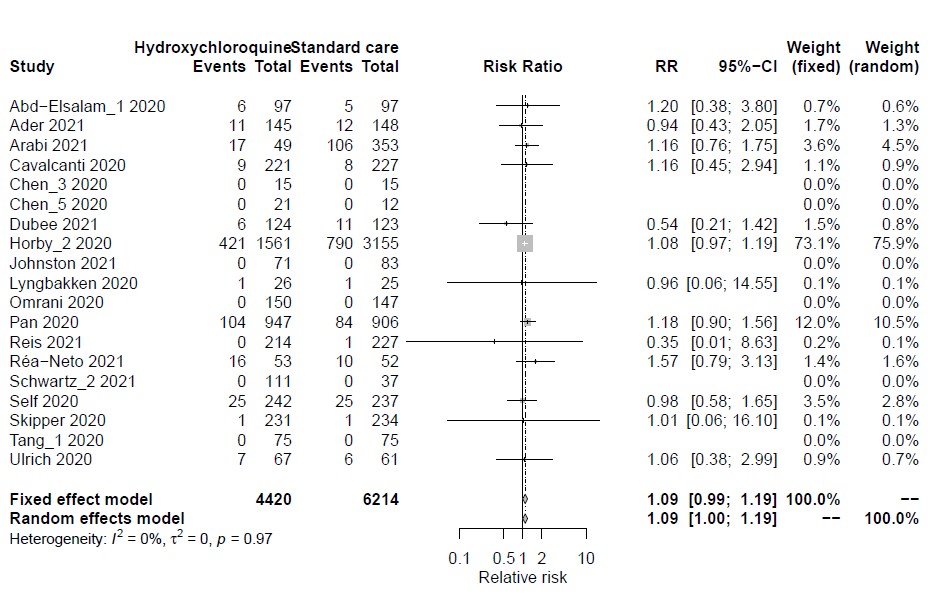

**Ivermectin for mortality**

**1 month**

*With pre-prints*

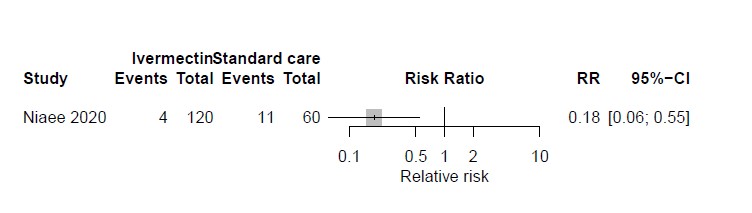

*Without pre-prints*

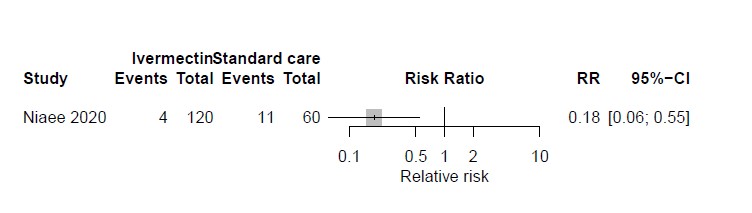

**3 months**

*With pre-prints*

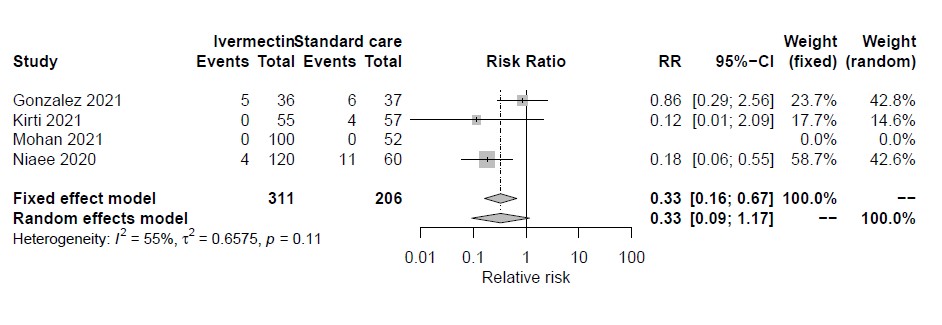

*Without pre-prints*

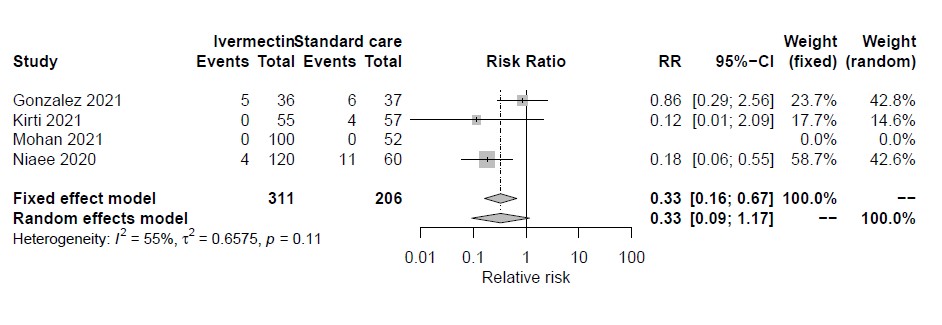

**6 months**

*With pre-prints*

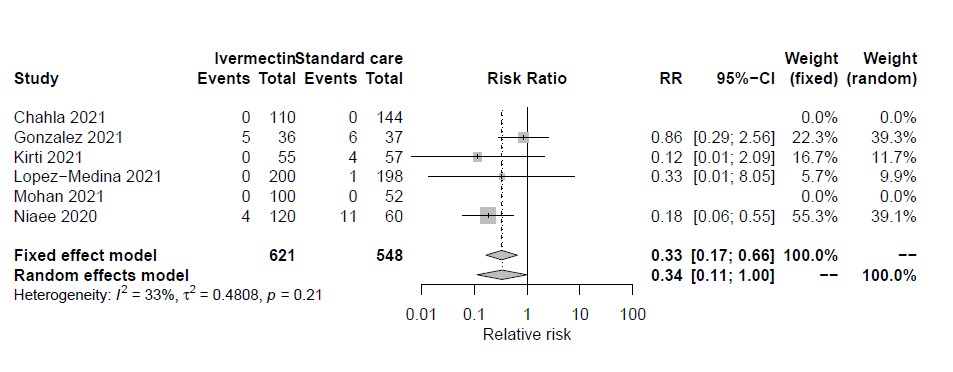

*Without pre-prints*

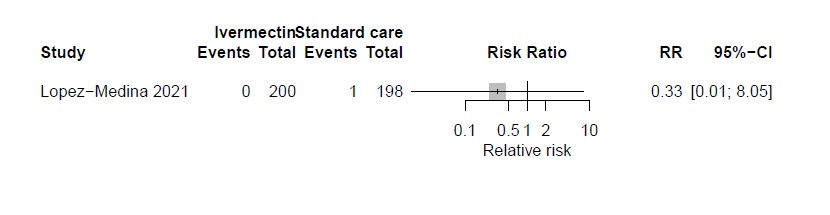

**Current**

*With pre-prints*

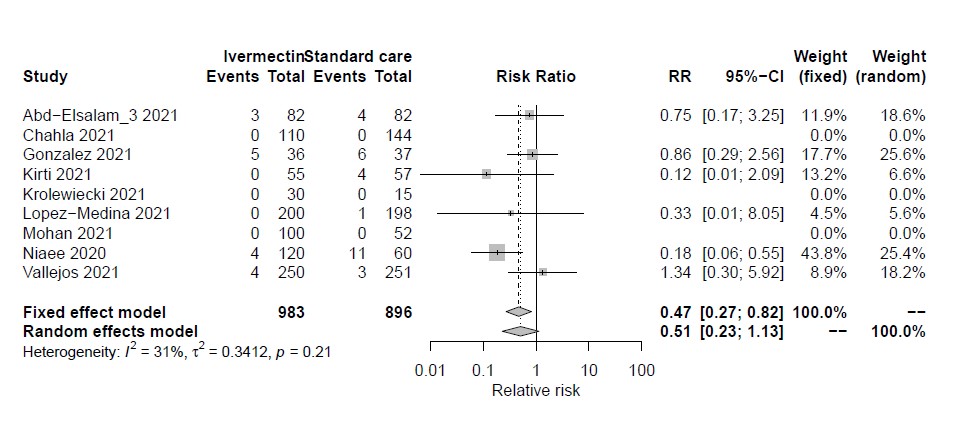

*Without pre-prints*

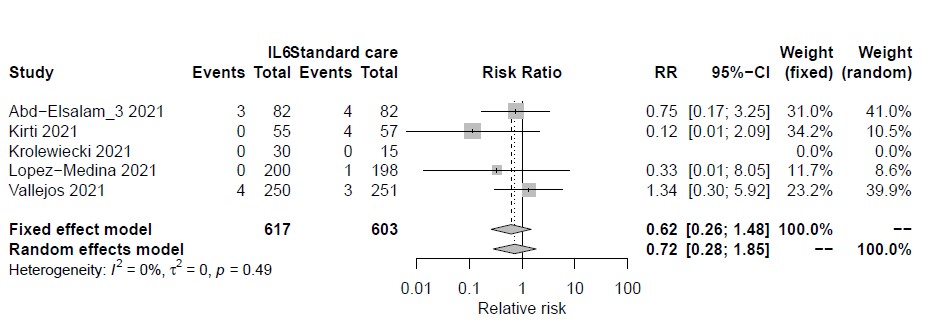

**IL-6 receptor blockers for mortality**

**1 month**

*With pre-prints*

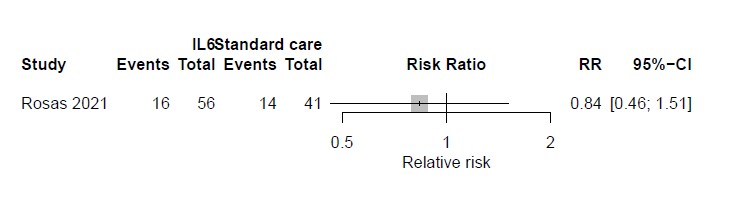

*Without pre-prints*

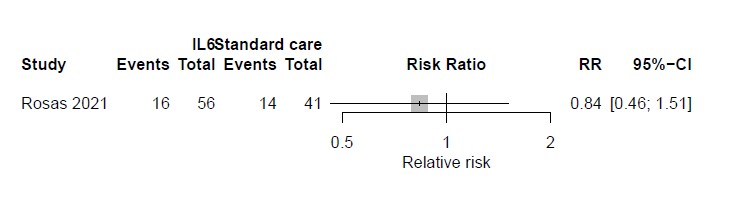

**3 months**

*With pre-prints*

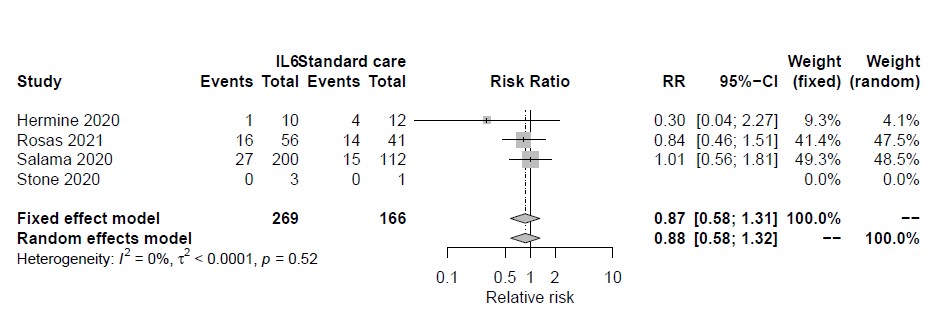

*Without pre-prints*

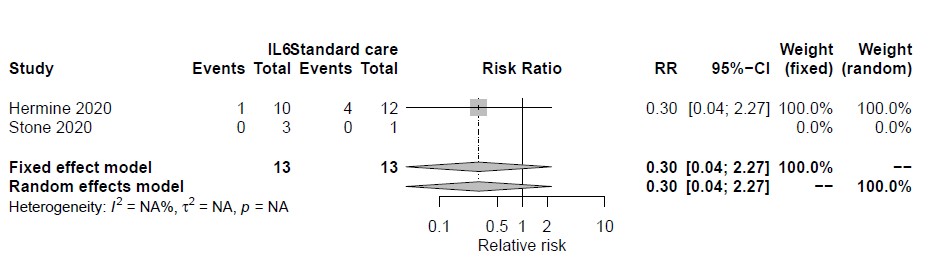

**6 months**

*With pre-prints*

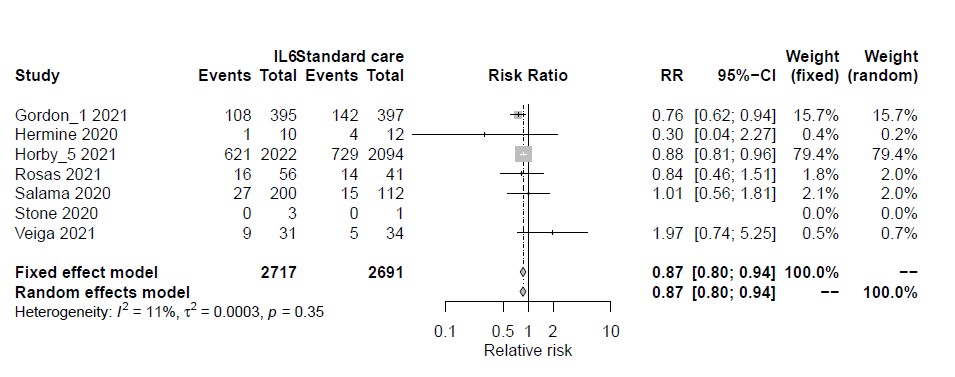

*Without pre-prints*

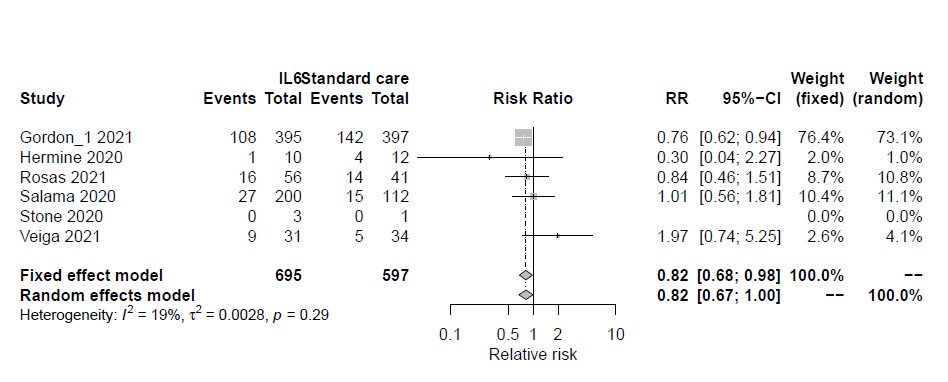

**Current**

*With pre-prints*

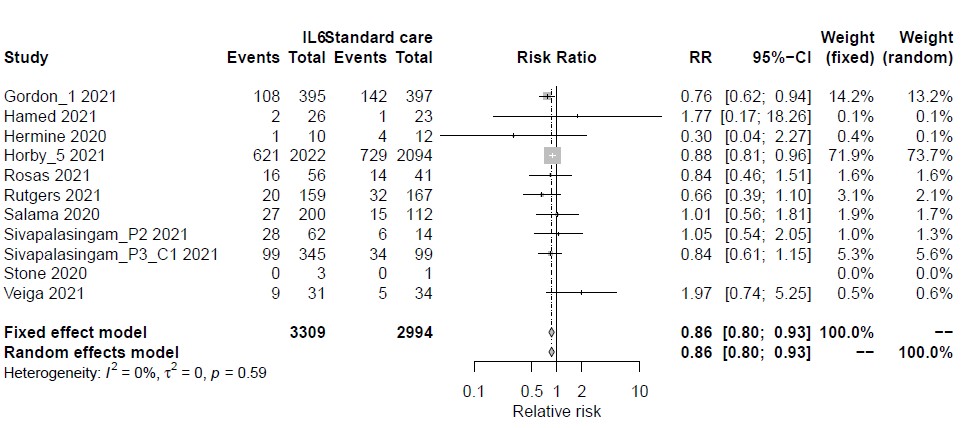

*Without pre-prints*

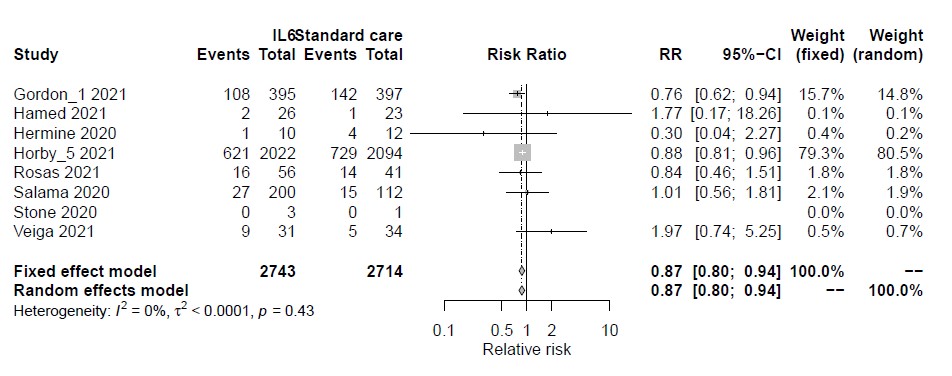

**Convalescent plasma for mortality**

**1 month**

*With pre-prints*

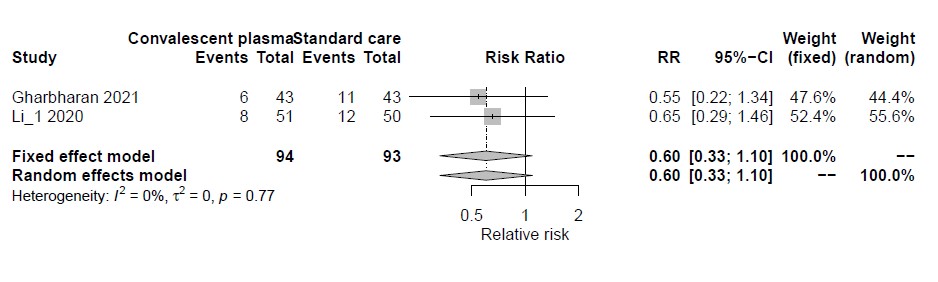

*Without pre-prints*

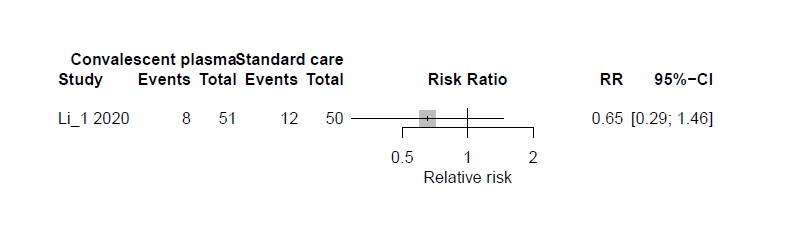

**3 months**

*With pre-prints*

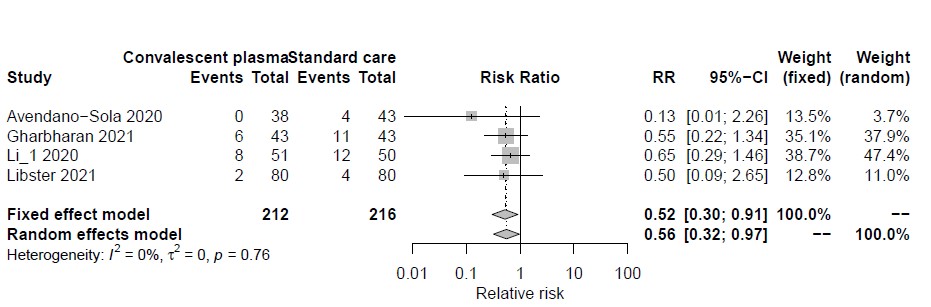

*Without pre-prints*

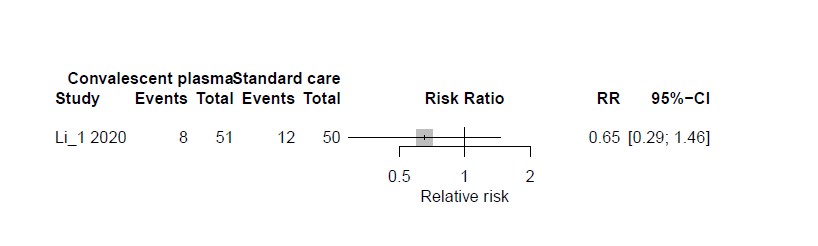

**6 months**

*With pre-prints*

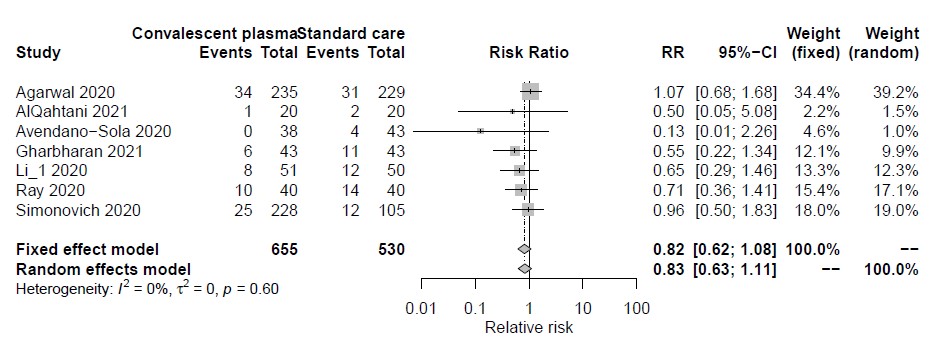

*Without pre-prints*

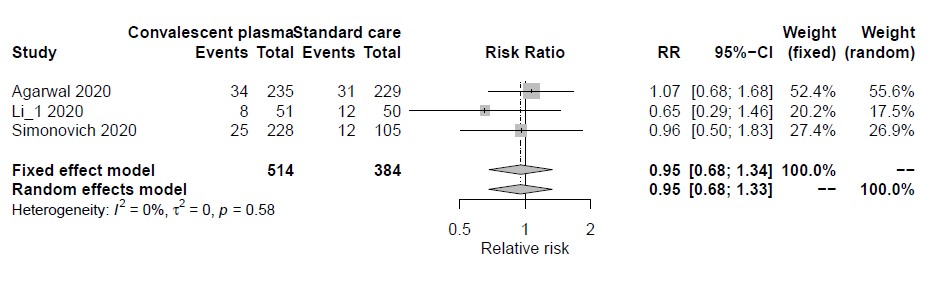

**Current**

*With pre-prints*

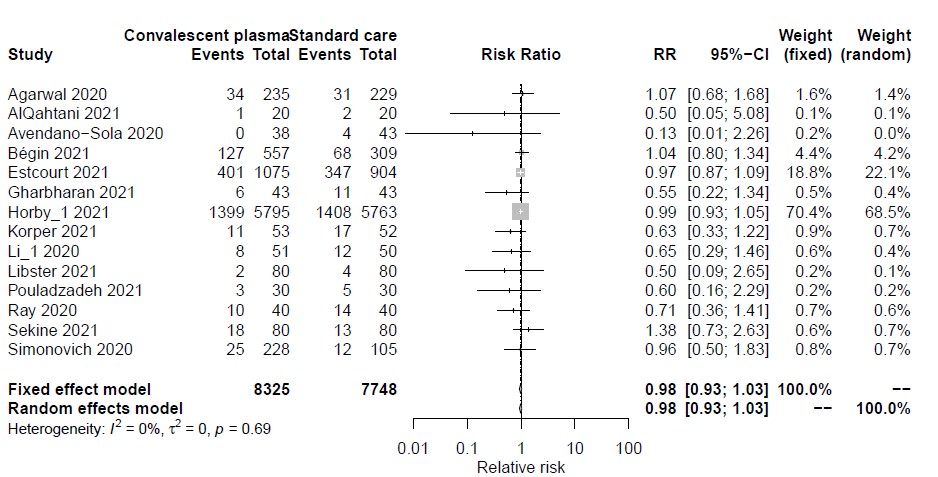

*Without pre-prints*

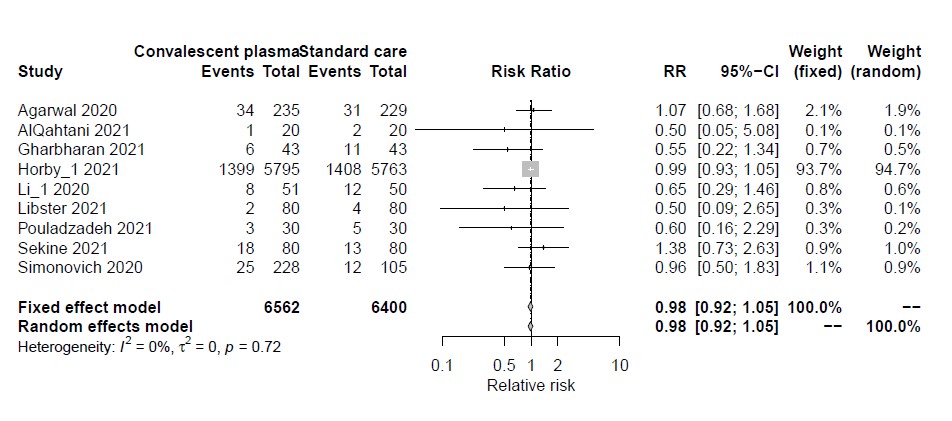

**(Hydroxy)chloroquine (prophylaxis) for mortality**

**1 month**

*With pre-prints*

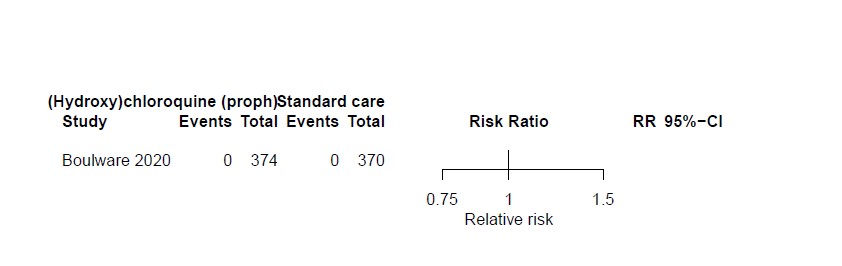

*Without pre-prints*

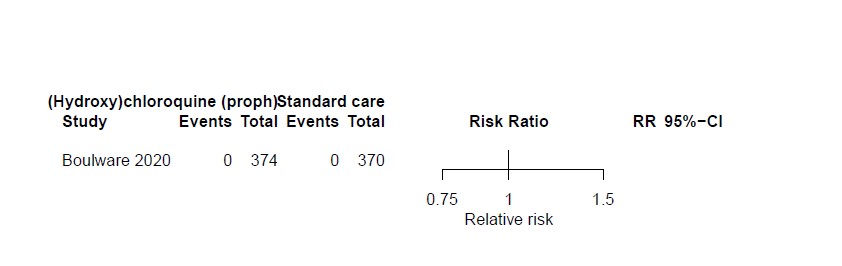

**3 months**

*With pre-prints*

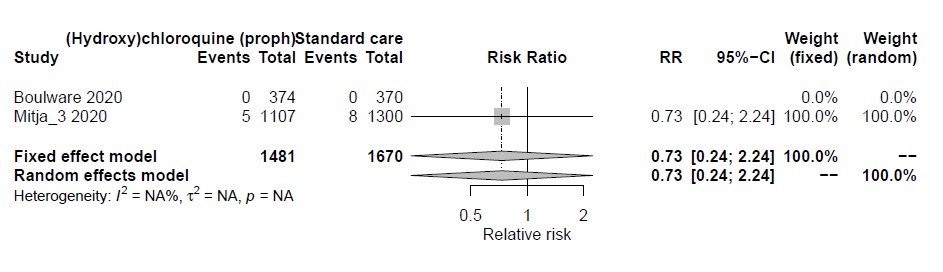

*Without pre-prints*

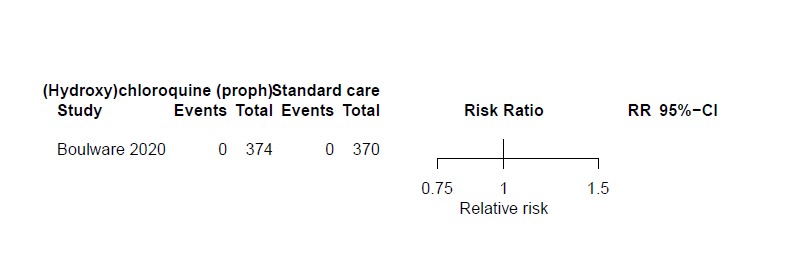

**6 months**

*With pre-prints*

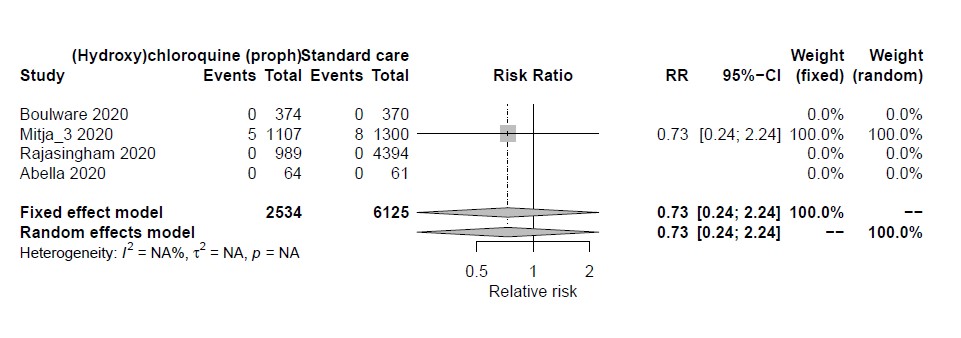

*Without pre-prints*

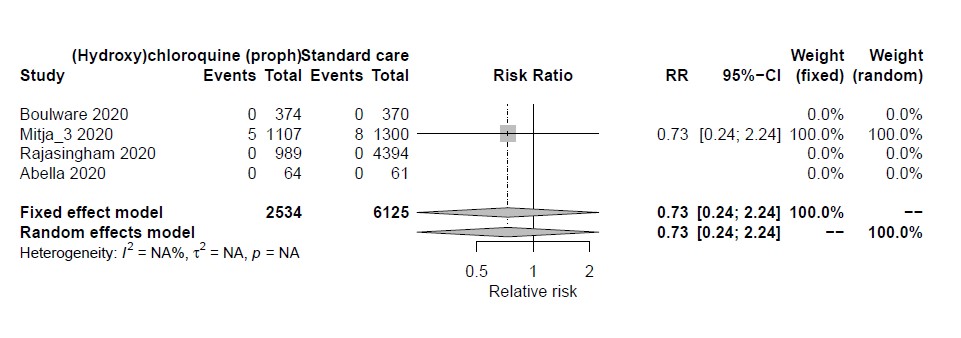

**Current**

*With pre-prints*

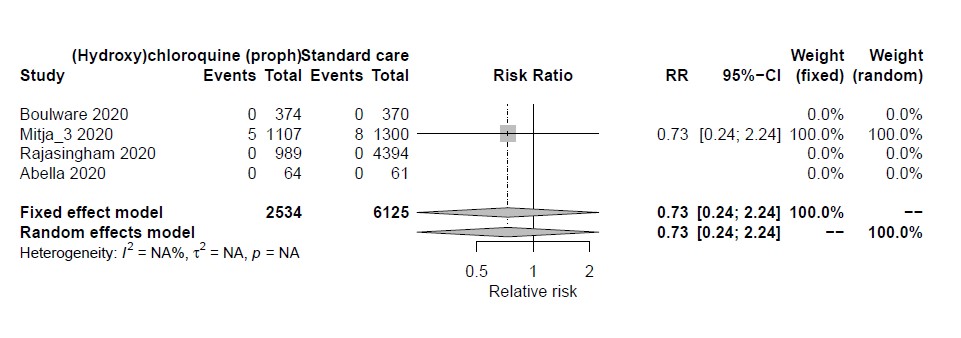

*Without pre-prints*

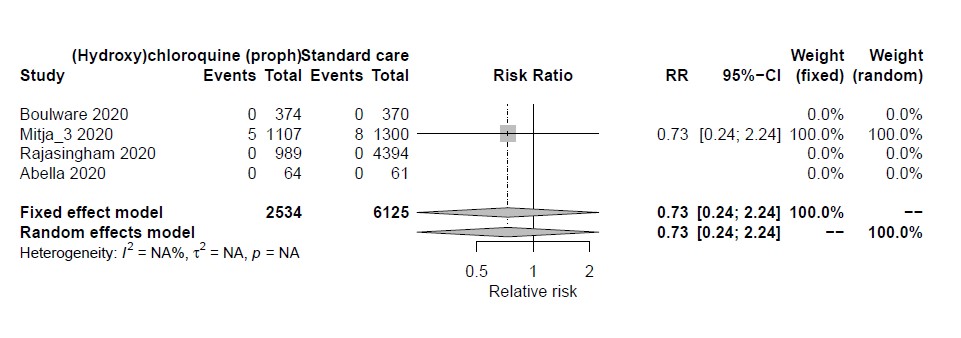

**Corticosteroids for mechanical ventilation**

**1 month**

*Without preprints*

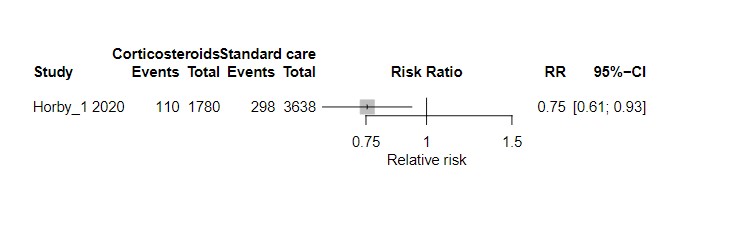

*With preprints*

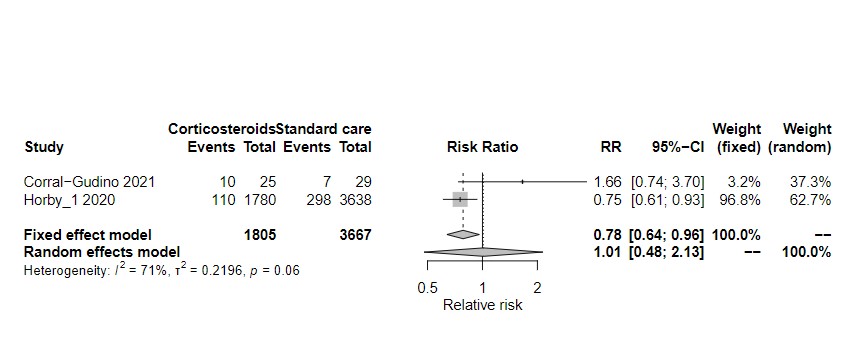

**3 months**

*Without preprints*

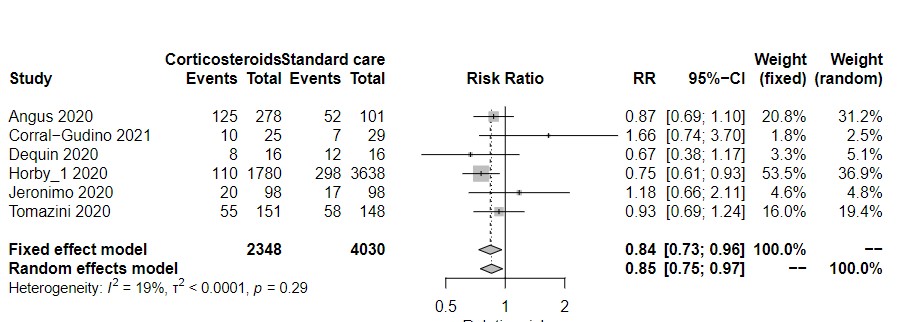

*With preprints*

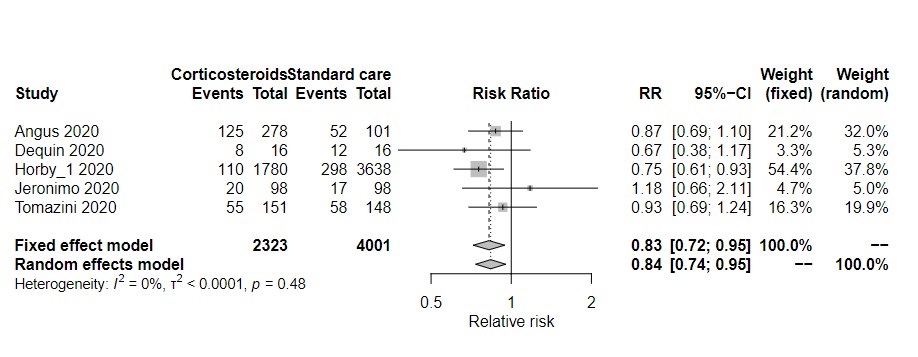

**6 months**

*Without preprints*

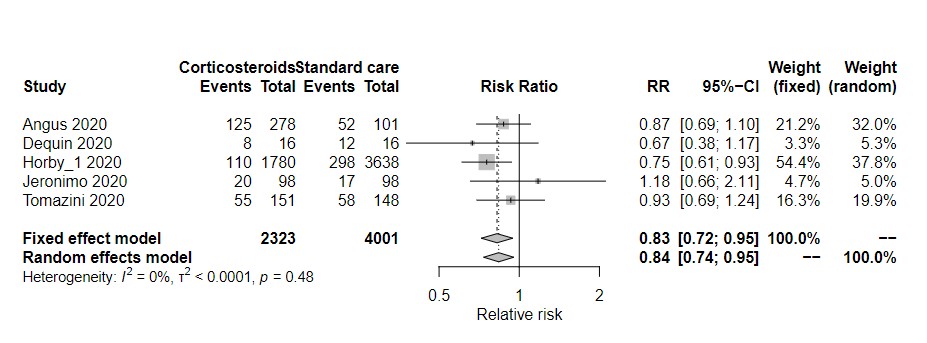

*With preprints*

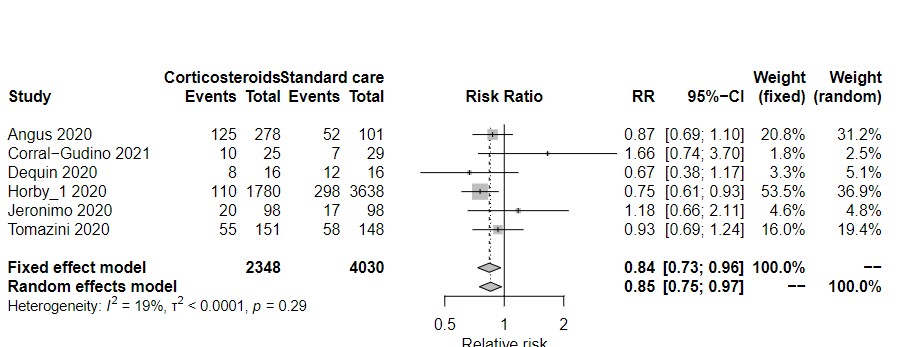

**Current**

*Without preprints*

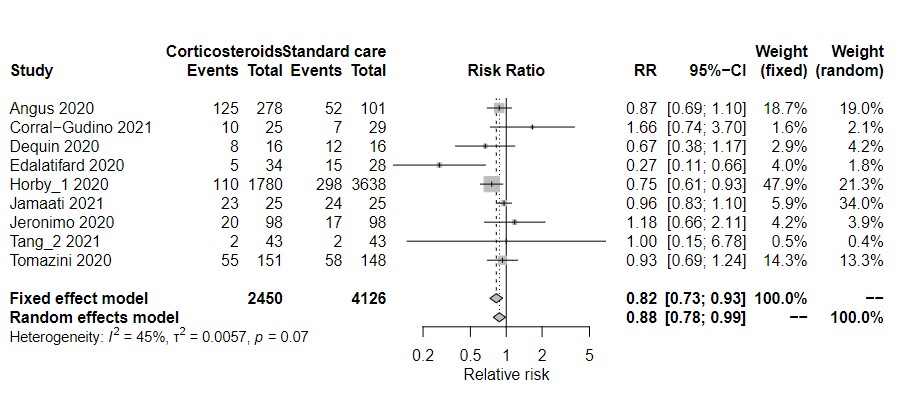

*With preprints*

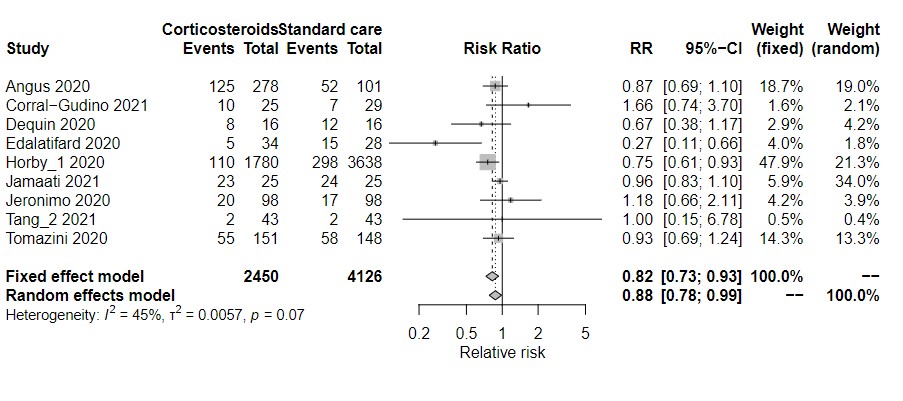

**Remesivir for mechanical ventilation**

**1 month**

*With preprints*

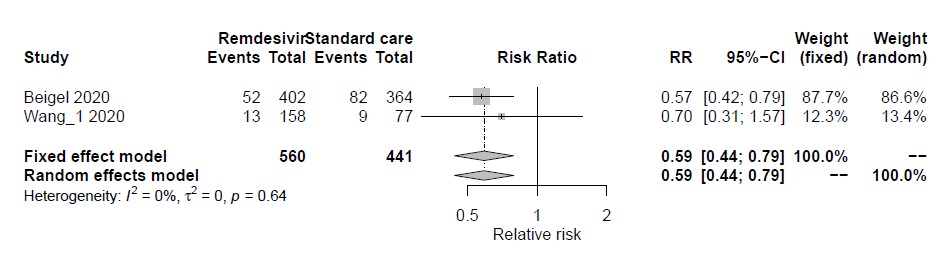

*Without preprints*

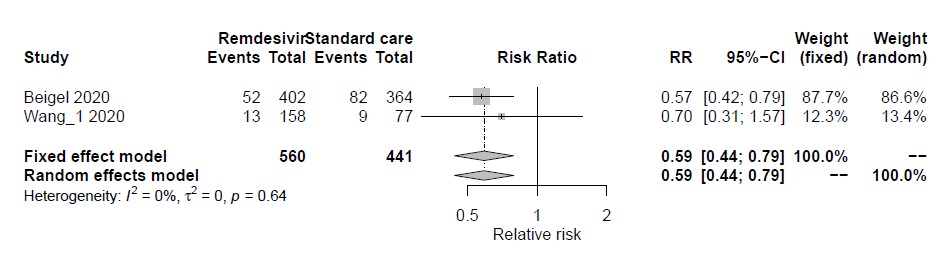

**3 months**

*With preprints*

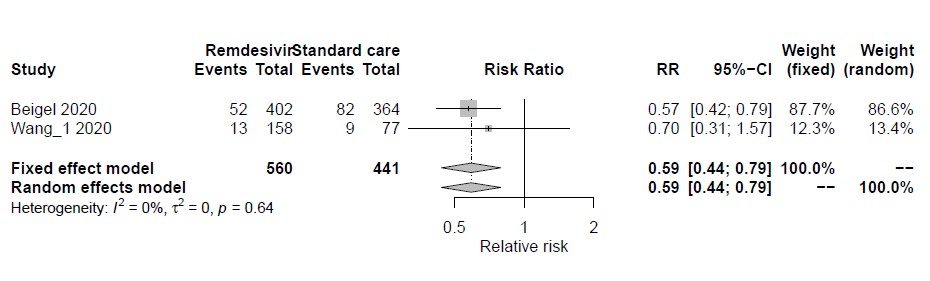

*Without preprints*

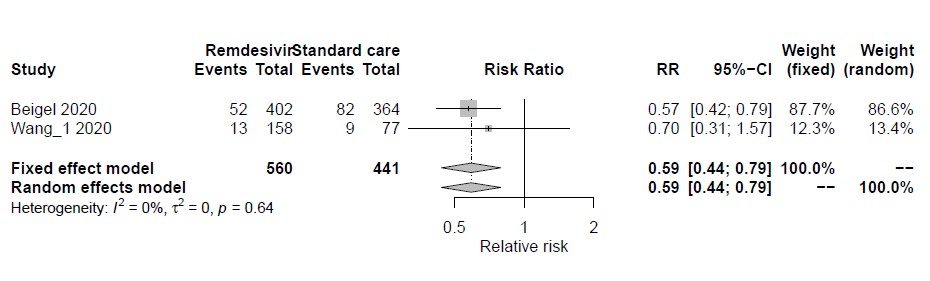

**6 months**

*With preprints*

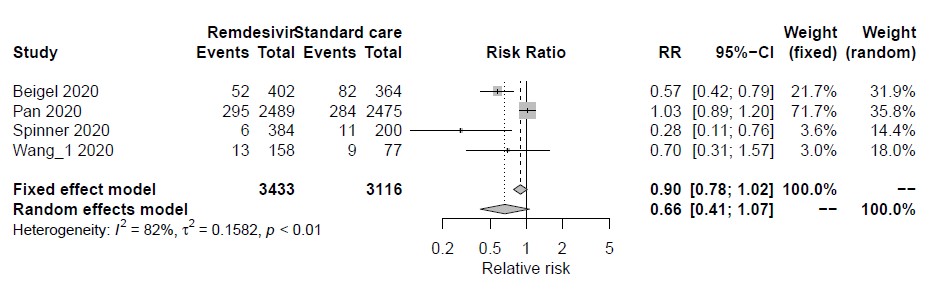

*Without preprints*

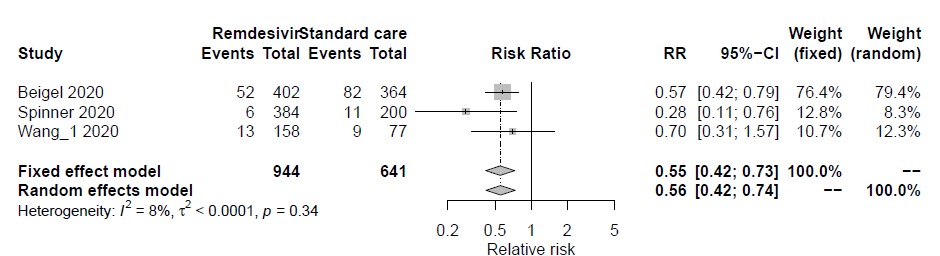

**Current**

*With preprints*

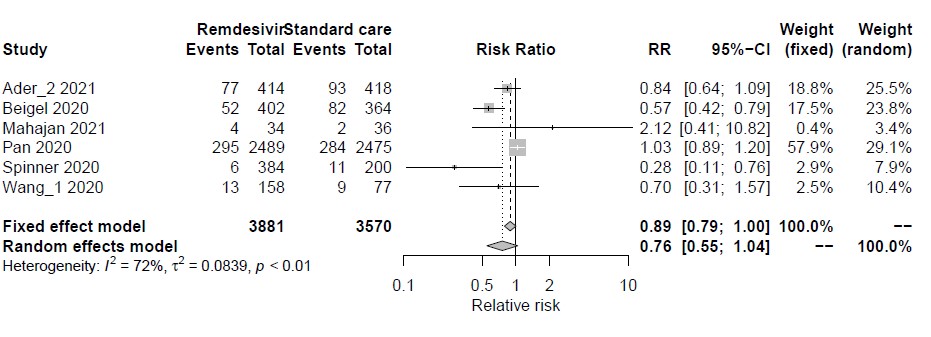

*Without preprints*

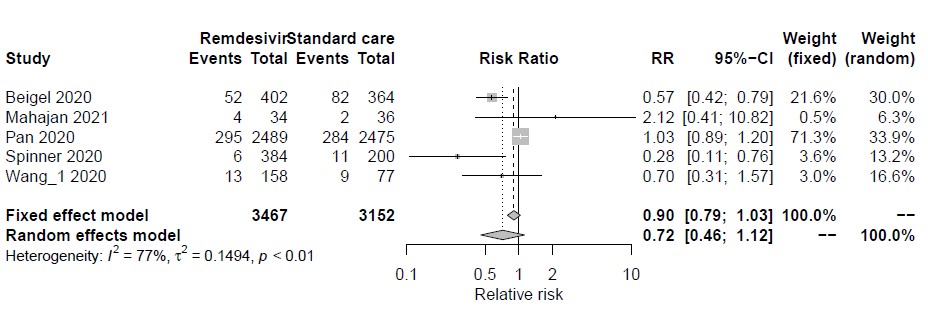

**Lopinavir-ritonavir for mechanical ventilation**

**1 month**

*With preprints*

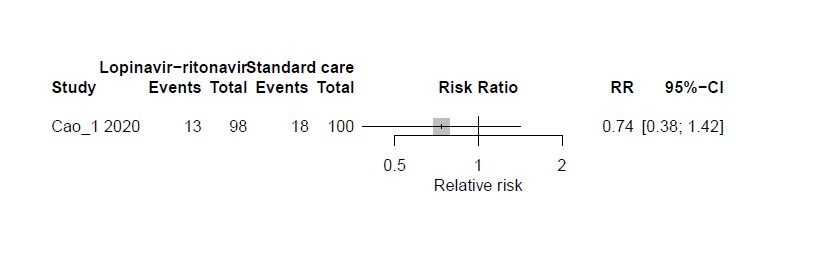

*Without preprints*

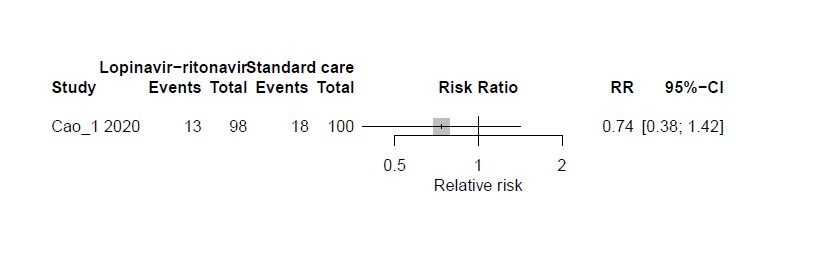

**3 months**

*With preprints*

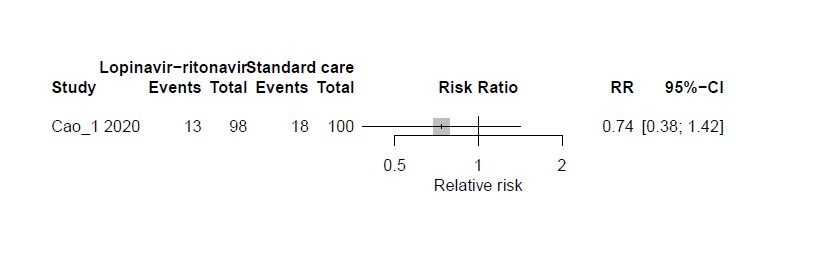

*Without preprints*

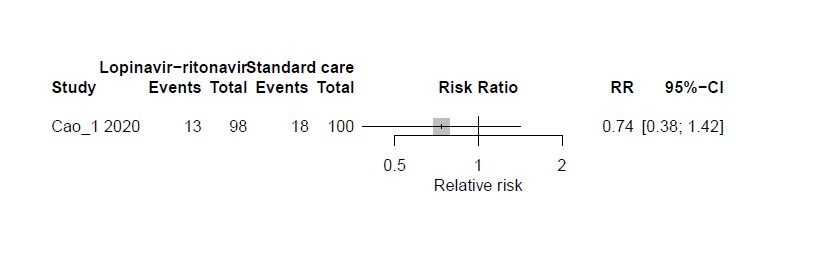

**6 months**

*With preprints*

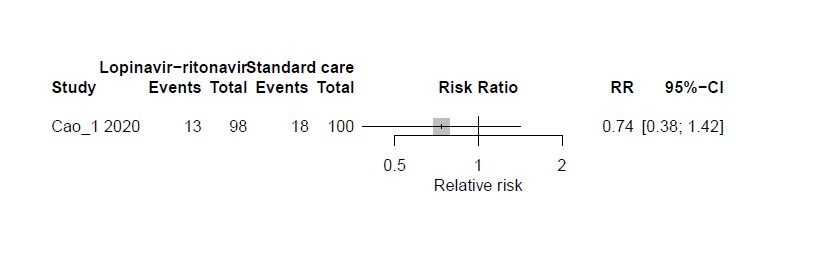

*Without preprints*

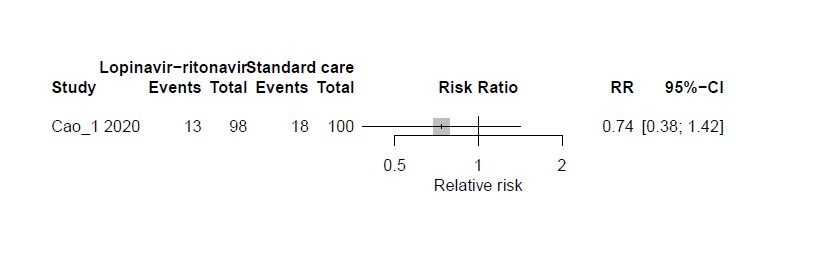

**Current**

*With preprints*

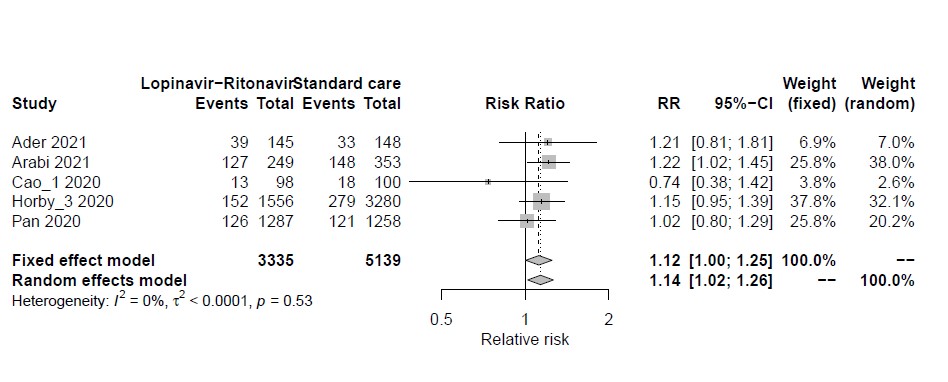

*Without preprints*

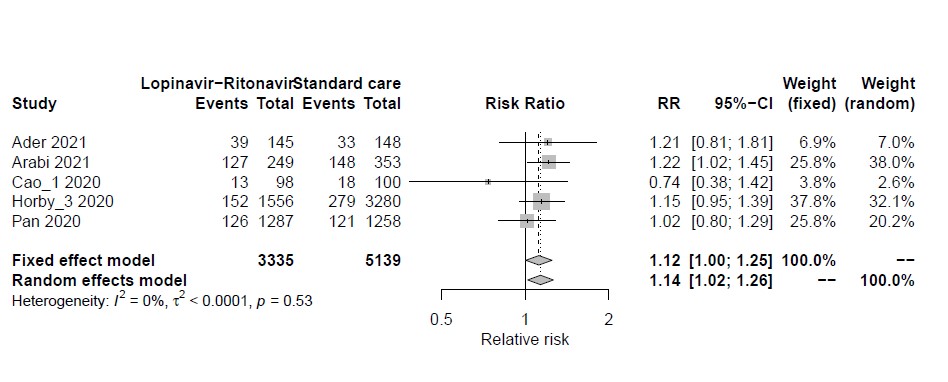

**Hydroxy(chloroquine) (treatment) for mechanical ventilation**

**1 month**

*With preprints*

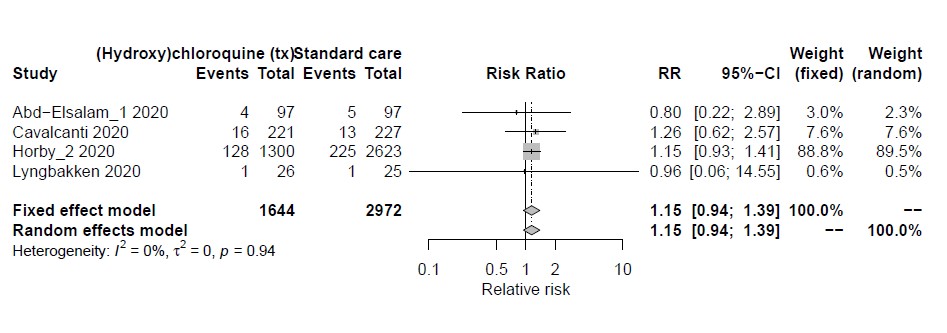

*Without preprints*

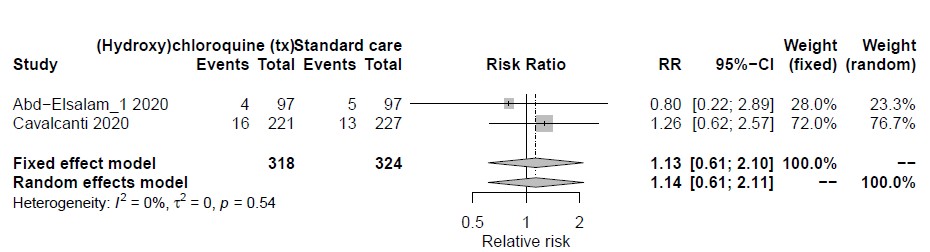

**3 months**

*With preprints*

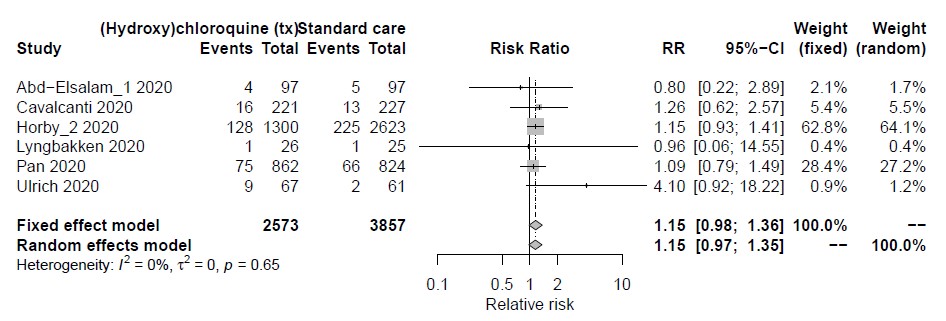

*Without preprints*

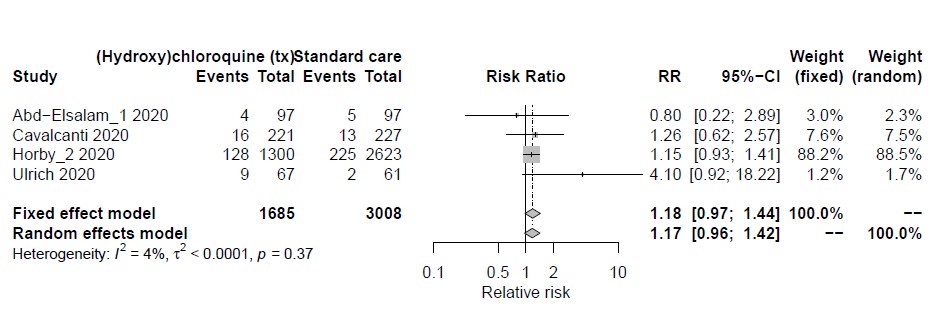

**6 months**

*With preprints*

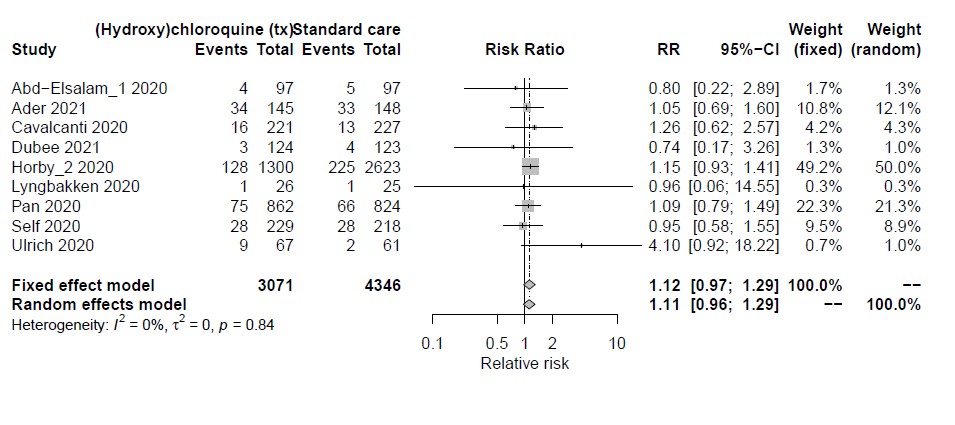

*Without preprints*

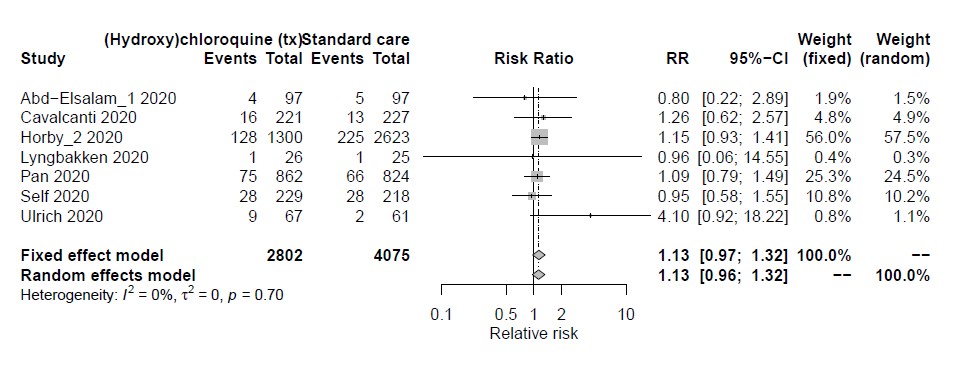

**Current**

*With preprints*

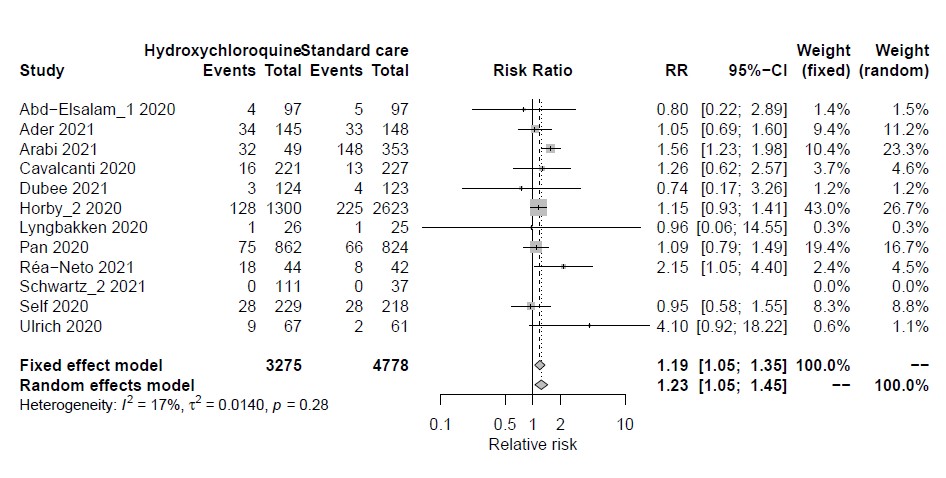

*Without preprints*

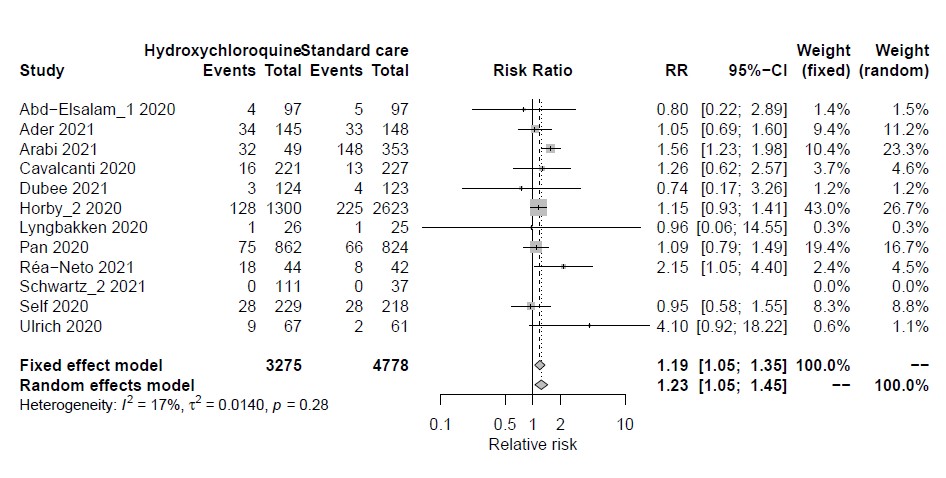

**Ivermectin for mechanical ventilation**

**1 month**

*With preprints*

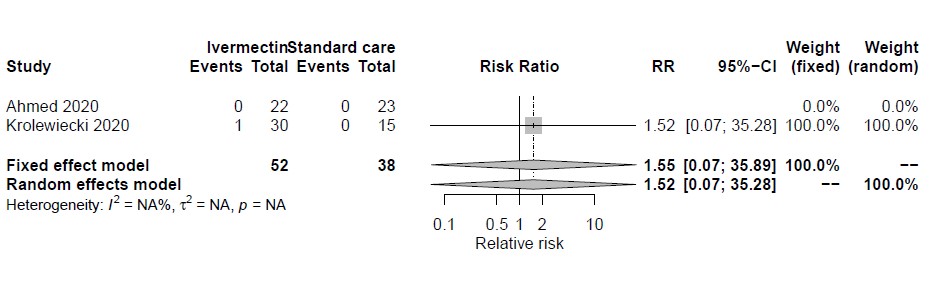

*Without preprints*

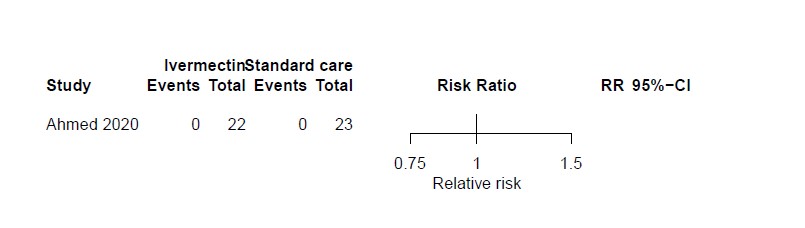

**3 months**

*With preprints*

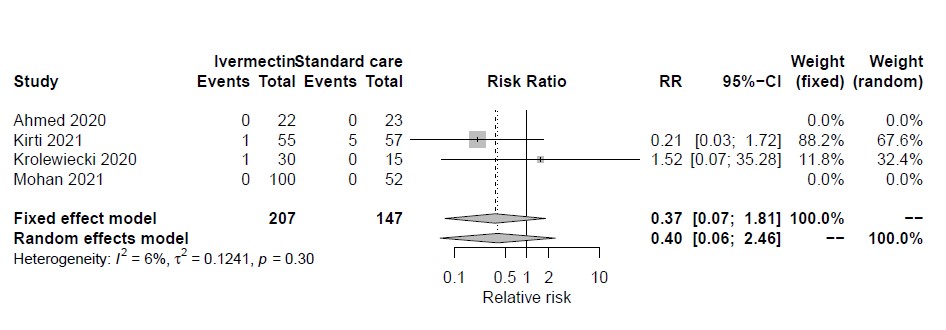

*Without pre-prints*

**6 months**

*With preprints*

*Without preprints*

**Current**

*With preprints*

*Without preprints*

**IL-6 receptor blockers for mechanical ventilation**

**1 month**

*With preprints*

*Without preprints*

**3 months**

*With preprints*

*Without preprints*

**6 months**

*With preprints*

*Without preprints*

**Current**

*With preprints*

*Without preprints*

**Convalescent plasma for mechanical ventilation**

**1 month**

*With preprints*

*Without preprints*

**3 months**

*With preprints*

*Without preprints*

**6 months**

*With preprints*

*Without preprints*

**Current**

*With preprints*

*Without preprints*
